# Addressing racial/ethnic disparities in the COVID-19 vaccination campaign

**DOI:** 10.1101/2021.04.21.21255878

**Authors:** Marissa B. Reitsma, Jeremy D. Goldhaber-Fiebert, Joshua A. Salomon

## Abstract

**Background:** As of April 19, all adults aged 16 years and older are eligible for COVID-19 vaccination. Unequal vaccination rates across racial/ethnic groups may compound existing disparities in cases, hospitalizations, and deaths among Black, Indigenous, and Hispanic communities.

**Methods:** From state websites, we extracted shares of people receiving ≥1 vaccine dose, stratified by age and separately by race/ethnicity, through March 31, 2021. Combining these data with demographic data from the American Community Survey, we estimated relative uptake rates by race/ethnicity within each state as the observed share of vaccinations for a racial/ethnic group, divided by the expected share if uptake across racial/ethnic groups within each age group were proportional to population size, an approach that allowed us to control for historical age-based eligibility. We modeled vaccination scale-up within each census tract in a state under three scenarios: 1) a scenario in which unequal uptake rates persist, 2) a scenario in which uptake rates are equalized across race/ethnicity groups over six weeks, and 3) a scenario in which uptake is equalized and states employ place-based allocation strategies that prioritizes disadvantaged census tracts.

**Results:** White adults received a disproportionate share of vaccinations compared to Black and Hispanic adults through March 31, 2021. Across states, relative uptake rates, adjusted for eligible population size, were a median 1.3 (IQR, 1.2-1.4) times higher for White compared to Black adults, and a median 1.4 (IQR, 1.2-1.8) times higher for White compared to Hispanic adults. Projecting vaccination coverage under persistence of current disparities in uptake, we found that Black and Hispanic populations would reach 75% coverage among adults almost one month later than White populations. In alternative scenarios, we found that interventions to equalize uptake rates across racial/ethnic groups could narrow but not erase these gaps, and that geographic targeting of vaccine doses to disadvantaged communities may be needed to produce a more equitable convergence of coverage by July.

**Discussion:** Interventions are urgently needed to eliminate disparities in COVID-19 vaccination rates. Eliminating access barriers and increasing vaccine confidence among marginalized populations can narrow gaps in coverage. Combining these interventions with place-based allocation strategies can accelerate vaccination in disadvantaged communities, who have borne a disproportionate burden from COVID-19.

## Introduction

As the COVID-19 vaccination campaign advances in the United States, unequal vaccination rates across racial/ethnic groups have compounded existing disparities in cases, hospitalizations, and deaths among Black, Indigenous, and Hispanic populations (1–3). Beginning in March, most states announced plans to expand eligibility to all adults ages 16 and older during the following weeks, and President Biden set a deadline for all US adults to be eligible by April 19, 2021. With this expansion of eligibility, equitable vaccine scale-up requires action to address the causes of differential coverage.

In this study, we quantify differential vaccine uptake rates by race/ethnicity within each US state in order to project racial/ethnic disparities in vaccine coverage through July 1, 2021, under a ‘*status quo*’ scenario of continued differential uptake, and alternative scenarios reflecting implementation of strategies to reduce disparities by addressing allocation, access, and acceptance.

## Methods

### Data Sources

We analyzed demographic data (population distribution by age, race/ethnicity, and census tract) from the American Community Survey (2015-2019) and vaccination data from state websites on the shares of people receiving at least one vaccine dose, stratified by age and separately by race/ethnicity, through March 31, 2021. We extracted data for the following race/ethnic groups, when available: American Indian or Alaska Native, Asian, Black, Hispanic, Native Hawaiian or Pacific Islander, White, and Other.

### Adjusting for Missing Values in Vaccination Data

Publicly available vaccination data from states have varying degrees of completeness of reporting on race/ethnicity and age. When computing the observed share of vaccinations by racial/ethnic group, we computed the share only among vaccinations with reported race/ethnicity, by state. This assumed that race/ethnicity was missing at random. Not all racial/ethnic groups were reported by each state, and some states reported ethnicity separately from race. The percentage of vaccinations for which combined race/ethnicity was unknown ranged across states from 2% to 35% (median 10%). The percent of vaccinations for which race was unknown, when reported separately from ethnicity, ranged from 2% to 27% (median 10%). The percent of vaccinations for which ethnicity was unknown, when reported separately from race, ranged from 3% to 85% (median 21%). We also found that the shares of vaccinations going to individuals reporting race/ethnicity as “Other” were several times larger than the census population category (which includes multiracial individuals), suggesting that self-reported “Other” in vaccination data does not map well to the census category. As a result, we treated “Other” as “Unknown.” Data on the racial/ethnic distribution of vaccinations were unavailable from North Dakota, Hawaii, Montana, New Hampshire, and Wyoming. For these states, along with specific racial/ethnic groups not included by some states, we used relative uptake rates aggregated at the census division level. Racial/ethnic groups that did not have state-specific data available together comprised 2.0% of the United States population aged 16 years and older.

The age distribution of vaccinations was unavailable from Arkansas, Georgia, Hawaii, Montana, New Hampshire, New York, and Wyoming as of March 31, 2021. For these states, we used the national distribution of vaccinations across age groups reported by the CDC.

### Relative Rates of Uptake

Combining the data on vaccination coverage extracted from state dashboards with demographic data from the American Community Survey, we estimated relative uptake rates by race/ethnicity within each state. We defined relative uptake rates as the observed share of vaccinations for a racial/ethnic group, divided by the expected share under proportionate uptake. The expected share was first computed within specific age groups assuming that vaccinations within an age group were received by persons in different racial/ethnic groups in proportion to population size in that age group. The all-age expected share for each racial/ethnic group was a weighted average of age-specific shares, weighted by the fraction of all vaccinations delivered to that age group. This approach allowed us to control for the interaction of historical age-based eligibility criteria and age-race population structures, thereby isolating the impacts of accessibility of vaccinations (e.g., language, internet access, appointment systems, transportation, time off work) and differences in vaccine confidence by race/ethnicity.

### Scale-Up Scenarios

We modeled scale-up of vaccinations within each census tract in a state under three different scenarios. In all scenarios, we assumed a steady vaccination rate based on state-specific seven-day averages reported by the CDC between March 26-April 1. We assumed that vaccination doses would be distributed to census tracts within each state either in proportion to population size, or prioritized based on the CDC Social Vulnerability Index, depending on the scenario. We assumed that within a census tract, vaccination doses would be allocated across racial ethnic groups in proportion to population size, weighted by the estimated relative uptake rate. The status quo scenario assumed that relative uptake rates would persist at the levels estimated through March 31. Alternative scenarios allowed for convergence of relative uptake rates to reach 1.0 for all race/ethnicity over a six-week period beginning April 1, 2021.

### Analysis

All analyses were conducted using R version 4.0.3. Data and code are available at mirrored repositories through the Prevention Policy Modeling Lab (https://github.com/PPML/covid_vaccination_disparities) and through SC-COSMO (https://github.com/SC-COSMO/covid_vaccination_disparities).

## Results

In most states, relative uptake rates among eligible White populations have been substantially higher than among Black and Hispanic populations, by a median factor of 1.3 times for White compared to Black adults (IQR, 1.2-1.4) and median 1.4 times for White compared to Hispanic adults (IQR, 1.2-1.8) (Figure 1). The joint effects of disproportionate uptake and age-based eligibility focused on older adults has resulted in estimated national coverage among Black and Hispanic adults (27%) being 39% lower than among White adults (44%) as of March 31, 2021.

**Figure 1.**
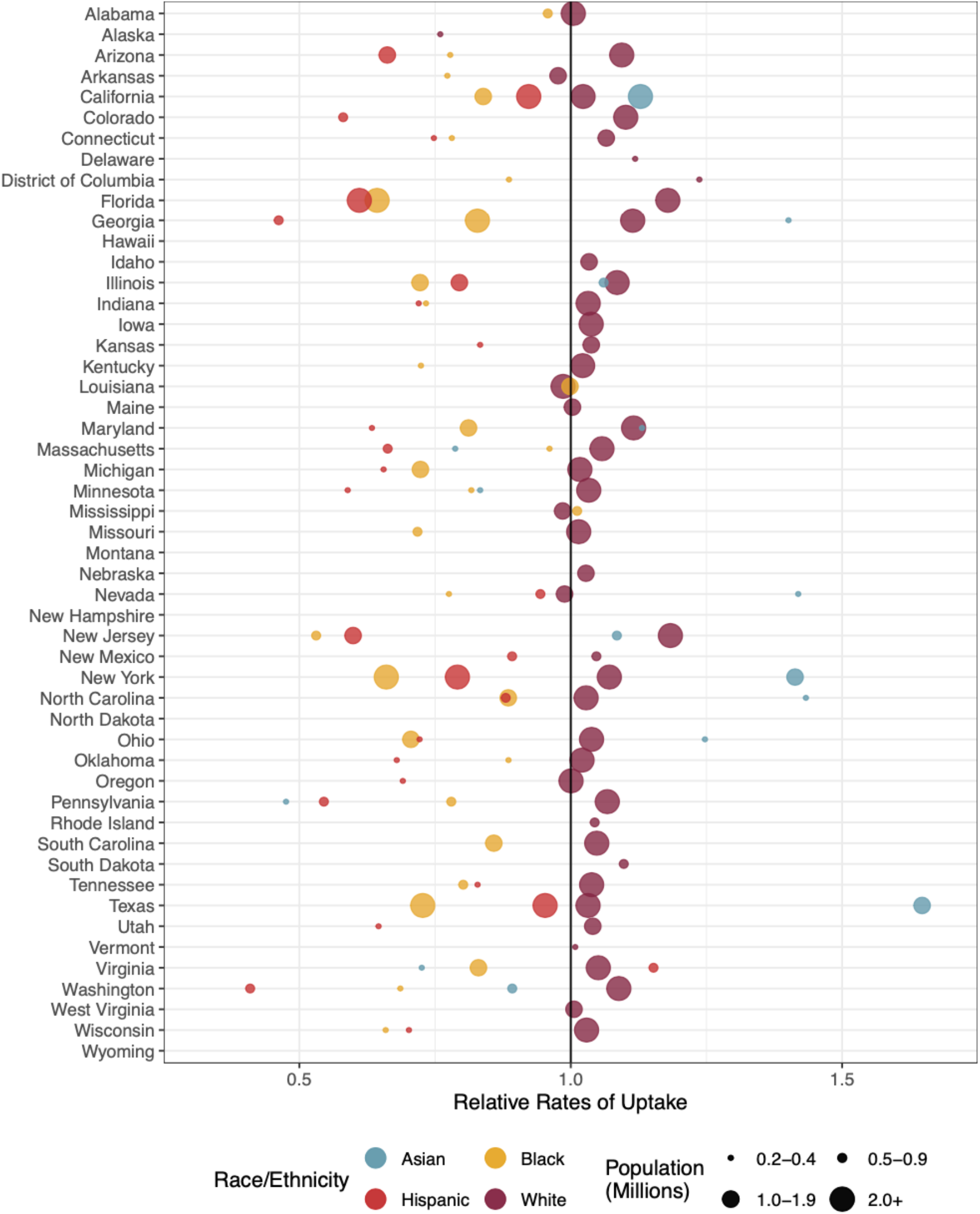
Relative rates of COVID-19 vaccination uptake, by race/ethnicity and state. Estimates shown for populations that exceed 200,000 and have data available on state reporting dashboards.

If current disparities in uptake rates persist among the eligible adult population (‘*Status quo*’ scenario), Hispanic adults and Black adults would reach 75% coverage of ≥1 vaccine dose nationally 23 days and 30 days later, respectively, than White adults (Figure 2, state-specific results in Supplement). When White adults reach 75% coverage nationally, coverage among Hispanic and Black adults would be 59%.

**Figure 2.**
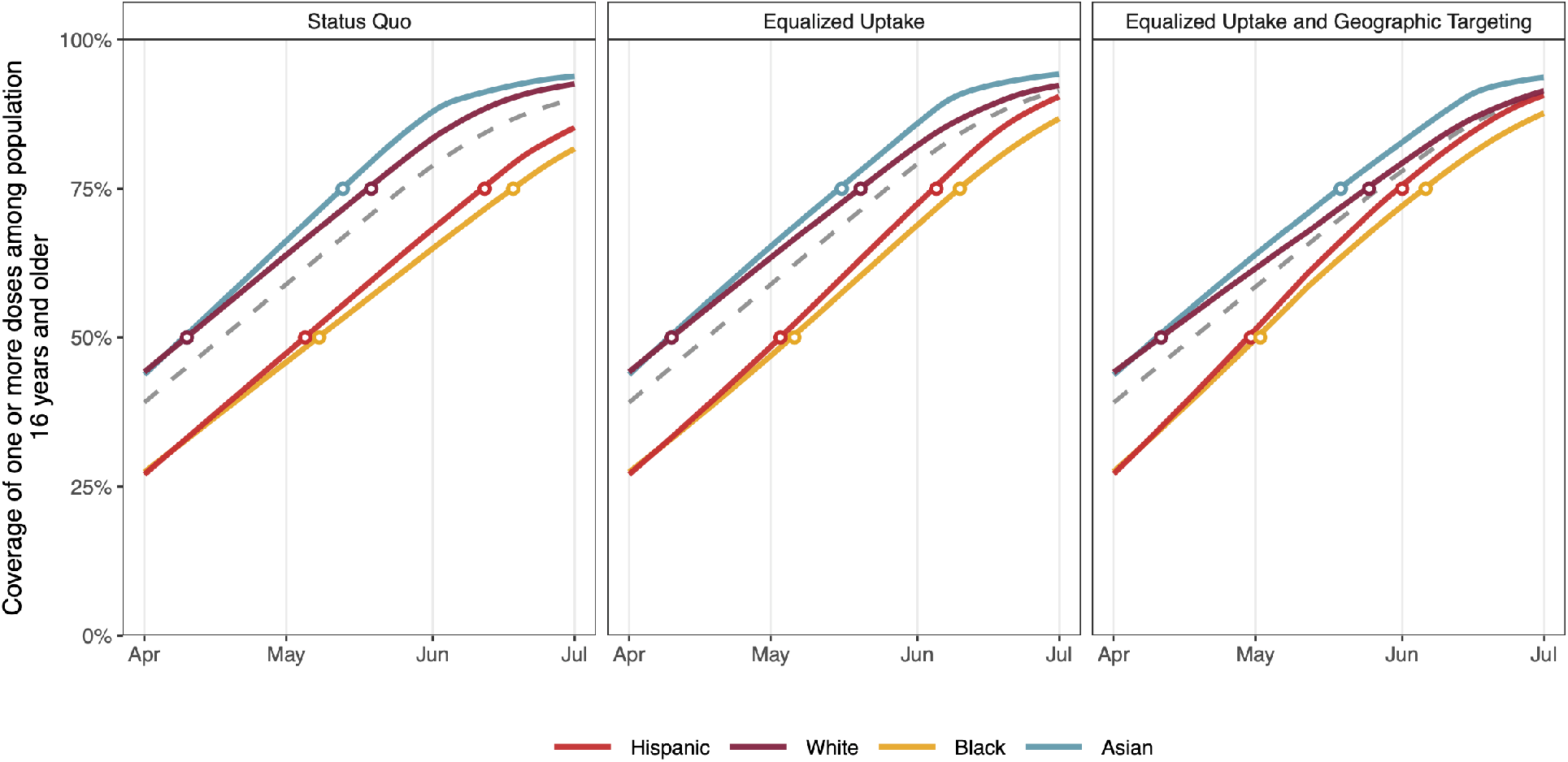
Coverage of one or more COVID-19 vaccination doses among population 16 years and older, by racial/ethnic group, aggregated to national level. Panels show scenarios: A) status quo, B) equalized uptake, and C) equalized uptake and geographic targeting. Dashed line shows overall coverage among the US population aged 16 years and older.

Analyzing status quo scale-up by state, the pace of vaccination has varied dramatically across the country. Projecting the vaccination rate on April 1^st^ forward, New Hampshire, New Mexico, Connecticut, South Dakota, Massachusetts, District of Columbia, and Virginia are on pace to reach 75% coverage of ≥1 vaccine dose among their adult population more than two weeks earlier than the national average. On the other hand, Mississippi, Missouri, Alabama, Alaska, Wyoming, Indiana, and Louisiana are projected to reach the 75% threshold more than three weeks later than the national average. Mississippi, with the slowest vaccination rate, is projected to reach the 75% threshold more than two months later (Figure 3).

**Figure 3.**
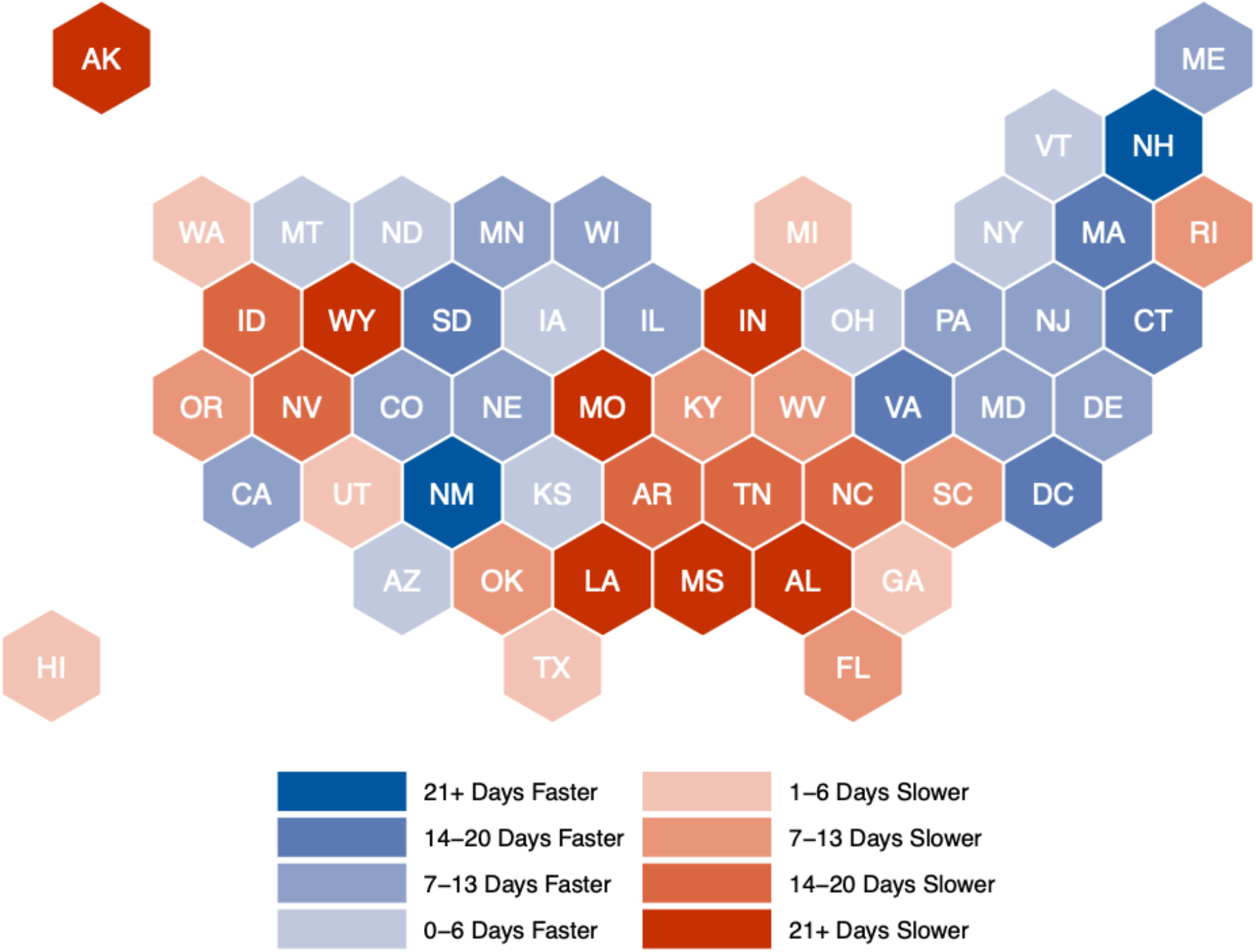
Time difference in reaching 75% coverage of ≥1 vaccine dose among adults ages 16+, compared to the national average, if differences in rates of uptake between states persist.

Within states, disparities in vaccination rates between White populations and racial/ethnic minorities are widespread. People of color in 11 states are projected to have a delay in reaching the 75% coverage threshold of longer than 35 days, compared to White adults in the same state. Five states are projected to have a delay of 28-34 days, 14 states are projected to have a delay of 21-27 days, seven states are projected to have a delay of 14-20 days, nine states are projected to have a delay of 7-13 days, and three states are projected to have a delay of 1-6 days. Only people of color in Alaska, Hawaii, and Washington, are projected to reach the 75% coverage threshold before White adults (Figure 4).

**Figure 4.**
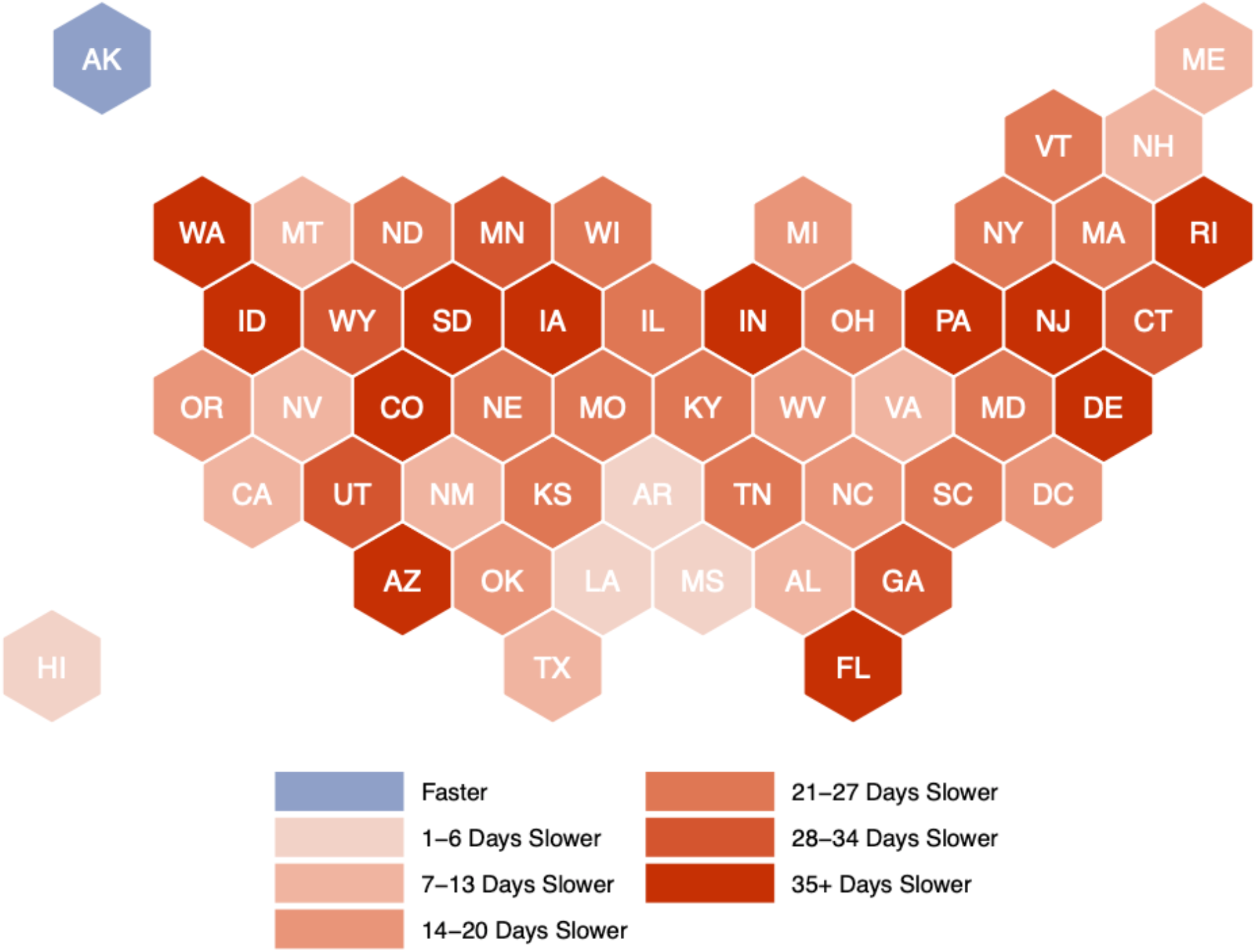
Delay in reaching 75% coverage of ≥1 vaccine dose among adults ages 16+ for people of color, compared to White people, by state if historical disparities in relative uptake rates persist.

If relative uptake rates across racial/ethnic groups trended steadily from starting values (Figure 1) toward 1.0 over six weeks (‘*Equalized uptake*’ scenario), delays in reaching 75% thresholds would shrink to 15 and 20 days for Hispanic and Black adults, respectively (Figure 2 & Supplement). Geographic targeting of vaccination doses to disadvantaged communities could further reduce disparities. For example, doubling per-capita dose allocations for the most disadvantaged quartiles of census tracts in each state, as measured by the CDC Social Vulnerability Index, further reduces delays to reach 75% thresholds to 6 days for Hispanic adults and 12 days for Black adults, and could narrow national coverage disparities on July 1, 2021 by 92% for Hispanic adults and 66% for Black adults (‘*Equalized uptake and geographic targeting*’ scenario in Figure 2 & Supplement).

## Discussion

Interventions are urgently needed to eliminate disparities in COVID-19 vaccination coverage. In all but three states, White populations are projected to reach 75% coverage among adults faster than non-White populations. Eliminating access barriers and increasing vaccine confidence among marginalized populations can narrow, but not eliminate, gaps in coverage. Combining these interventions with place-based allocation strategies can further accelerate vaccination in disadvantaged communities, which have borne a disproportionate burden from COVID-19.

Previous efforts have highlighted that Black and Hispanic populations have been vaccinated at a lower rate compared to White populations (4). One contribution of our analysis is to derive relative vaccination uptake rates across racial/ethnic groups from vaccination data reported by states, that adjust for differential eligibility relating to demographic differences, in order to capture the combined effects of disparities in vaccination accessibility and vaccine confidence. After controlling for the impact of age-based eligibility criteria on the expected distribution of vaccinations by race, we observe persistently lower in relative uptake rates among Black and Hispanic populations across states. Although age-based eligibility criteria contributed to lower coverage among non-White populations, expanding eligibility to all adults will not resolve coverage disparities and threatens to exacerbate disparities for a period of time due to increased competition for appointments.

We find that interventions to equalize uptake rates across racial/ethnic groups could narrow gaps in coverage, but that disparities produced by unequal update over the first four months of the vaccine campaign are sufficiently large that even equal uptake rates would not close these gaps over the next four months. On the other hand, geographic targeting of vaccine doses to disadvantaged communities has potential to accelerate progress toward a more equitable convergence of coverage by July. Previous studies have suggested that disadvantage indices should be used to prioritize allocation (5,6). Beyond their role in improving the equity of the nation’s vaccination campaign, place-based allocation strategies are also likely more effective overall, since individuals living in disadvantaged areas, predominantly low-income individuals and people of color, have borne a disproportionate burden of COVID-19 cases, hospitalizations, and deaths (7).

The findings of our study should be interpreted in the context of its limitations. First, our relative uptake rates were estimated based on a cross-sectional snapshot of vaccination data. Future work should monitor trends in uptake rates and utilize these to inform projections. Second, projection scenarios are based on a simplifying assumption that state-level vaccination rates would remain constant at the seven-day average levels observed on April 1. Although supply is likely to fluctuate in actuality, within-state disparities between racial/ethnic groups are unlikely to be markedly different under alternative supply assumptions. Third, a substantial proportion of vaccinations are attributed to individuals of either “Other” or “Unknown” race/ethnicity. Despite these limitations, our study provides important information on disparities in vaccination, the persistence of inequitable risks if unequal uptake continues, and key elements of vaccination campaigns for states to consider in order to address these inequities.

States should work to achieve equitable vaccination coverage through interventions that act on both vaccine supply and demand. Multiple states have implemented place-based allocation schemes (4). Actions are also needed to eliminate transportation and language barriers, minimize unfair competition for appointments (e.g. by adopting pre-registration systems), increase vaccine confidence among marginalized populations, and accommodate work schedules and time off for vaccination. As the country races toward coverage goals required to control the epidemic, pro-equity policies are critical to ensuring that underserved communities are not left behind (8).

## Data Availability

Data and code are available at mirrored repositories through the Prevention Policy Modeling Lab (https://github.com/PPML/covid_vaccination_disparities) and through SC-COSMO (https://github.com/SC-COSMO/covid_vaccination_disparities).

