## Supplement for "Addressing racial/ethnic disparities in the COVID-19 vaccination campaign"

Supplemental Appendix

### Technical Notes

1. Vaccination data for Hispanic adults in Alabama were treated as missing because ethnicity is missing for more than half of data. Data for Georgia were from March 22, 2021, as data on race/ethnicity stopped being reported due to a technical issue. Data for Indiana and Maine did not include individuals receiving the Janssen vaccine because the reporting structure for these states did not permit combining individuals receiving first-dose of a two-dose vaccine and individuals receiving the single-dose vaccine. Data for North Dakota were not included because the reporting structure does not permit computation of vaccine distribution by race/ethnicity. Hawaii, Montana, New Hampshire, and Wyoming did not report vaccine distribution by race/ethnicity as of March 31, 2021.
2. The age distribution of vaccinations was unavailable from Arkansas, Georgia, Hawaii, Montana, New Hampshire, New York, and Wyoming as of March 31, 2021. We used the national distribution of vaccinations across age groups for these states.
3. When state-specific data to permit estimation of relative uptake rates by race/ethnicity were unavailable, we used relative uptake rates estimated at the census division level. Racial/ethnic groups that did not have state-specific data available together comprised 2.0% of the United States population aged 16 years and older.
4. State-specific figures in the Supplement only visualize data for racial/ethnic groups with at least 200,000 individuals aged 16 years or older.

### Supplemental Figures: State-Specific Scale-Up Scenarios

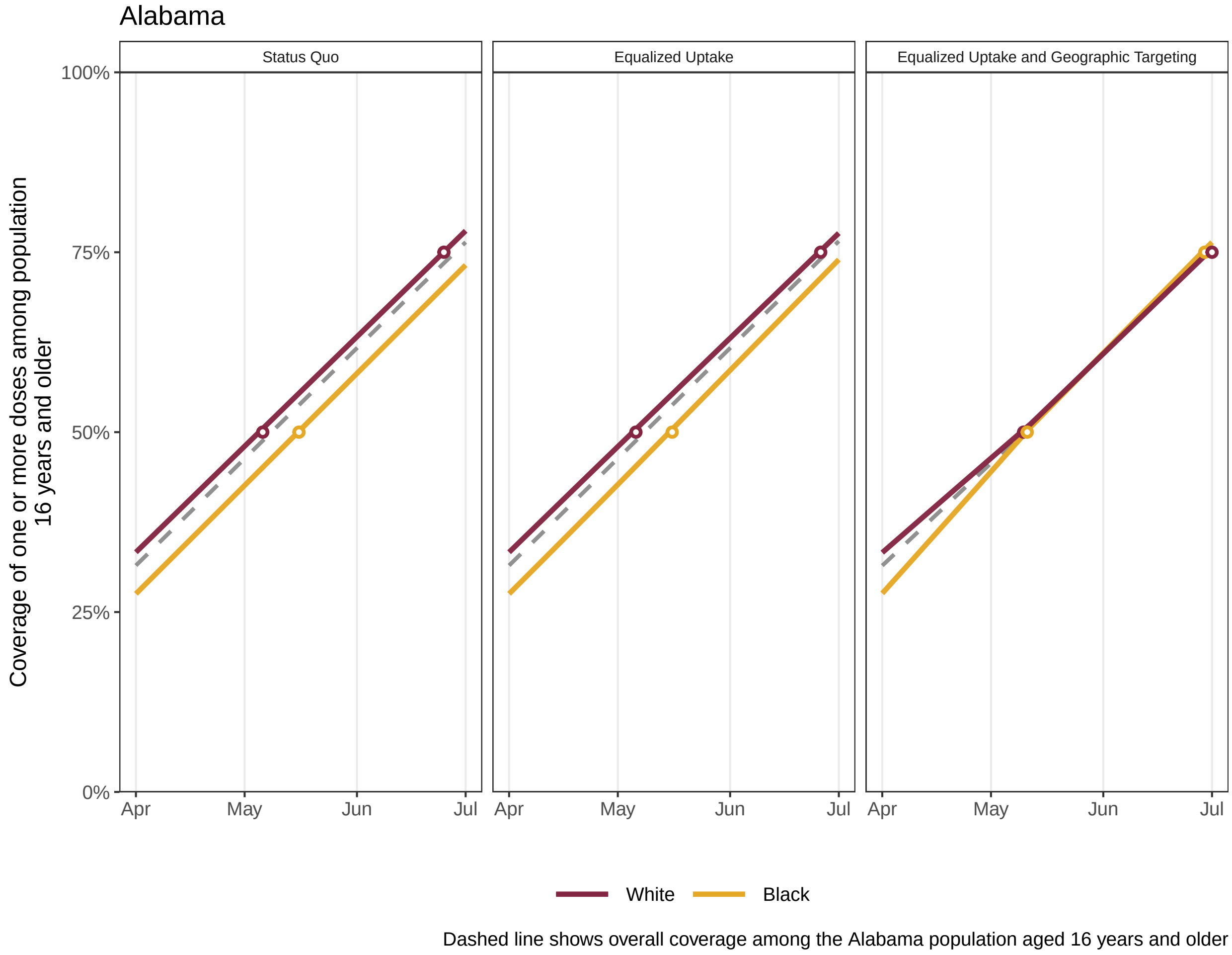

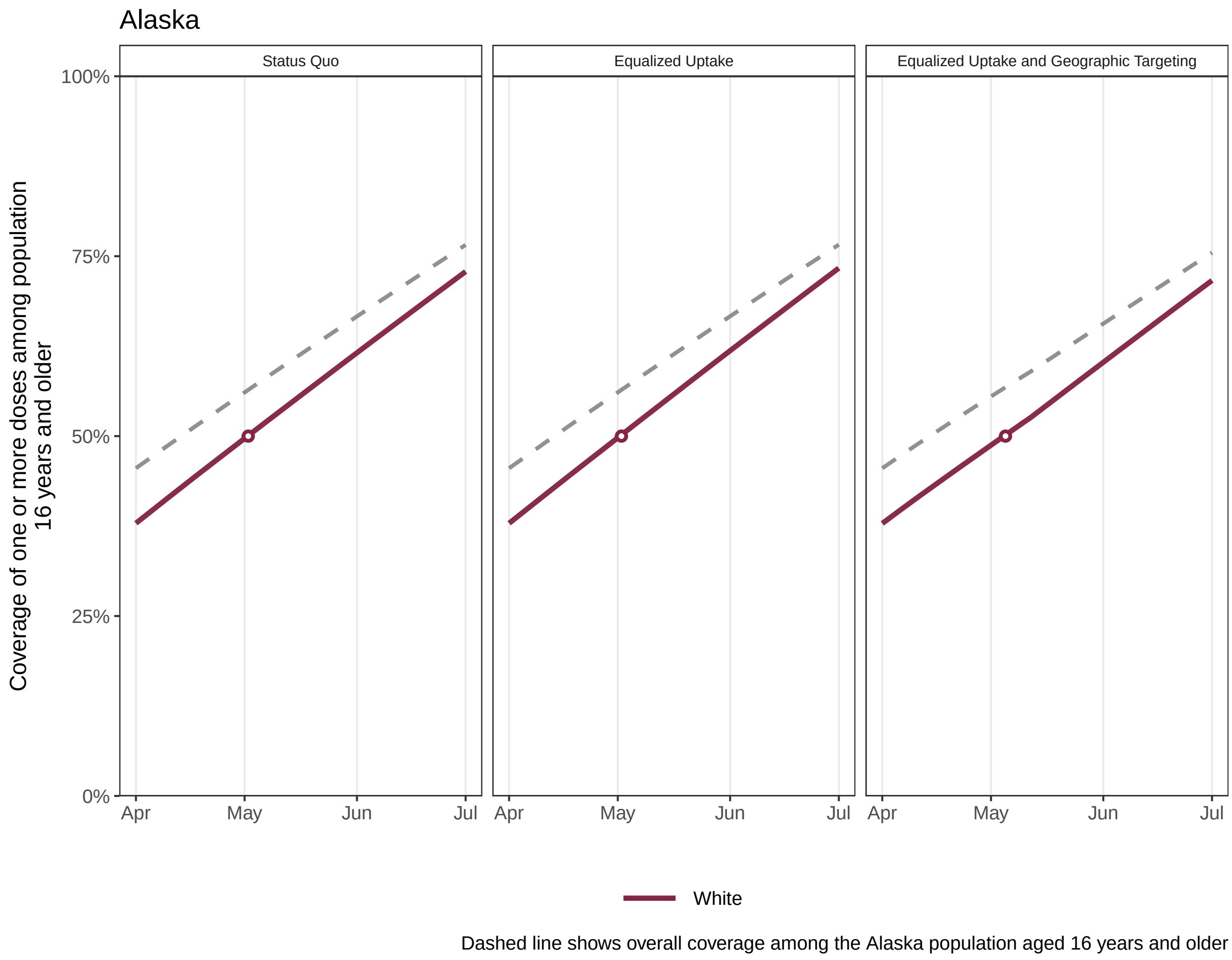
Alaska
Status Quo
Equalized Uptake
Equalized Uptake and Geographic Targeting
Coverage of one or more doses among population 16 years and older
100%
75%
50%
25%
0%
Apr
May
Jun
Jul
White
Dashed line shows overall coverage among the Alaska population aged 16 years and older

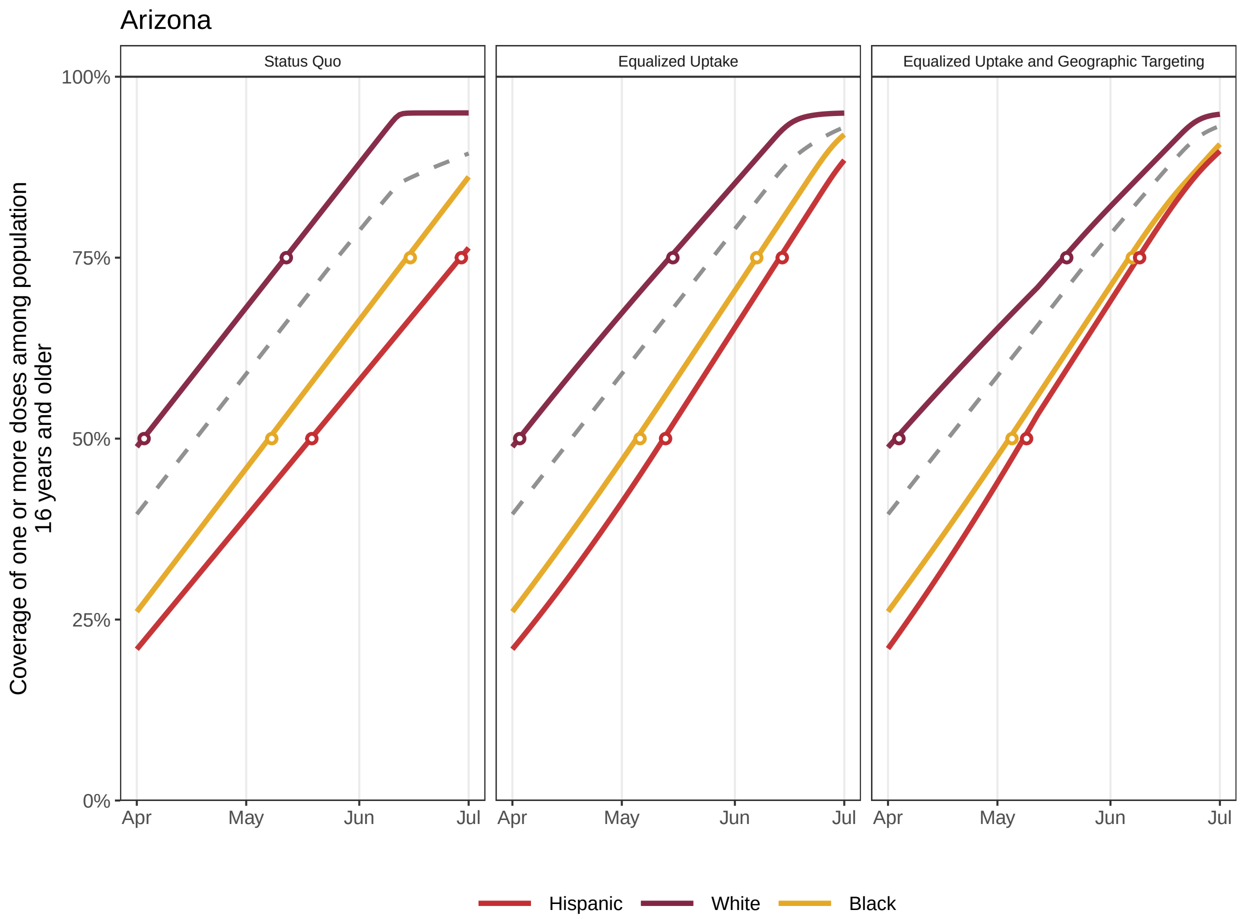


Dashed line shows overall coverage among the Arizona population aged 16 years and older

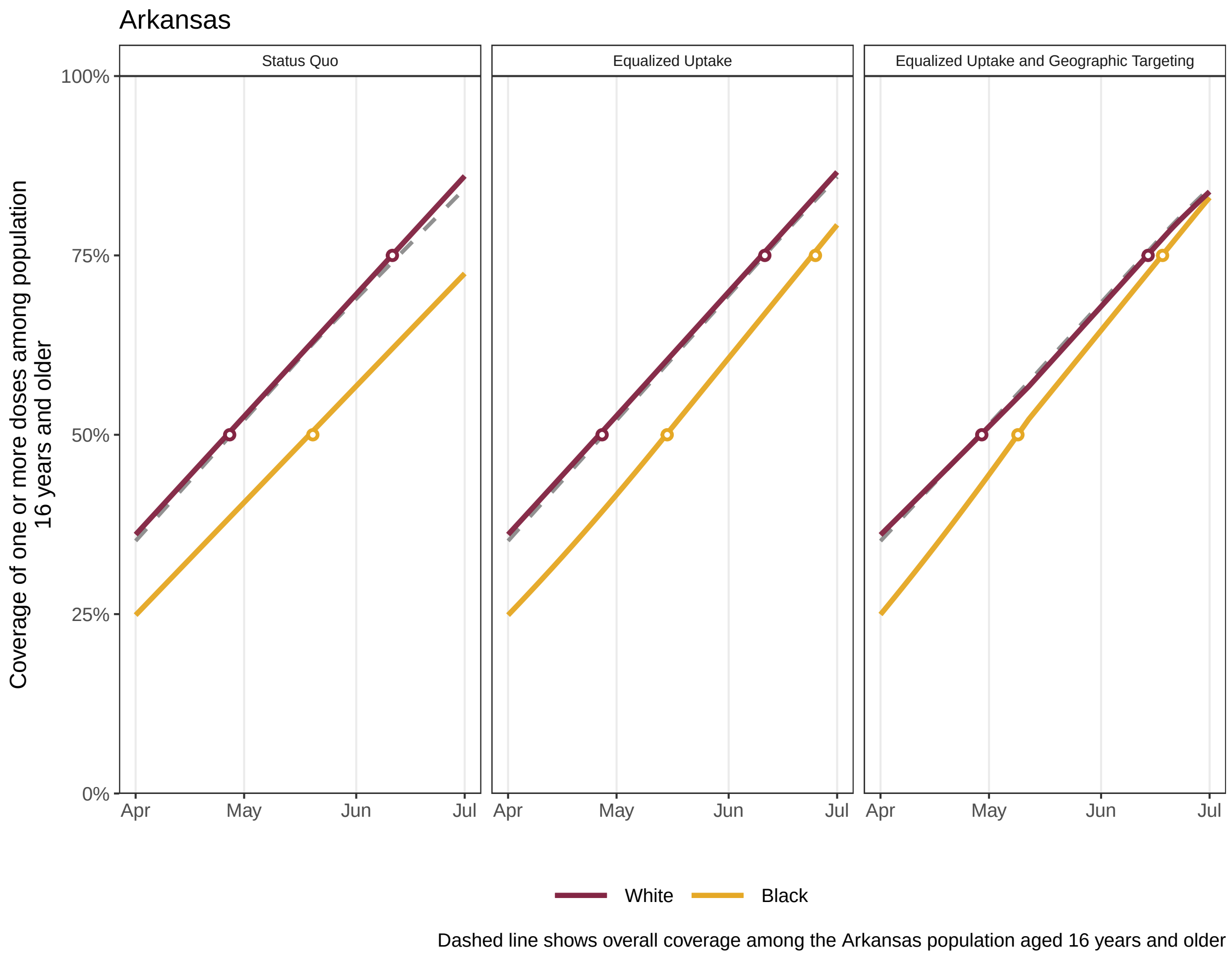
Arkansas
Status Quo
Equalized Uptake
Equalized Uptake and Geographic Targeting
Coverage of one or more doses among population 16 years and older
100%
75%
50%
25%
0%
Apr
May
Jun
Jul
White
Black
Dashed line shows overall coverage among the Arkansas population aged 16 years and older

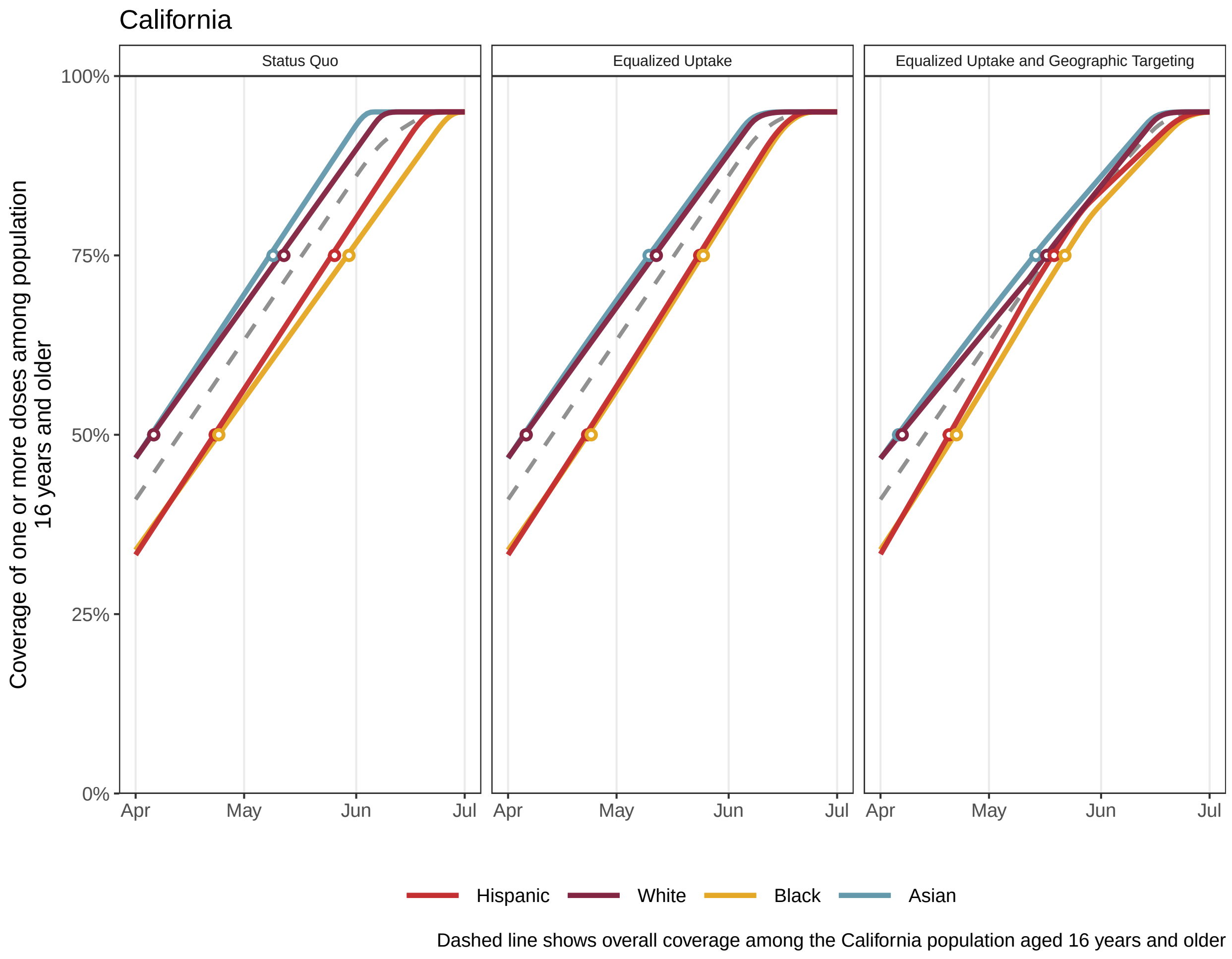
California
Status Quo
Equalized Uptake
Equalized Uptake and Geographic Targeting
Coverage of one or more doses among population
16 years and older
100%
75%
50%
25%
0%
Apr
May
Jun
Jul
Hispanic
White
Black
Asian
Dashed line shows overall coverage among the California population aged 16 years and older

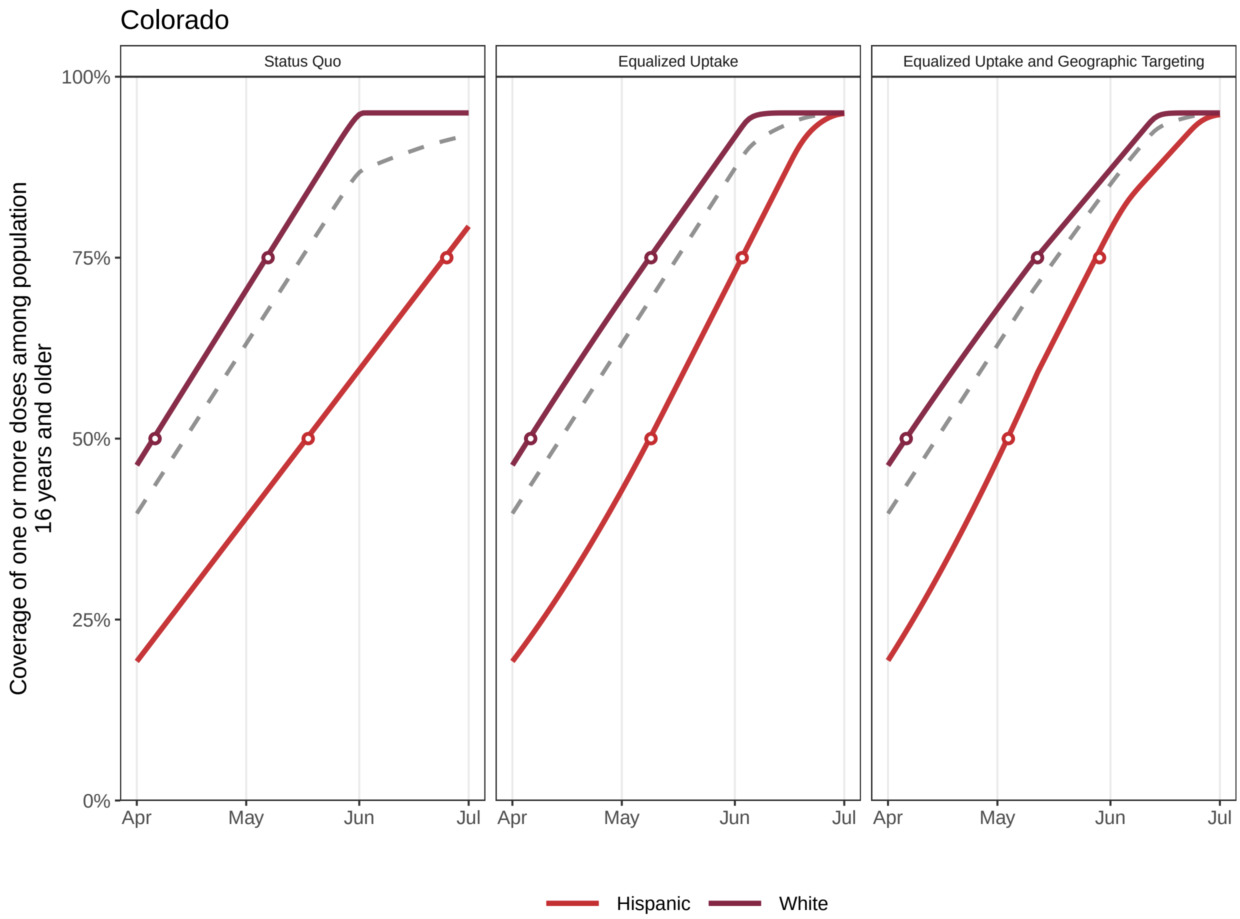


Dashed line shows overall coverage among the Colorado population aged 16 years and older

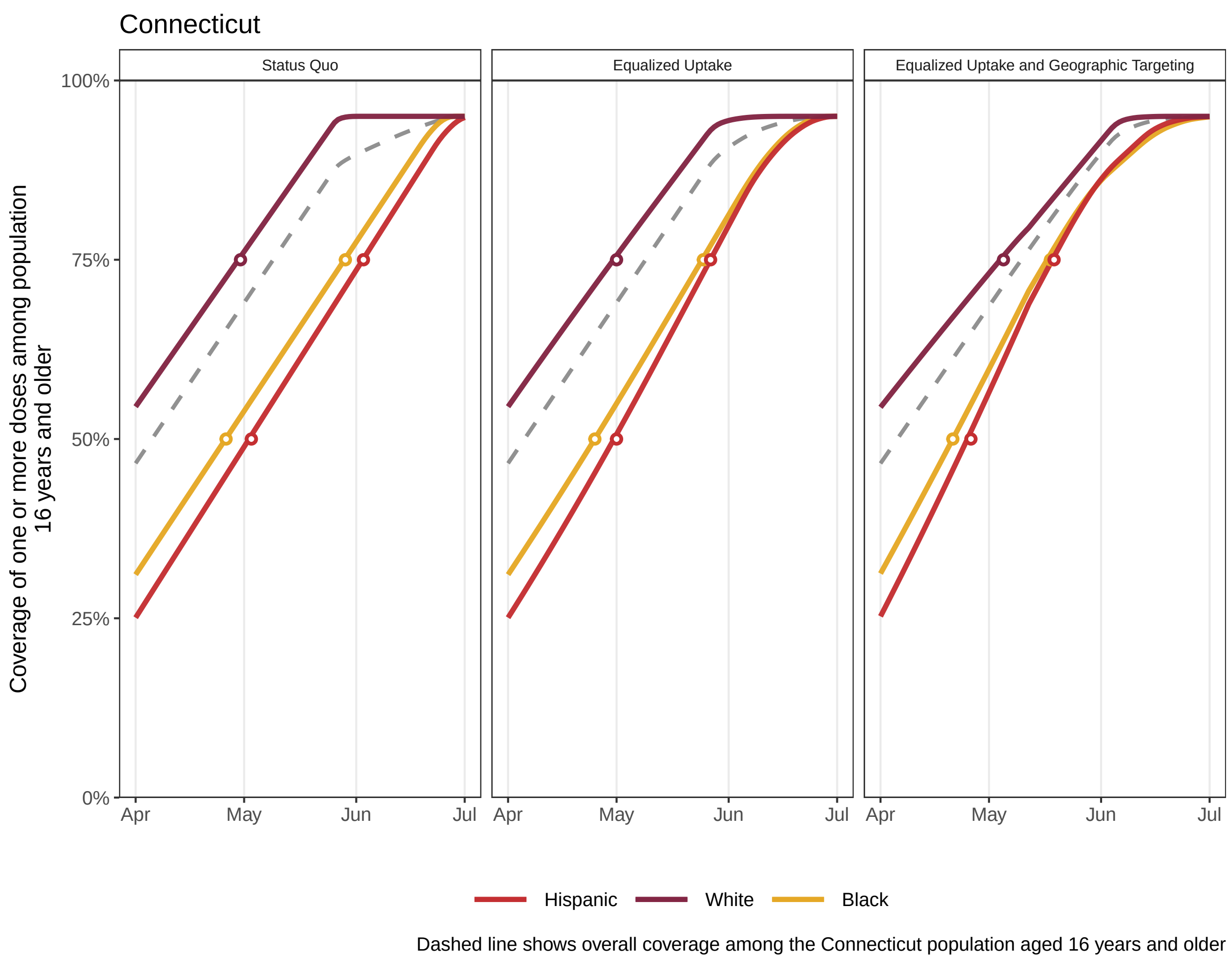
Connecticut
Status Quo
Equalized Uptake
Equalized Uptake and Geographic Targeting
100%
75%
50%
25%
0%
Coverage of one or more doses among population 16 years and older
Apr
May
Jun
Jul
Hispanic
White
Black
Dashed line shows overall coverage among the Connecticut population aged 16 years and older

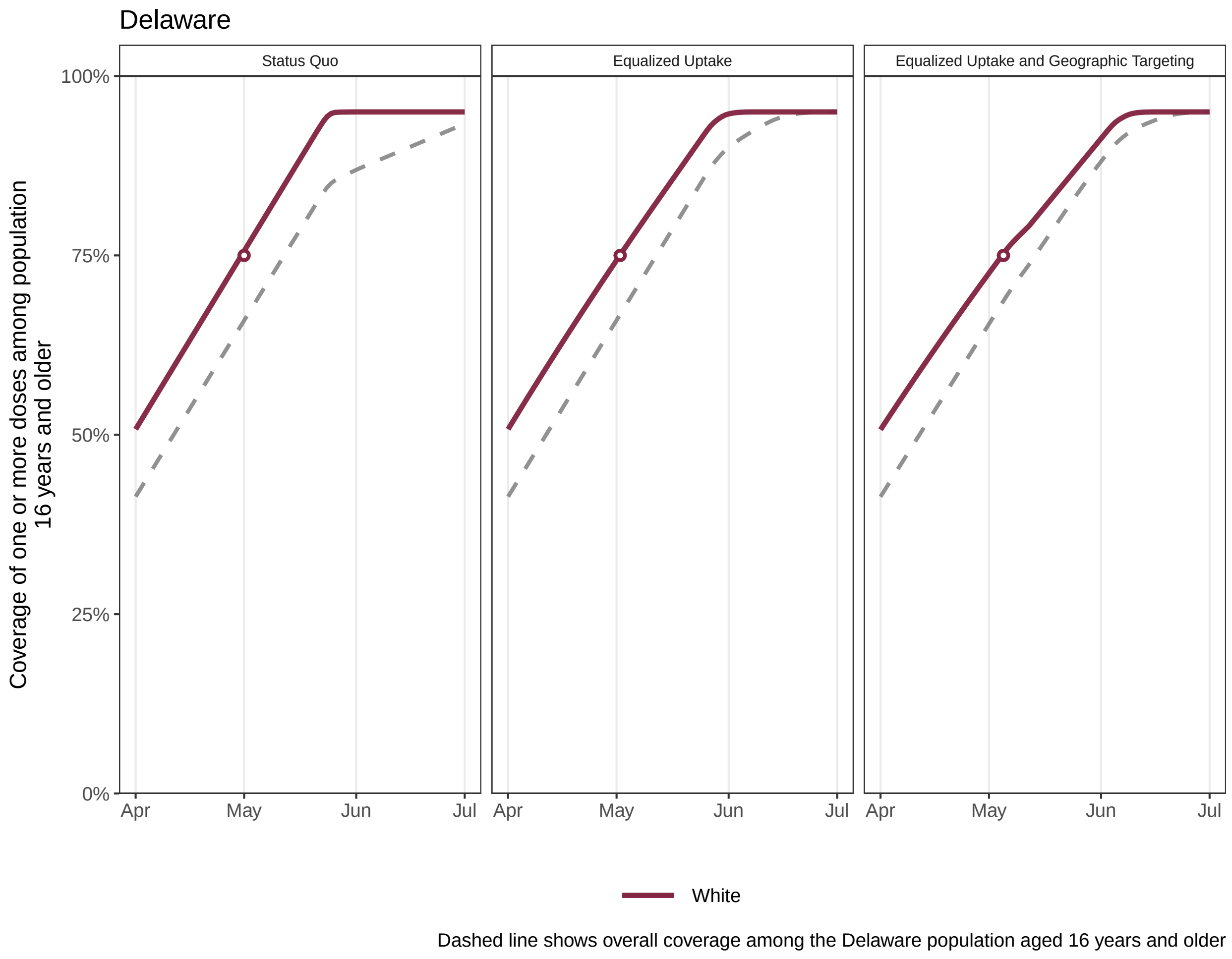

Delaware
Status Quo
Equalized Uptake
Equalized Uptake and Geographic Targeting
100%
75%
50%
25%
0%
Coverage of one or more doses among population 16 years and older
Apr
May
Jun
Jul
White
Dashed line shows overall coverage among the Delaware population aged 16 years and older

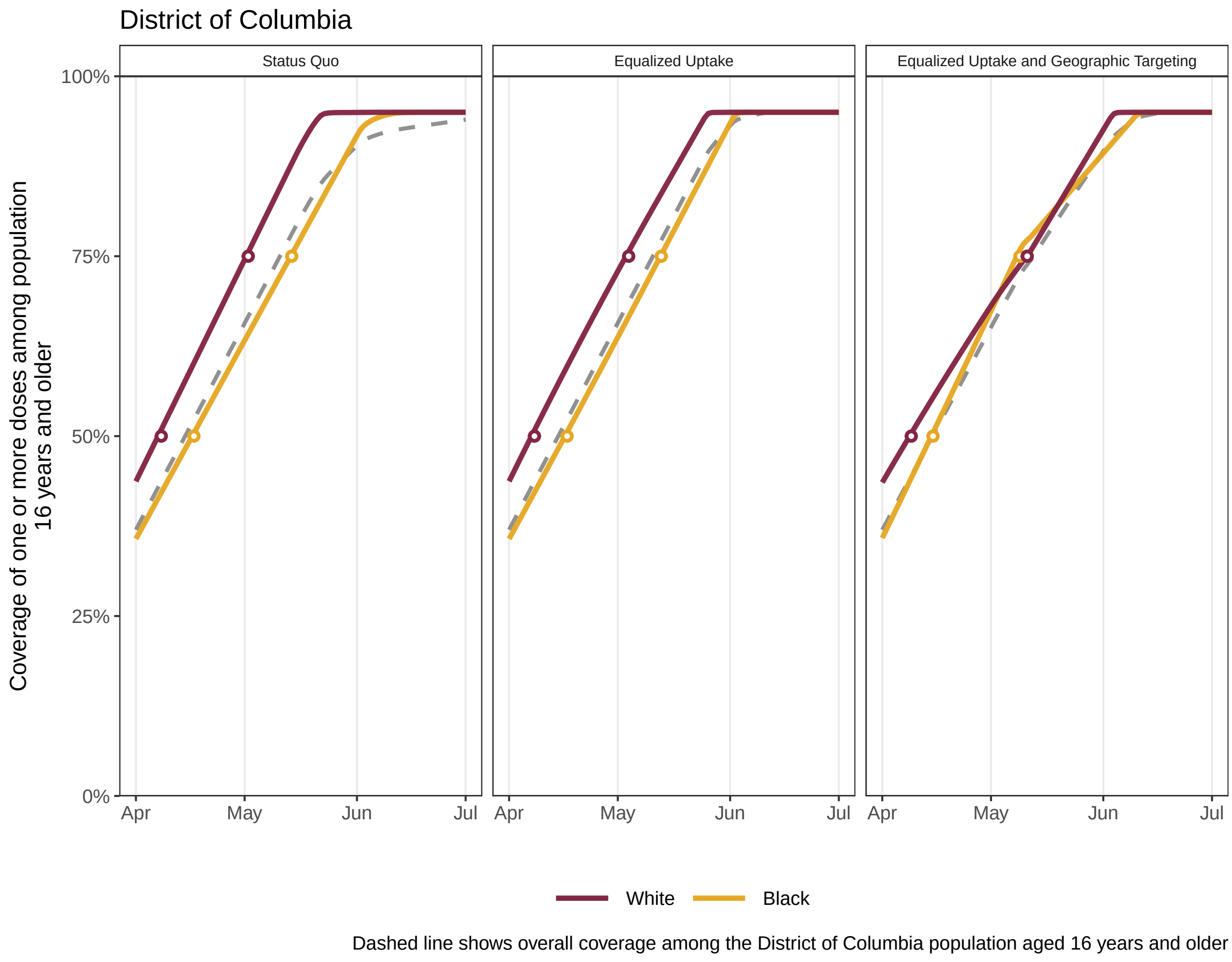
District of Columbia
Status Quo
Equalized Uptake
Equalized Uptake and Geographic Targeting
Coverage of one or more doses among population 16 years and older
100%
75%
50%
25%
0%
Apr
May
Jun
Jul
White
Black
Dashed line shows overall coverage among the District of Columbia population aged 16 years and older

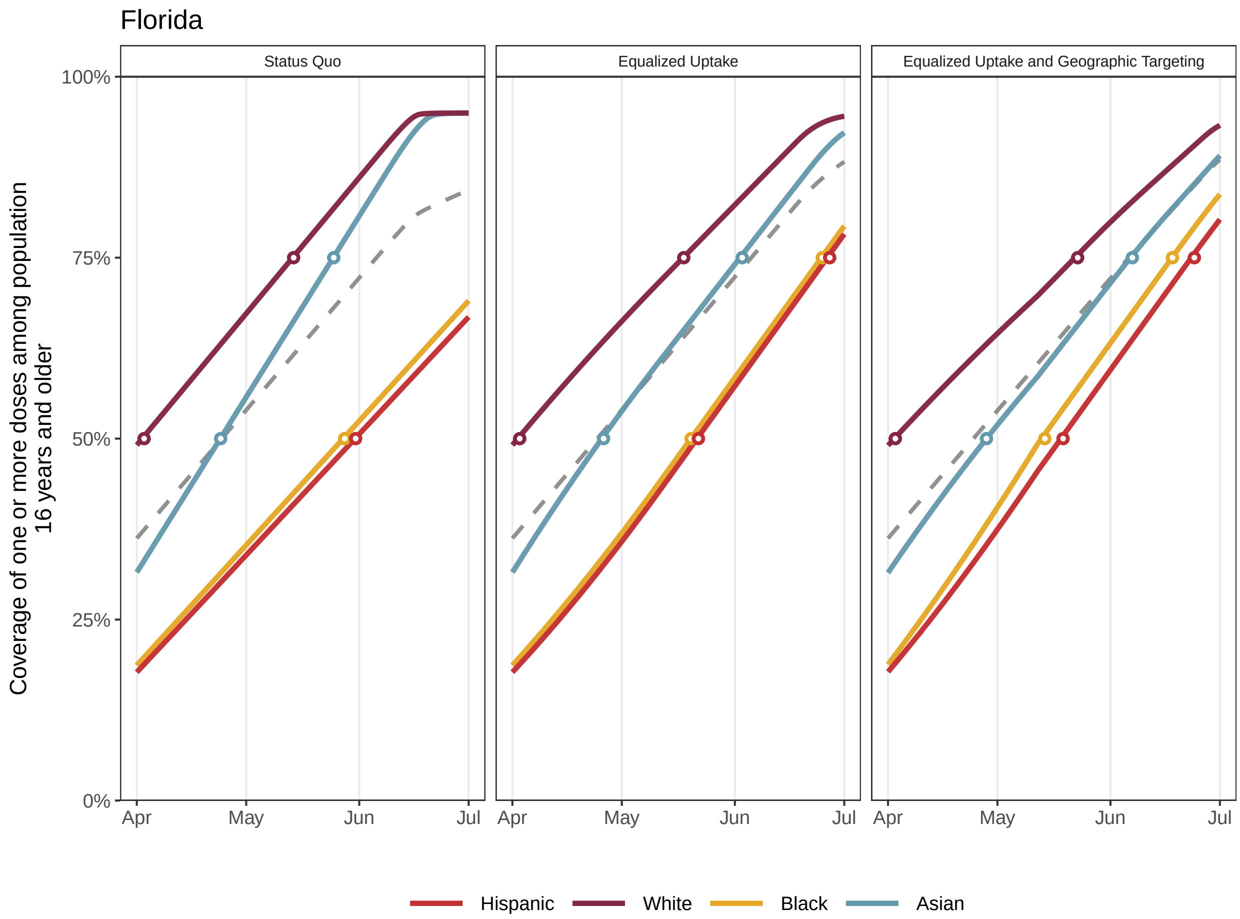


Dashed line shows overall coverage among the Florida population aged 16 years and older

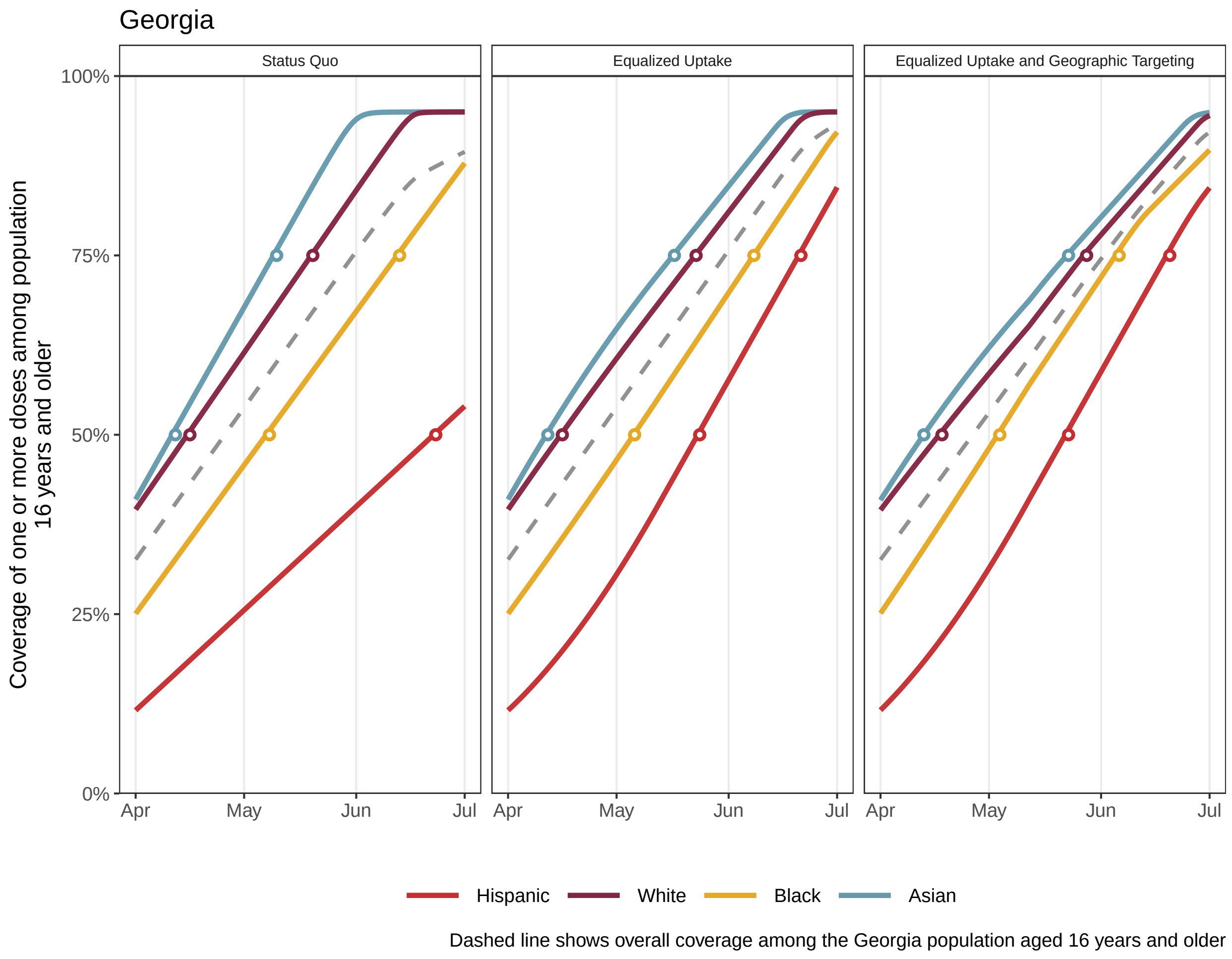
Georgia
Status Quo
Equalized Uptake
Equalized Uptake and Geographic Targeting
Coverage of one or more doses among population 16 years and older
100%
75%
50%
25%
0%
Apr
May
Jun
Jul
Hispanic
White
Black
Asian
Dashed line shows overall coverage among the Georgia population aged 16 years and older

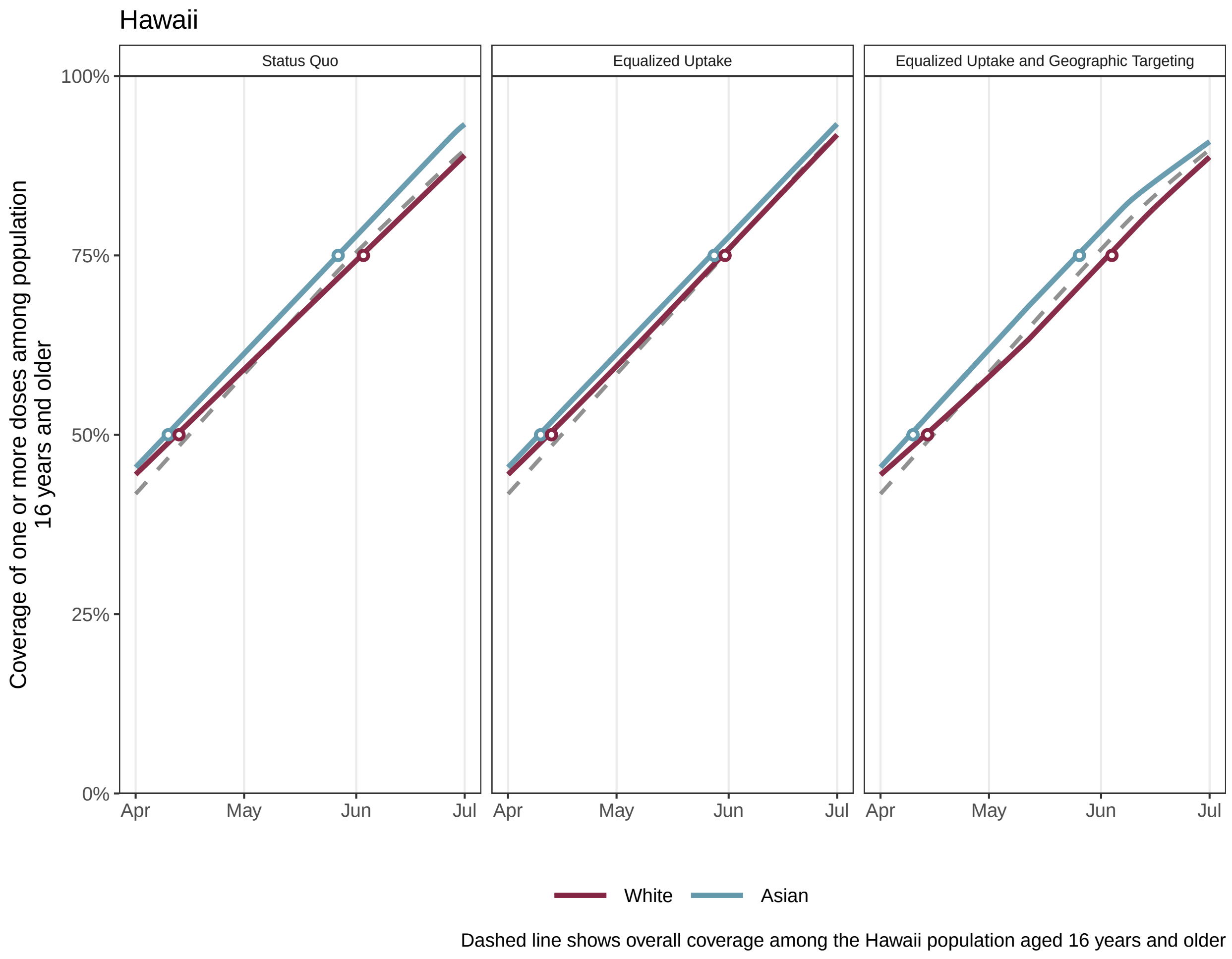
Hawaii
Status Quo
Equalized Uptake
Equalized Uptake and Geographic Targeting
Coverage of one or more doses among population 16 years and older
100%
75%
50%
25%
0%
Apr
May
Jun
Jul
White
Asian
Dashed line shows overall coverage among the Hawaii population aged 16 years and older

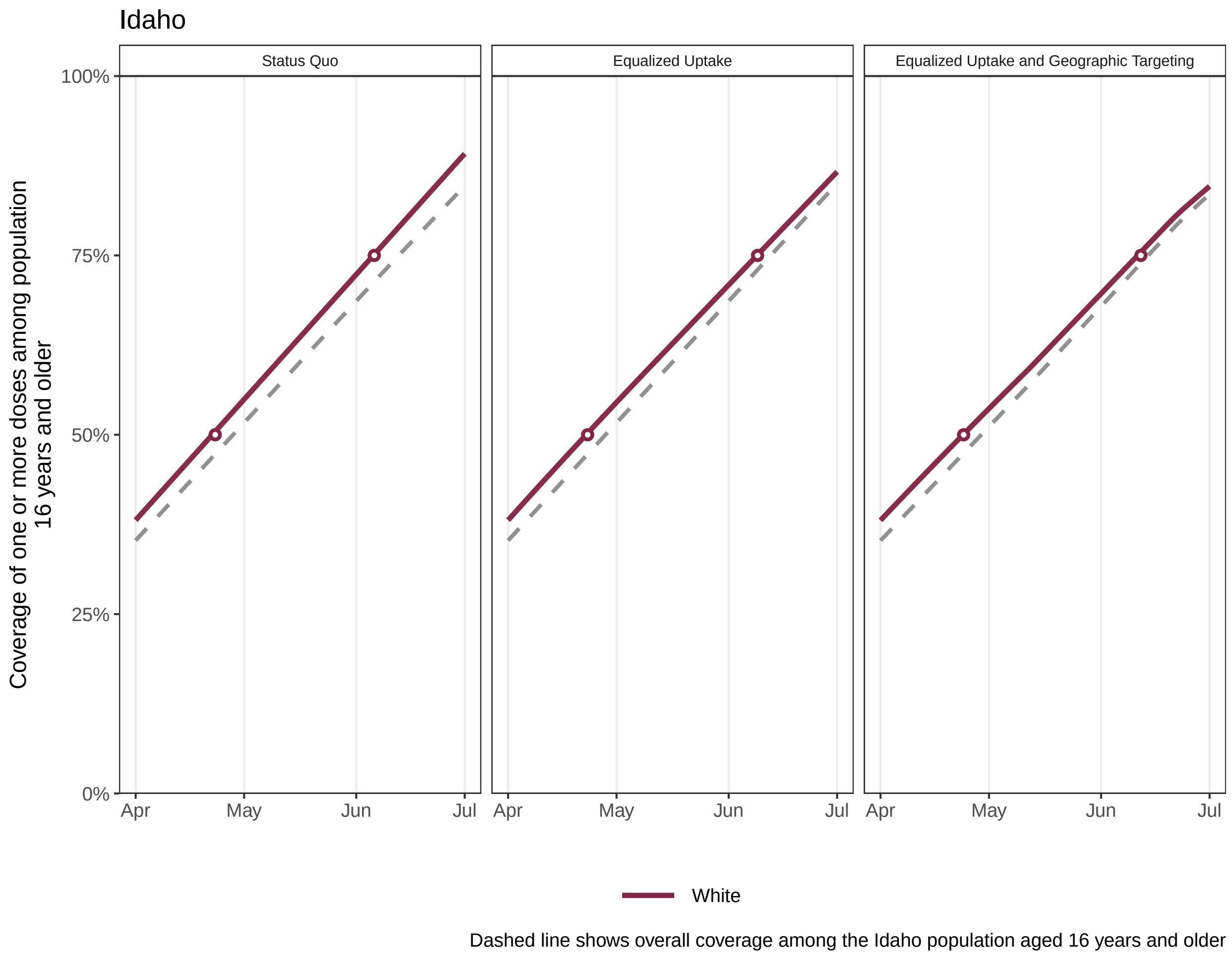
Idaho
Status Quo
Equalized Uptake
Equalized Uptake and Geographic Targeting
Coverage of one or more doses among population 16 years and older
100%
75%
50%
25%
0%
Apr
May
Jun
Jul
White
Dashed line shows overall coverage among the Idaho population aged 16 years and older

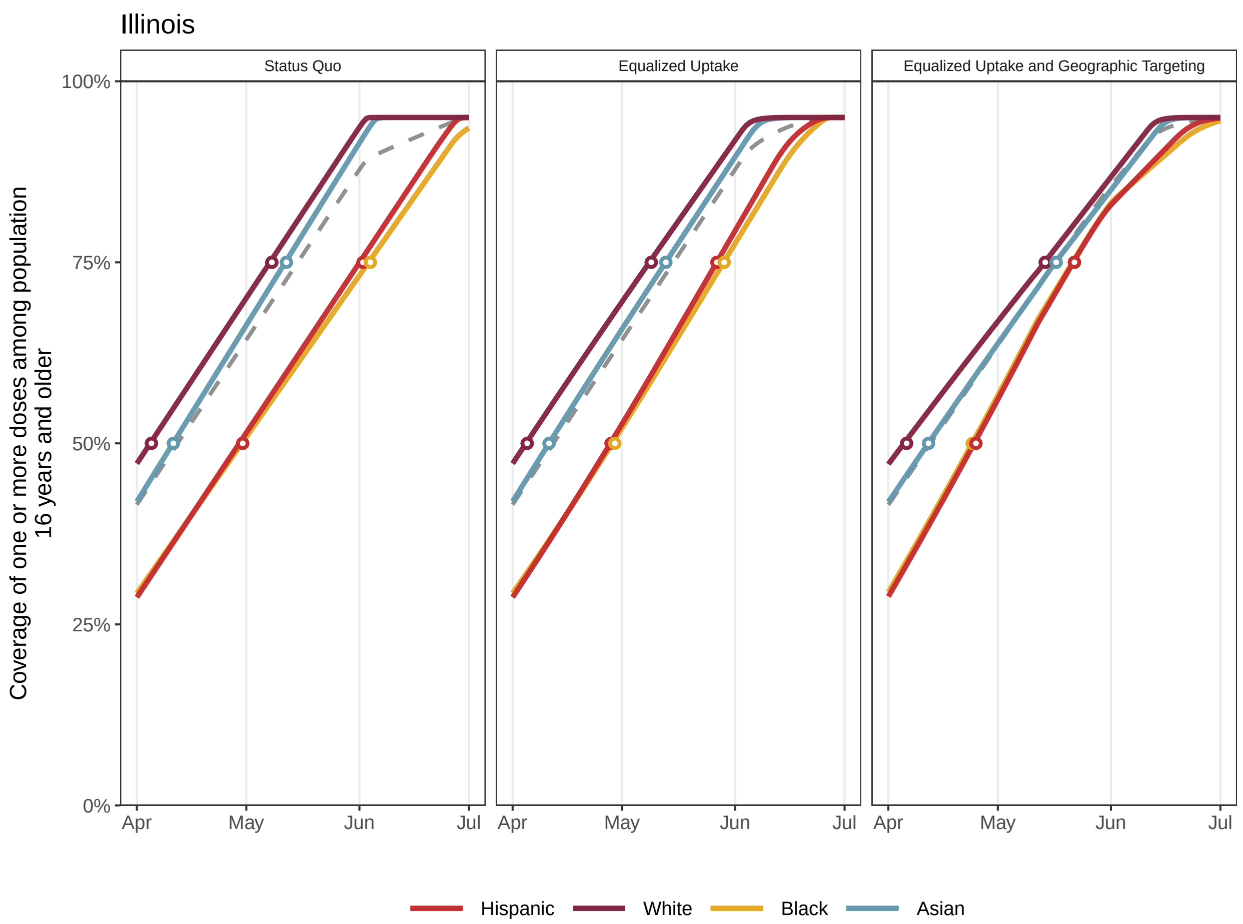


Dashed line shows overall coverage among the Illinois population aged 16 years and older

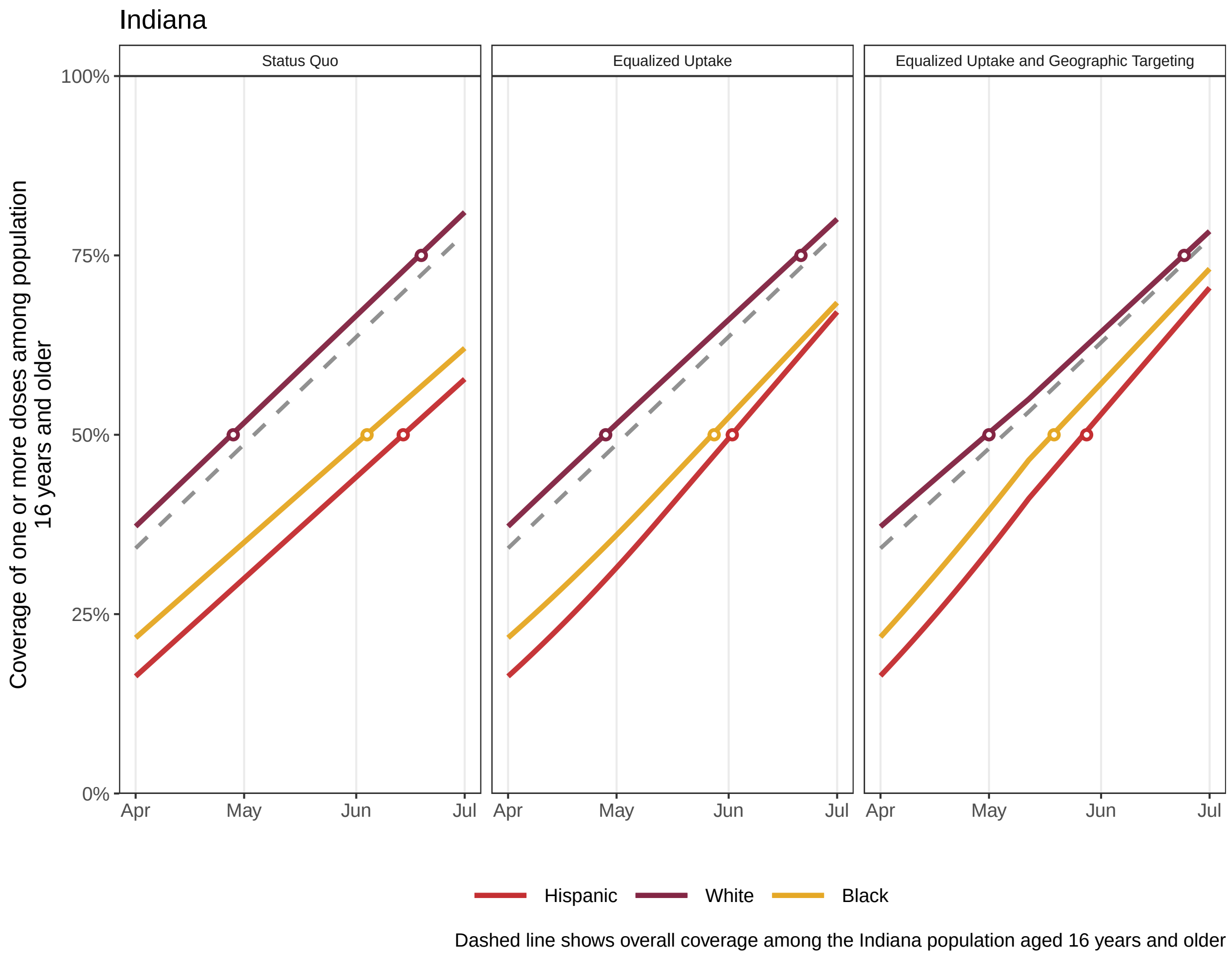
Indiana
Status Quo
Equalized Uptake
Equalized Uptake and Geographic Targeting
Coverage of one or more doses among population 16 years and older
100%
75%
50%
25%
0%
Apr
May
Jun
Jul
Hispanic
White
Black
Dashed line shows overall coverage among the Indiana population aged 16 years and older

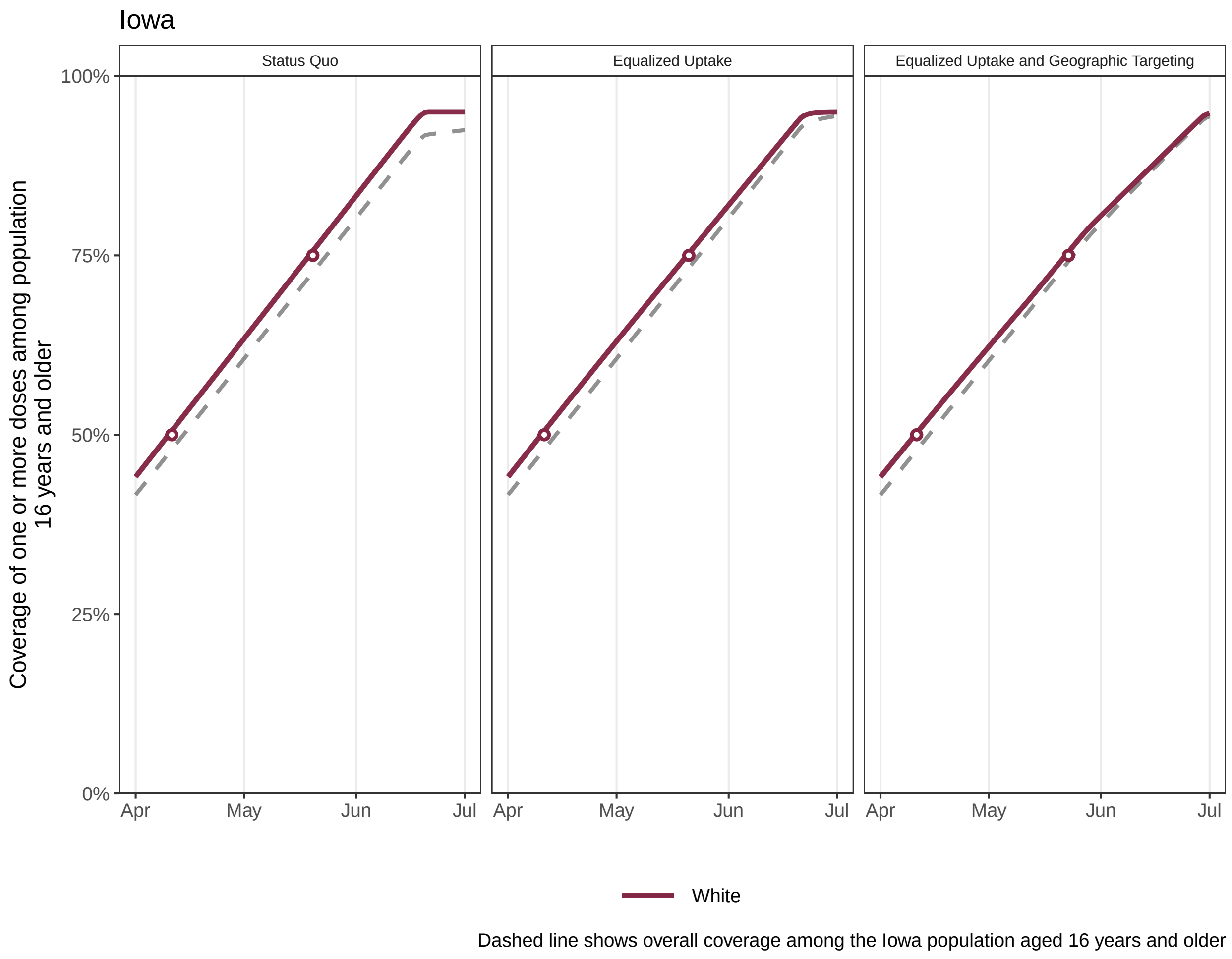

Iowa
Status Quo
Equalized Uptake
Equalized Uptake and Geographic Targeting
Coverage of one or more doses among population 16 years and older
100%
75%
50%
25%
0%
Apr
May
Jun
Jul
White
Dashed line shows overall coverage among the Iowa population aged 16 years and older

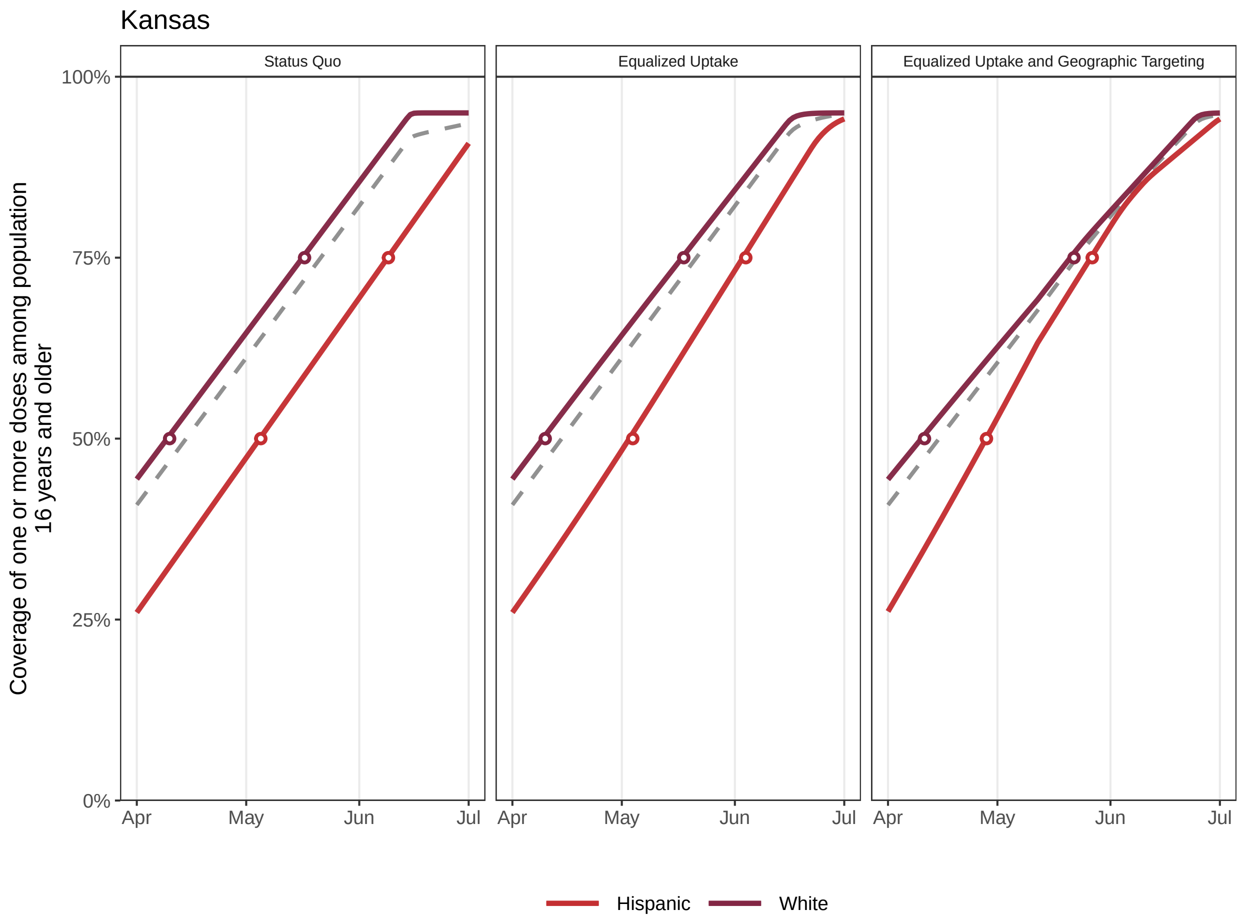


Dashed line shows overall coverage among the Kansas population aged 16 years and older

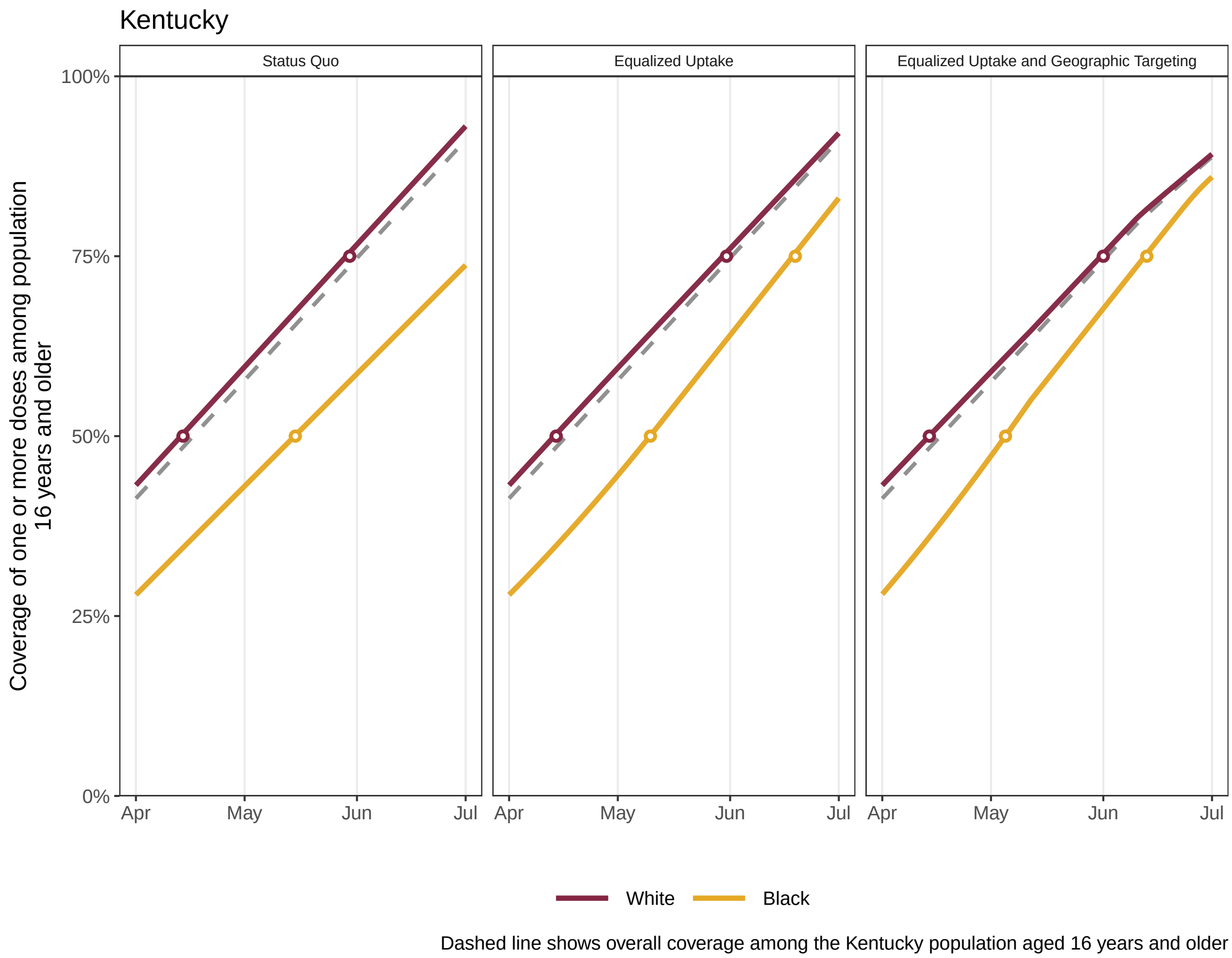
Kentucky
Status Quo
Equalized Uptake
Equalized Uptake and Geographic Targeting
100%
75%
50%
25%
0%
Apr
May
Jun
Jul
Coverage of one or more doses among population 16 years and older
White
Black
Dashed line shows overall coverage among the Kentucky population aged 16 years and older

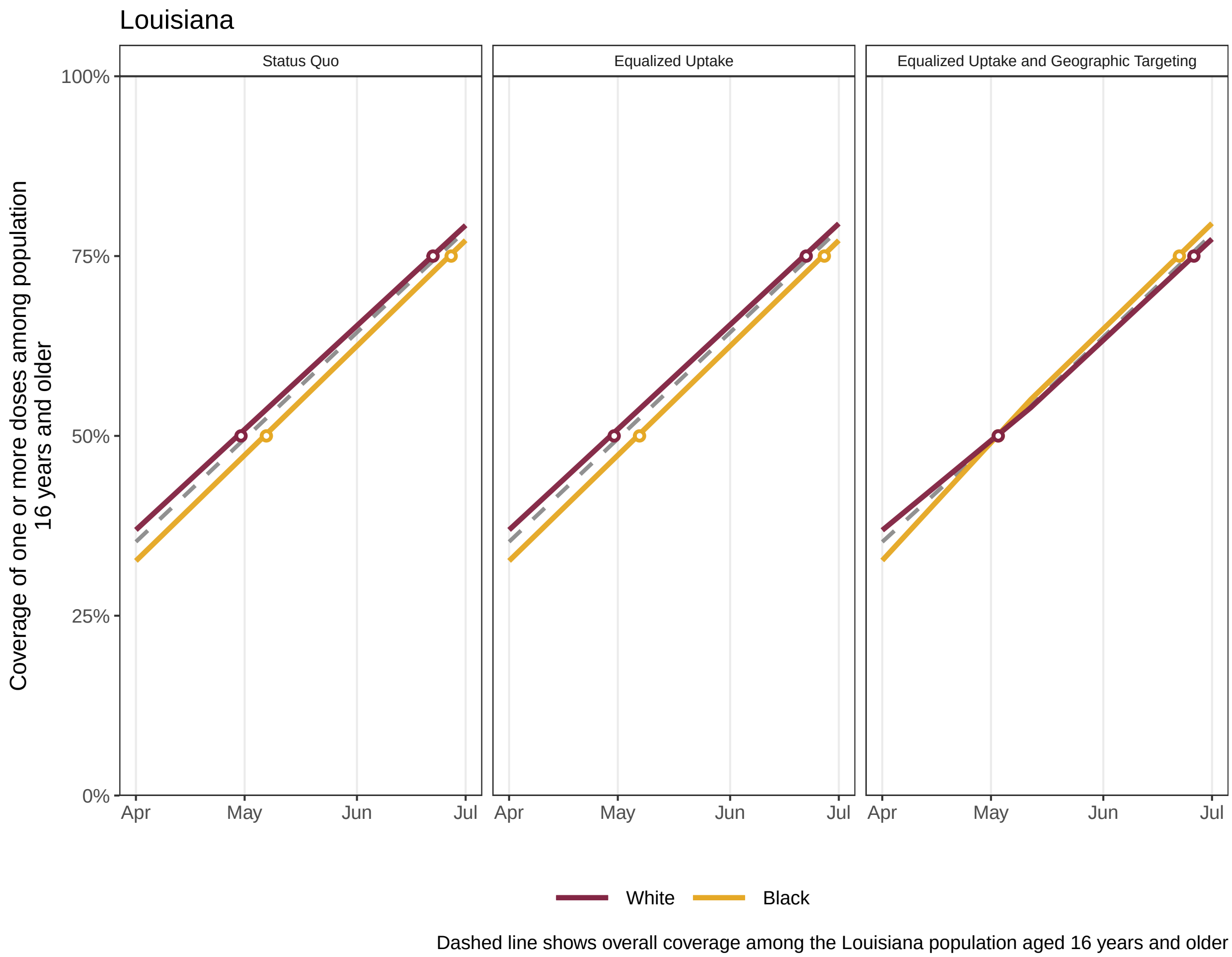

Louisiana
Status Quo
Equalized Uptake
Equalized Uptake and Geographic Targeting
100%
75%
50%
25%
0%
Apr
May
Jun
Jul
Coverage of one or more doses among population 16 years and older
White
Black
Dashed line shows overall coverage among the Louisiana population aged 16 years and older

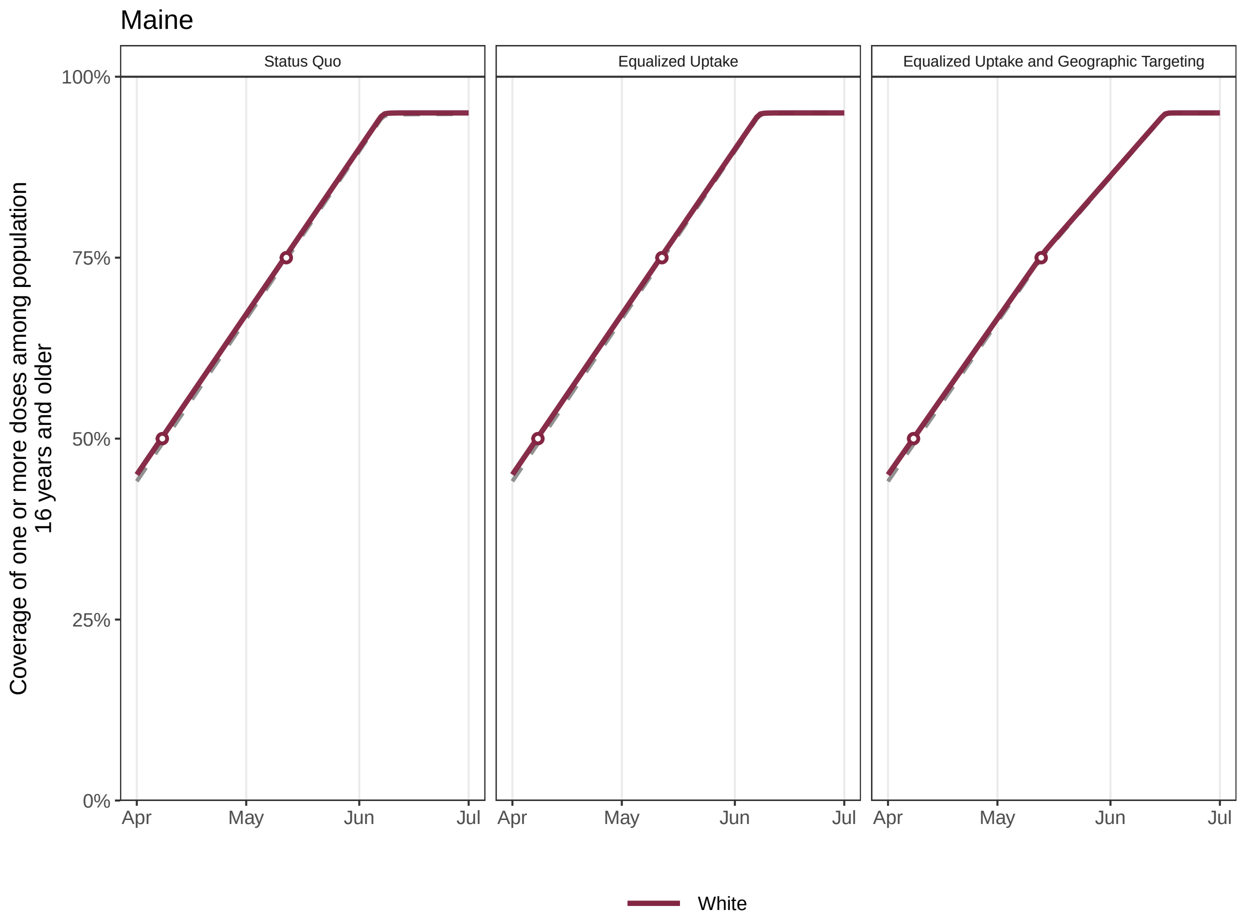


Dashed line shows overall coverage among the Maine population aged 16 years and older

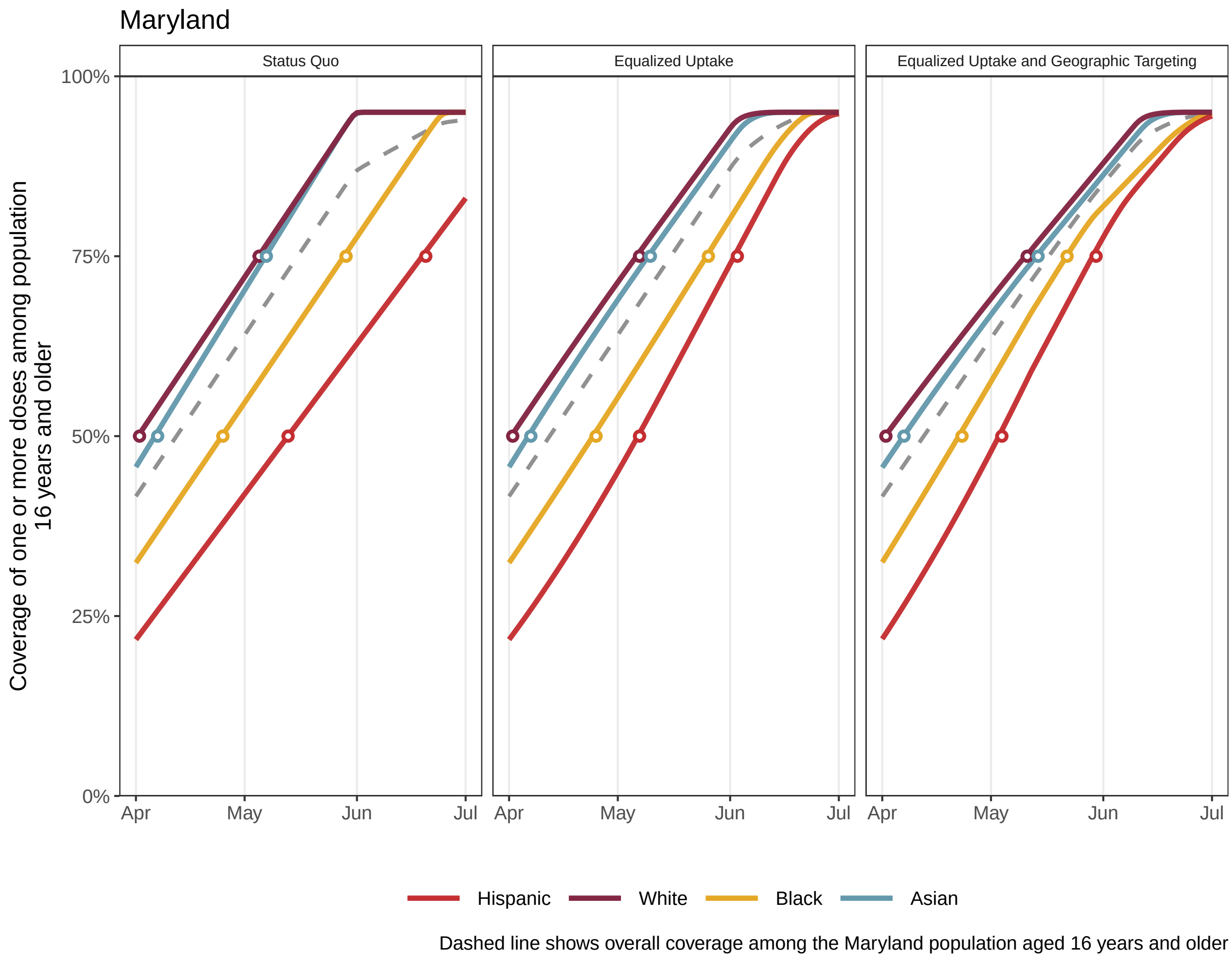
Maryland
Status Quo
Equalized Uptake
Equalized Uptake and Geographic Targeting
100%
75%
50%
25%
0%
Coverage of one or more doses among population 16 years and older
Apr
May
Jun
Jul
Hispanic
White
Black
Asian
Dashed line shows overall coverage among the Maryland population aged 16 years and older

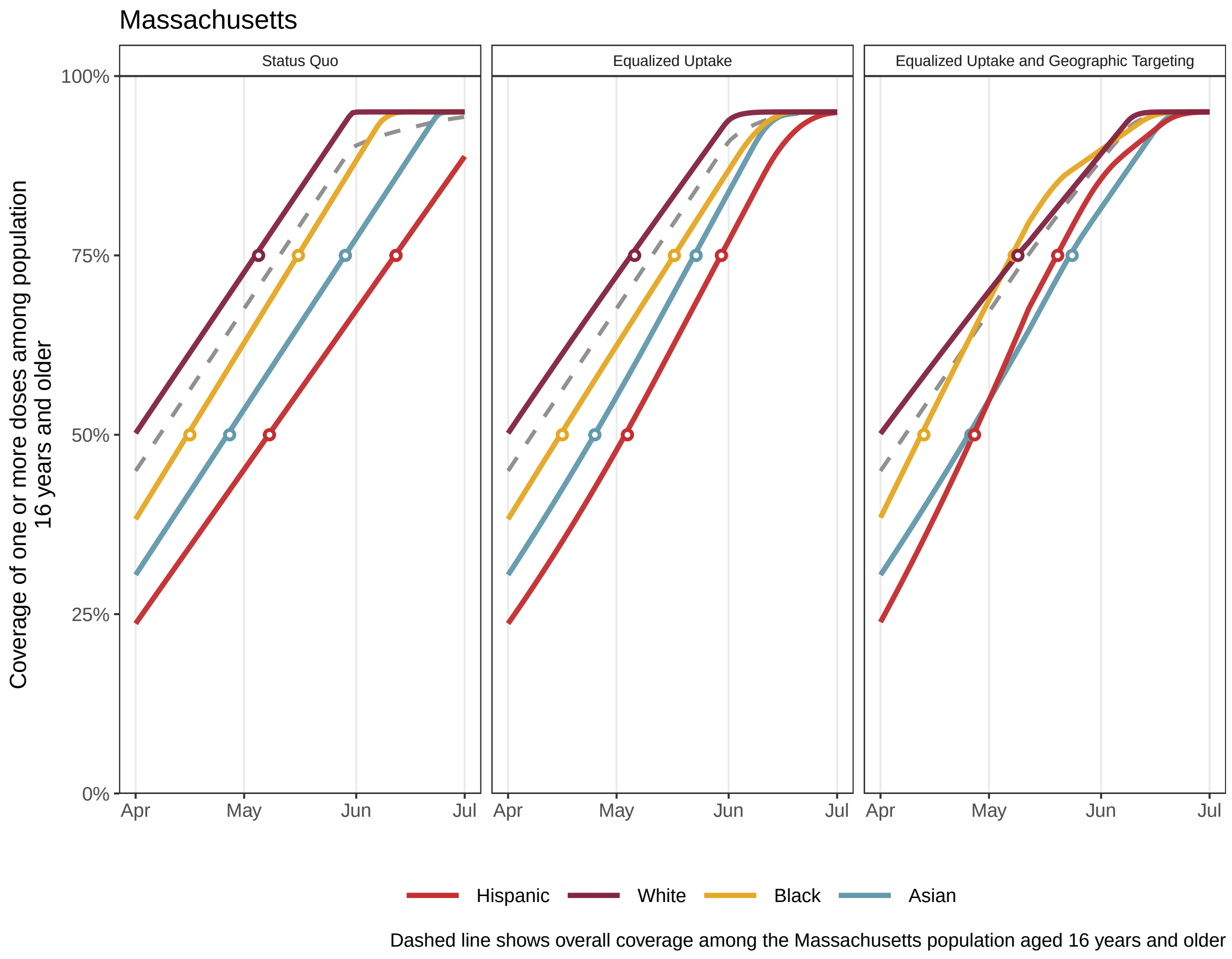
Massachusetts
Status Quo
Equalized Uptake
Equalized Uptake and Geographic Targeting
Coverage of one or more doses among population 16 years and older
100%
75%
50%
25%
0%
Apr
May
Jun
Jul
Hispanic
White
Black
Asian
Dashed line shows overall coverage among the Massachusetts population aged 16 years and older

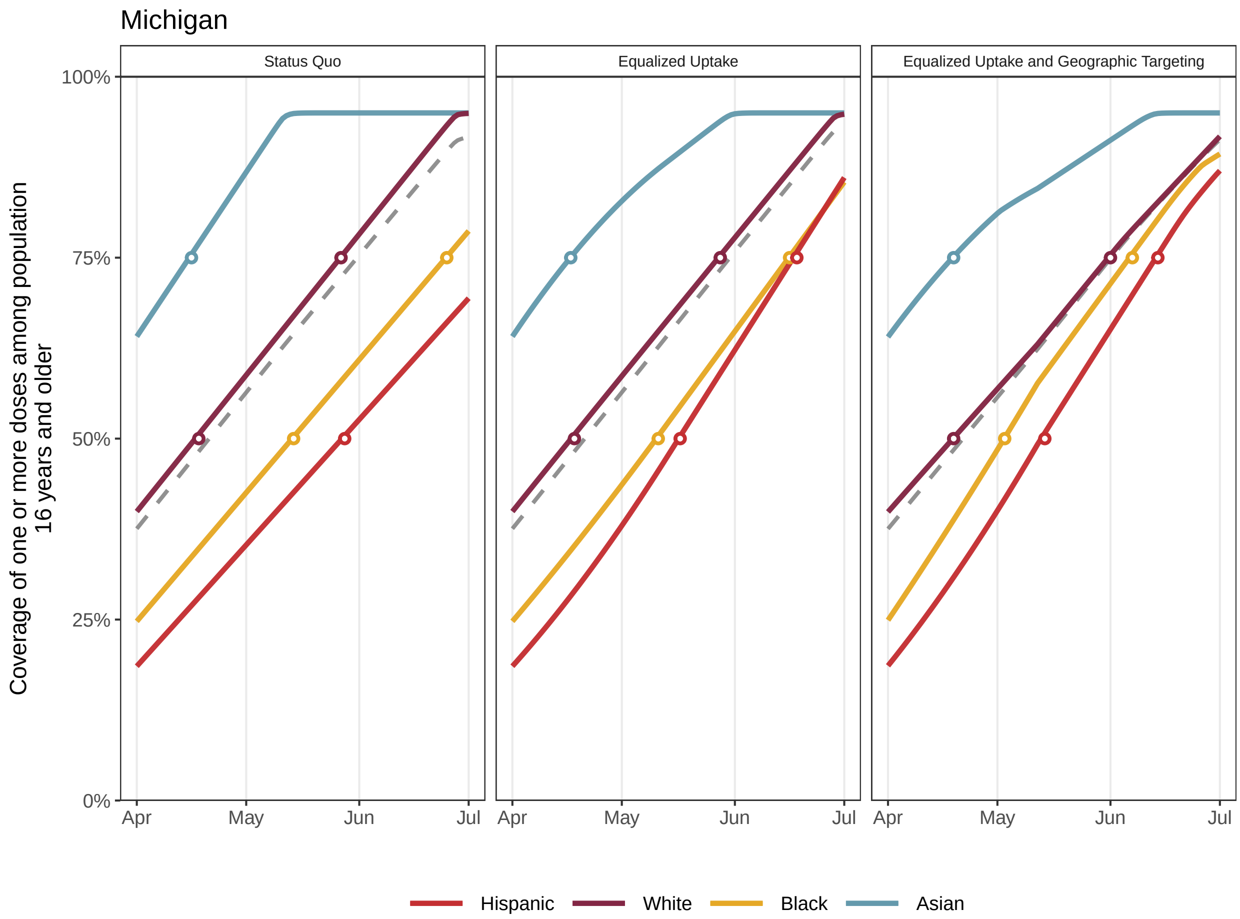


Dashed line shows overall coverage among the Michigan population aged 16 years and older

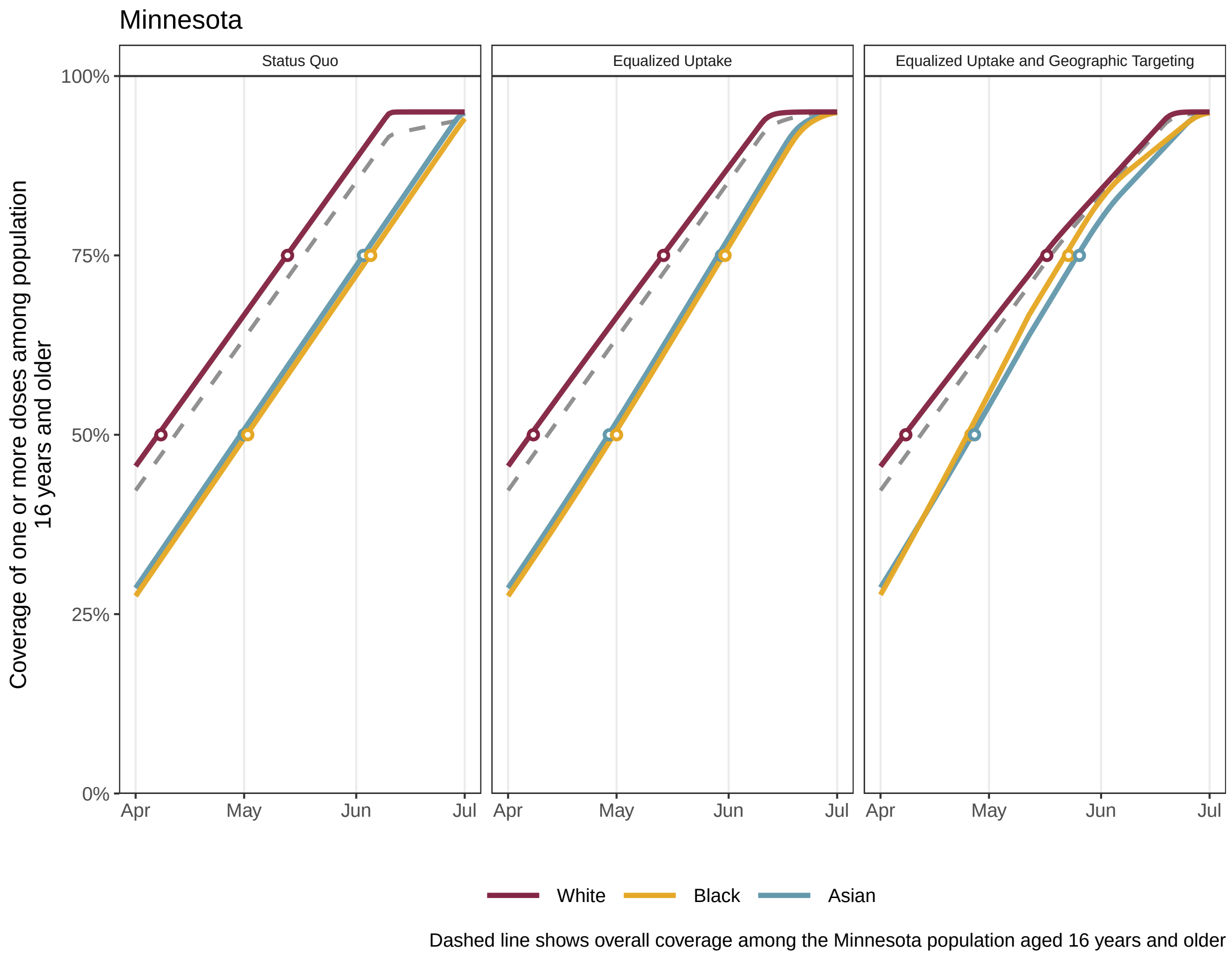
Minnesota
Status Quo
Equalized Uptake
Equalized Uptake and Geographic Targeting
Coverage of one or more doses among population 16 years and older
100%
75%
50%
25%
0%
Apr
May
Jun
Jul
White
Black
Asian
Dashed line shows overall coverage among the Minnesota population aged 16 years and older

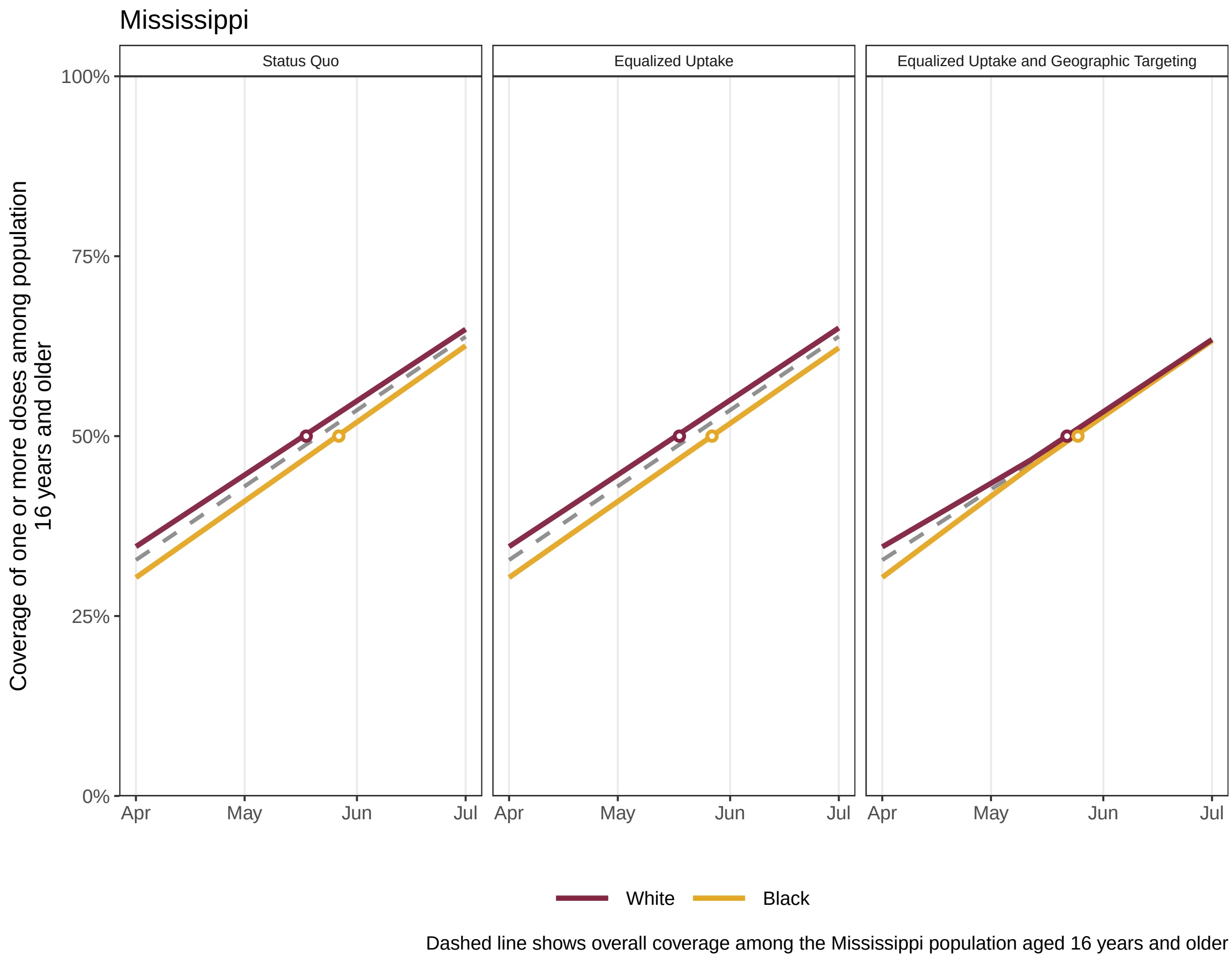
Mississippi
Status Quo
Equalized Uptake
Equalized Uptake and Geographic Targeting
Coverage of one or more doses among population 16 years and older
100%
75%
50%
25%
0%
Apr
May
Jun
Jul
White
Black
Dashed line shows overall coverage among the Mississippi population aged 16 years and older

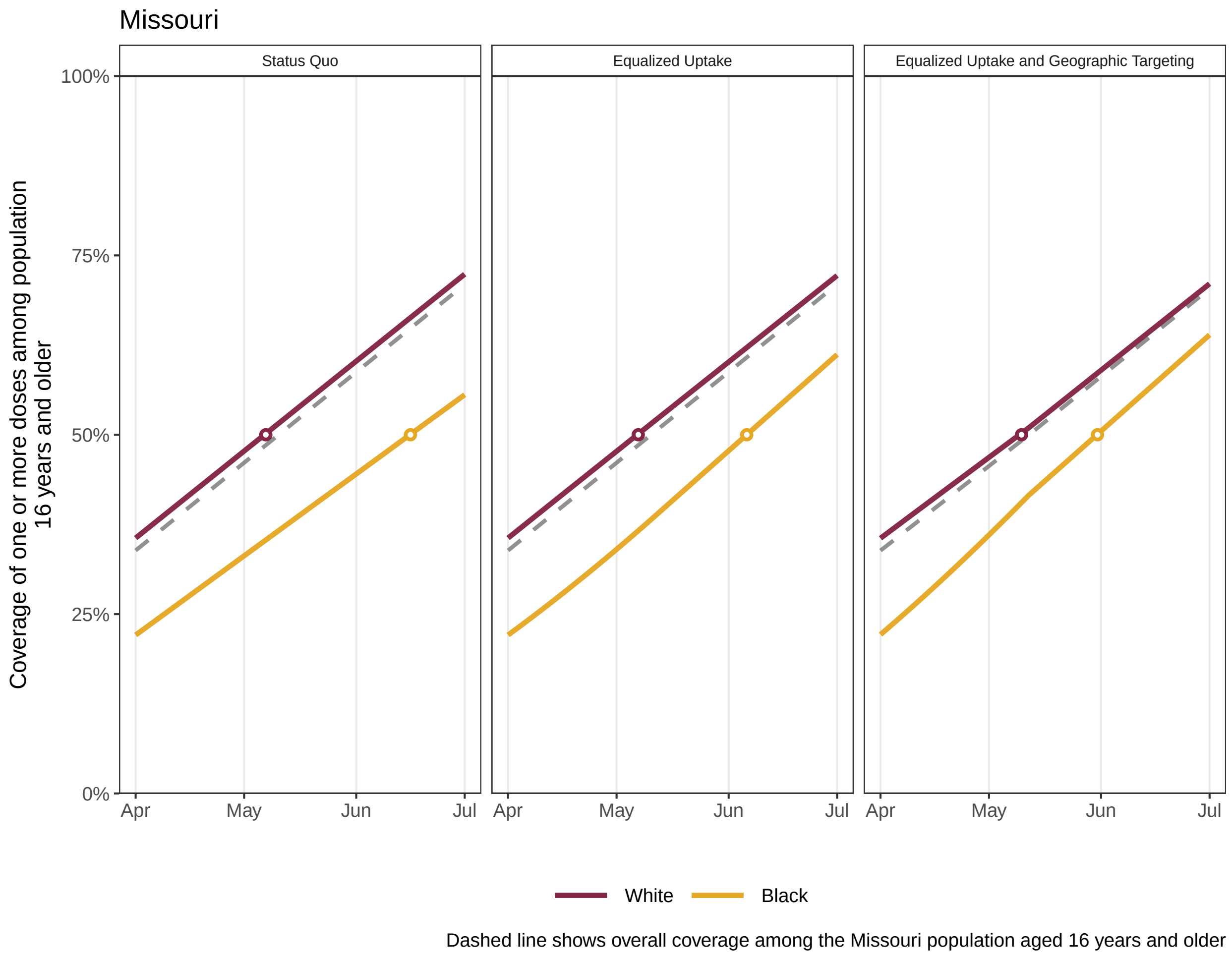

Missouri
Status Quo
Equalized Uptake
Equalized Uptake and Geographic Targeting
100%
75%
50%
25%
0%
Apr
May
Jun
Jul
Coverage of one or more doses among population
16 years and older
White
Black
Dashed line shows overall coverage among the Missouri population aged 16 years and older

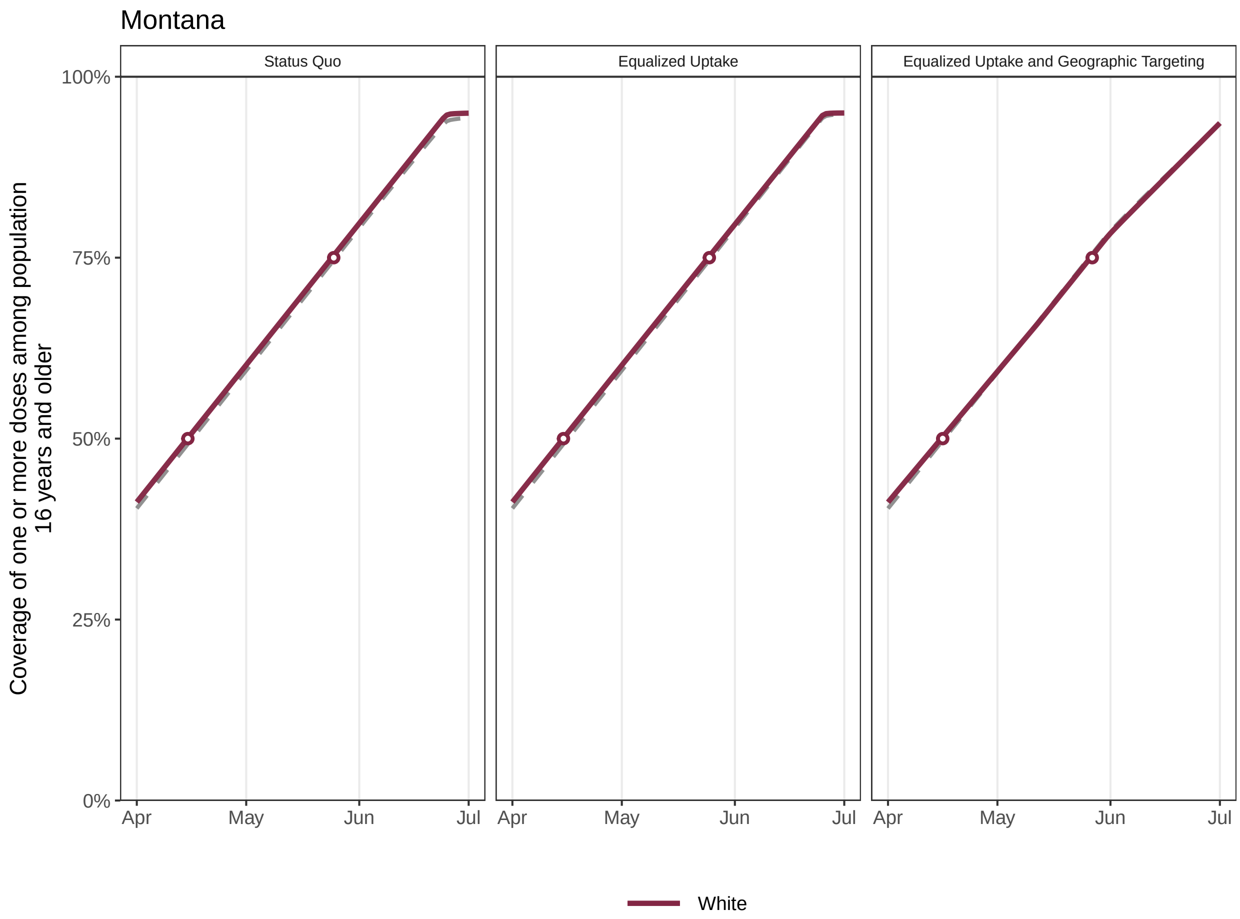

Dashed line shows overall coverage among the Montana population aged 16 years and older

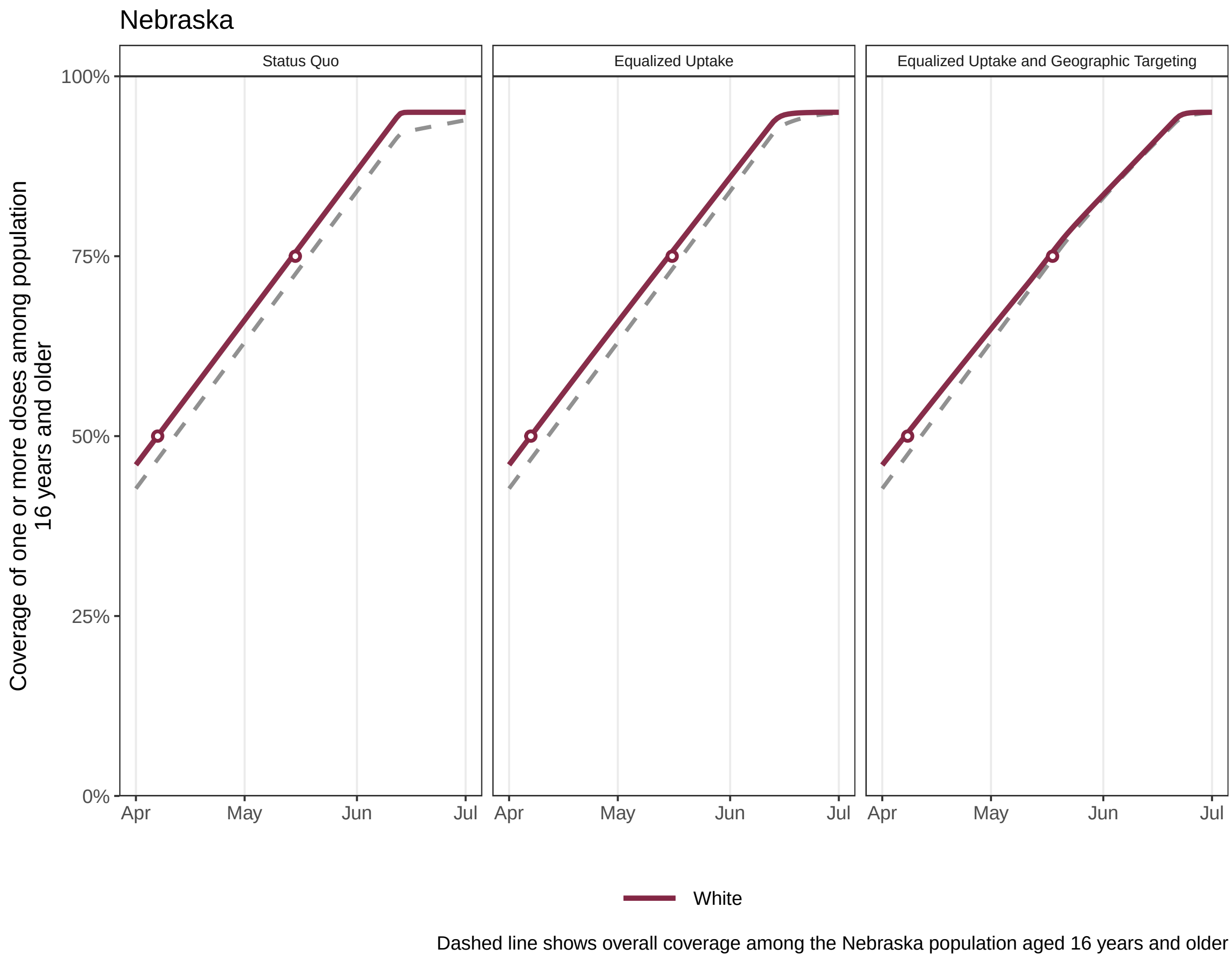

Nebraska
Status Quo
Equalized Uptake
Equalized Uptake and Geographic Targeting
Coverage of one or more doses among population 16 years and older
100%
75%
50%
25%
0%
Apr
May
Jun
Jul
White
Dashed line shows overall coverage among the Nebraska population aged 16 years and older

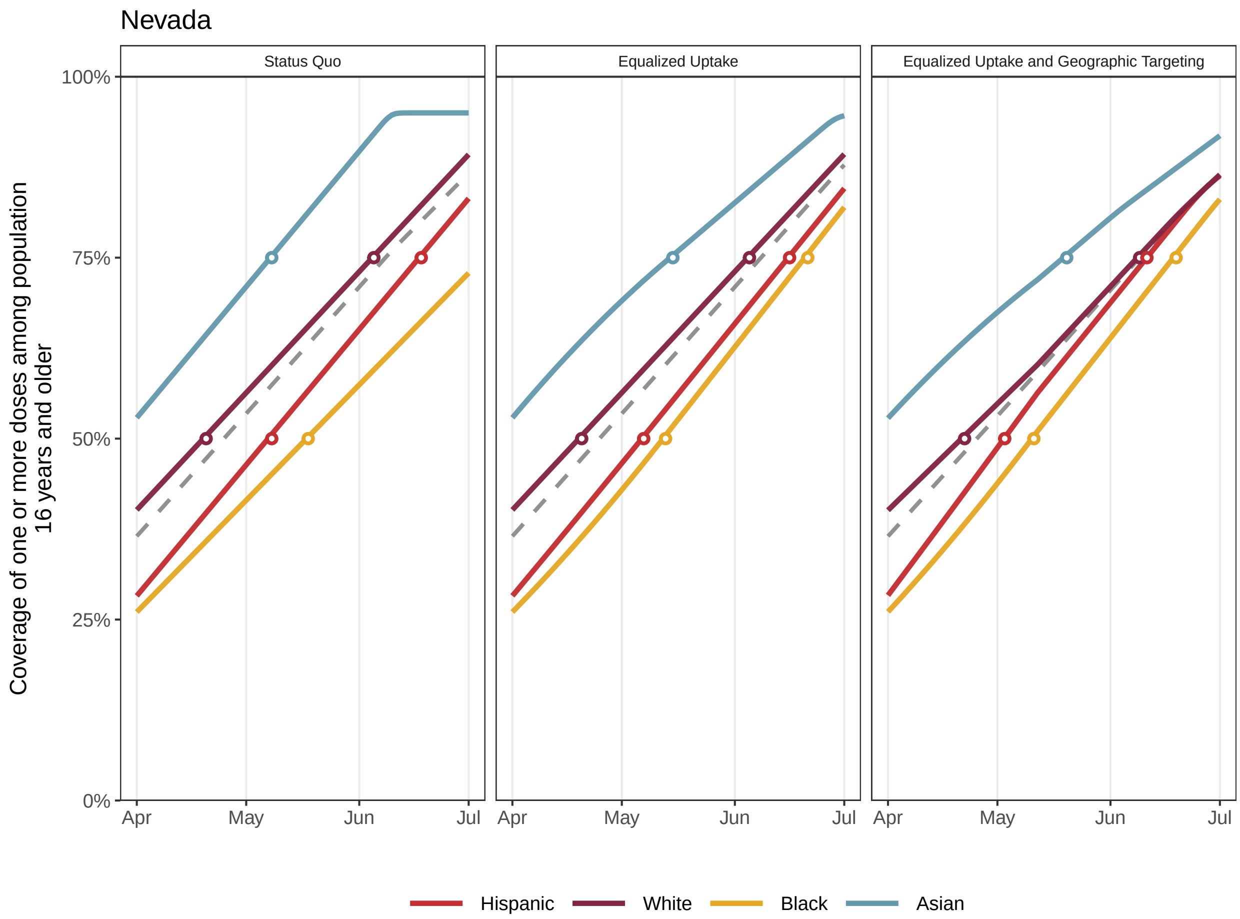

Dashed line shows overall coverage among the Nevada population aged 16 years and older

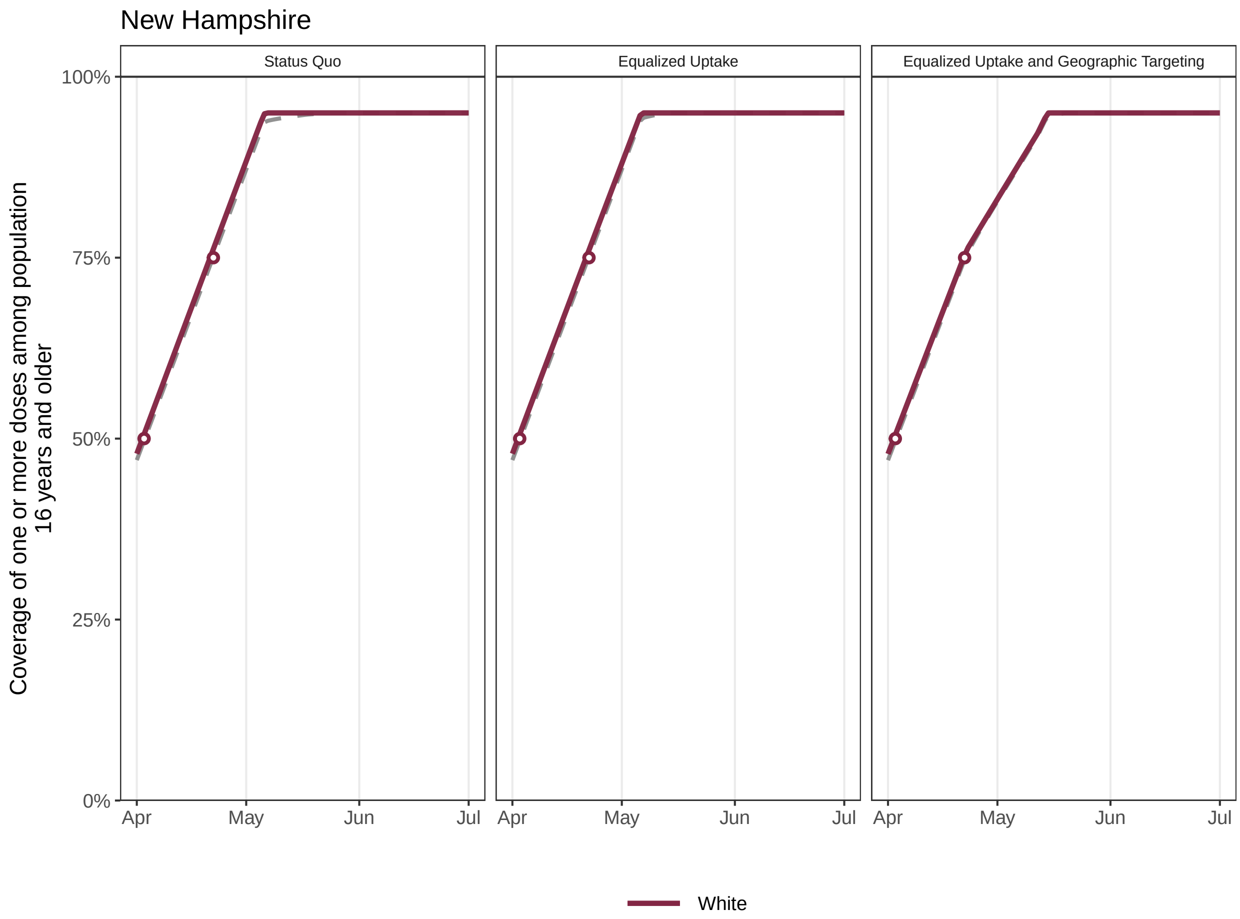


Dashed line shows overall coverage among the New Hampshire population aged 16 years and older

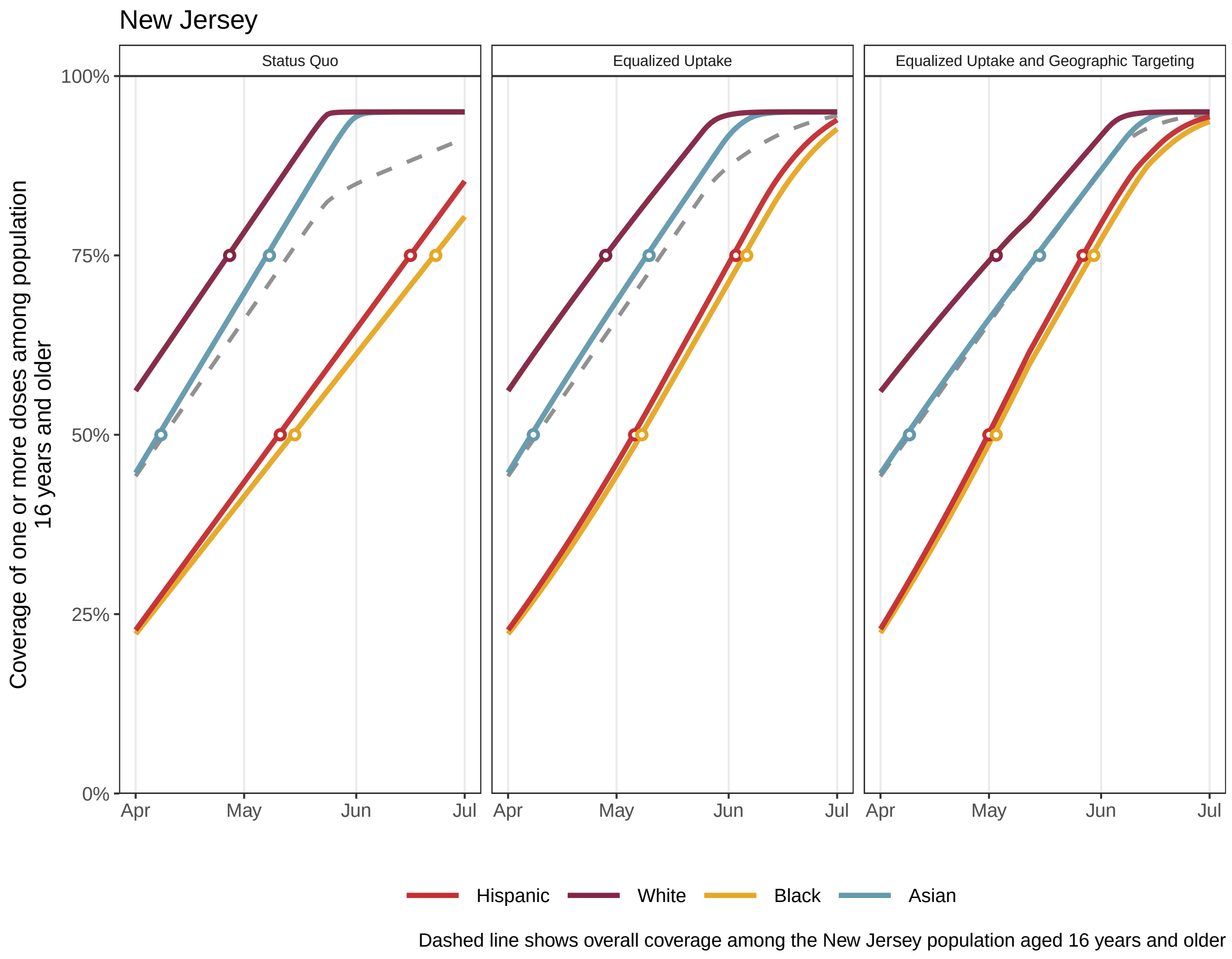
New Jersey
Status Quo
Equalized Uptake
Equalized Uptake and Geographic Targeting
Coverage of one or more doses among population 16 years and older
100%
75%
50%
25%
0%
Apr
May
Jun
Jul
Hispanic
White
Black
Asian
Dashed line shows overall coverage among the New Jersey population aged 16 years and older

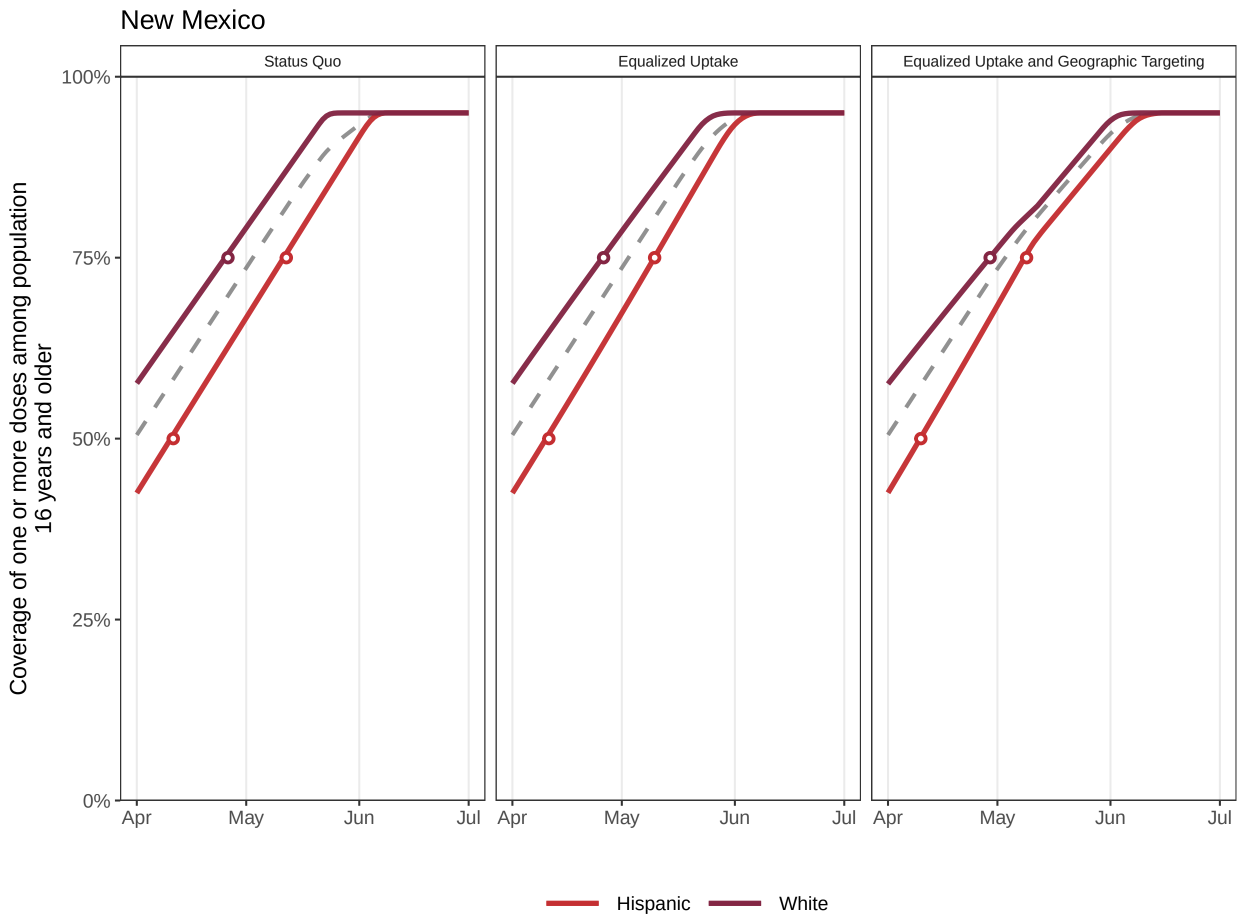


Dashed line shows overall coverage among the New Mexico population aged 16 years and older

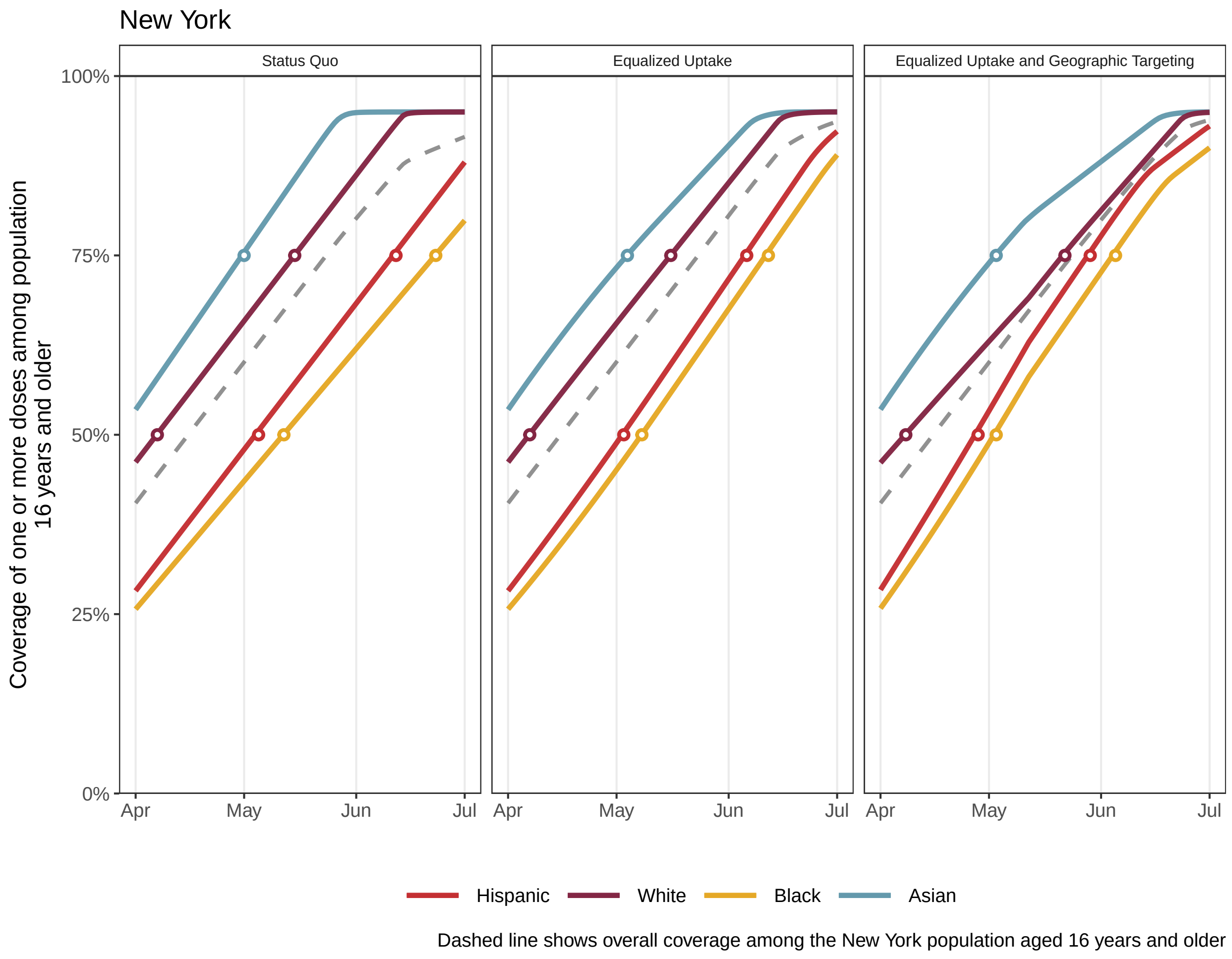
New York
Status Quo
Equalized Uptake
Equalized Uptake and Geographic Targeting
Coverage of one or more doses among population 16 years and older
100%
75%
50%
25%
0%
Apr
May
Jun
Jul
Hispanic
White
Black
Asian
Dashed line shows overall coverage among the New York population aged 16 years and older

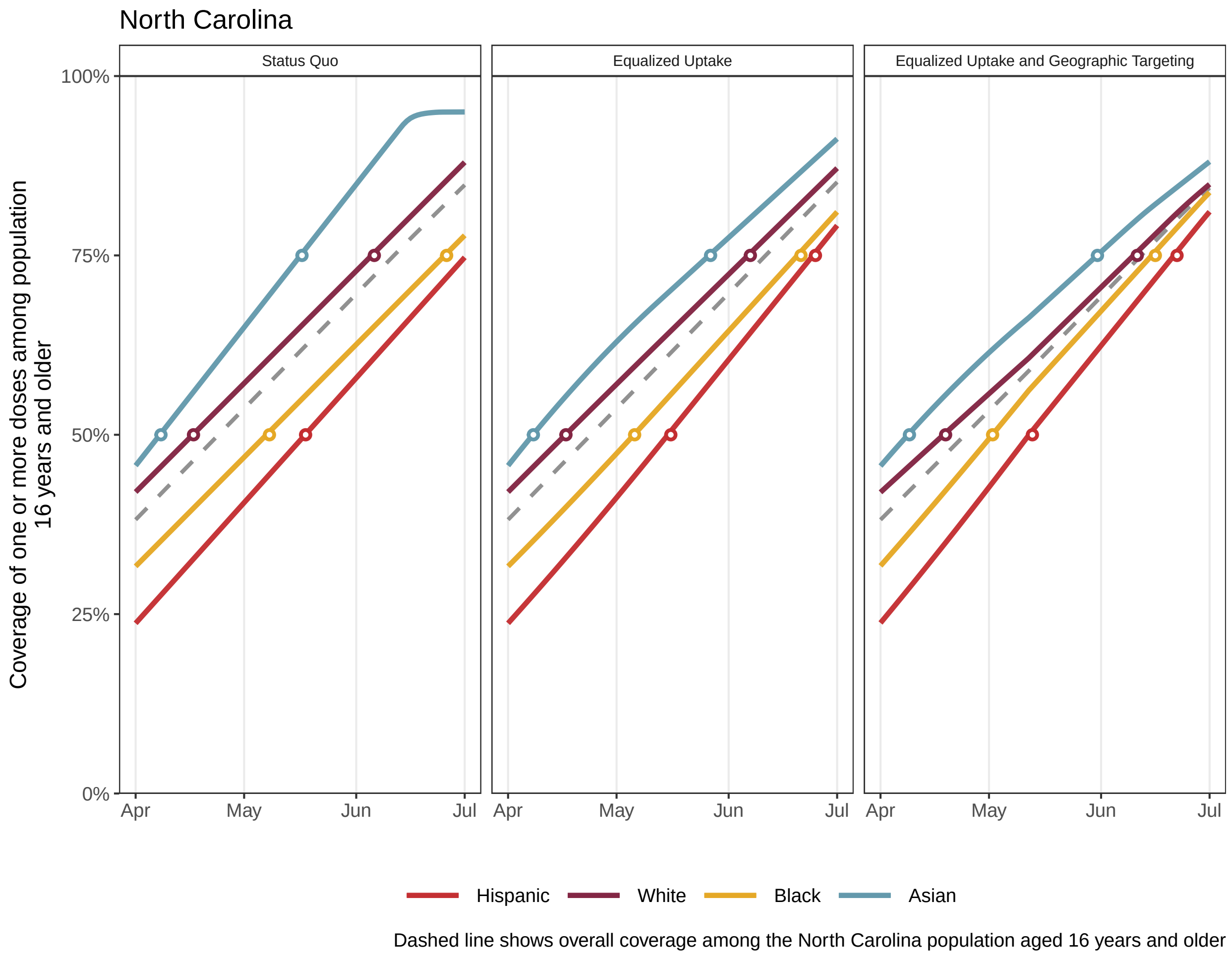

North Carolina
Status Quo
Equalized Uptake
Equalized Uptake and Geographic Targeting
Coverage of one or more doses among population 16 years and older
100%
75%
50%
25%
0%
Apr
May
Jun
Jul
Hispanic
White
Black
Asian
Dashed line shows overall coverage among the North Carolina population aged 16 years and older

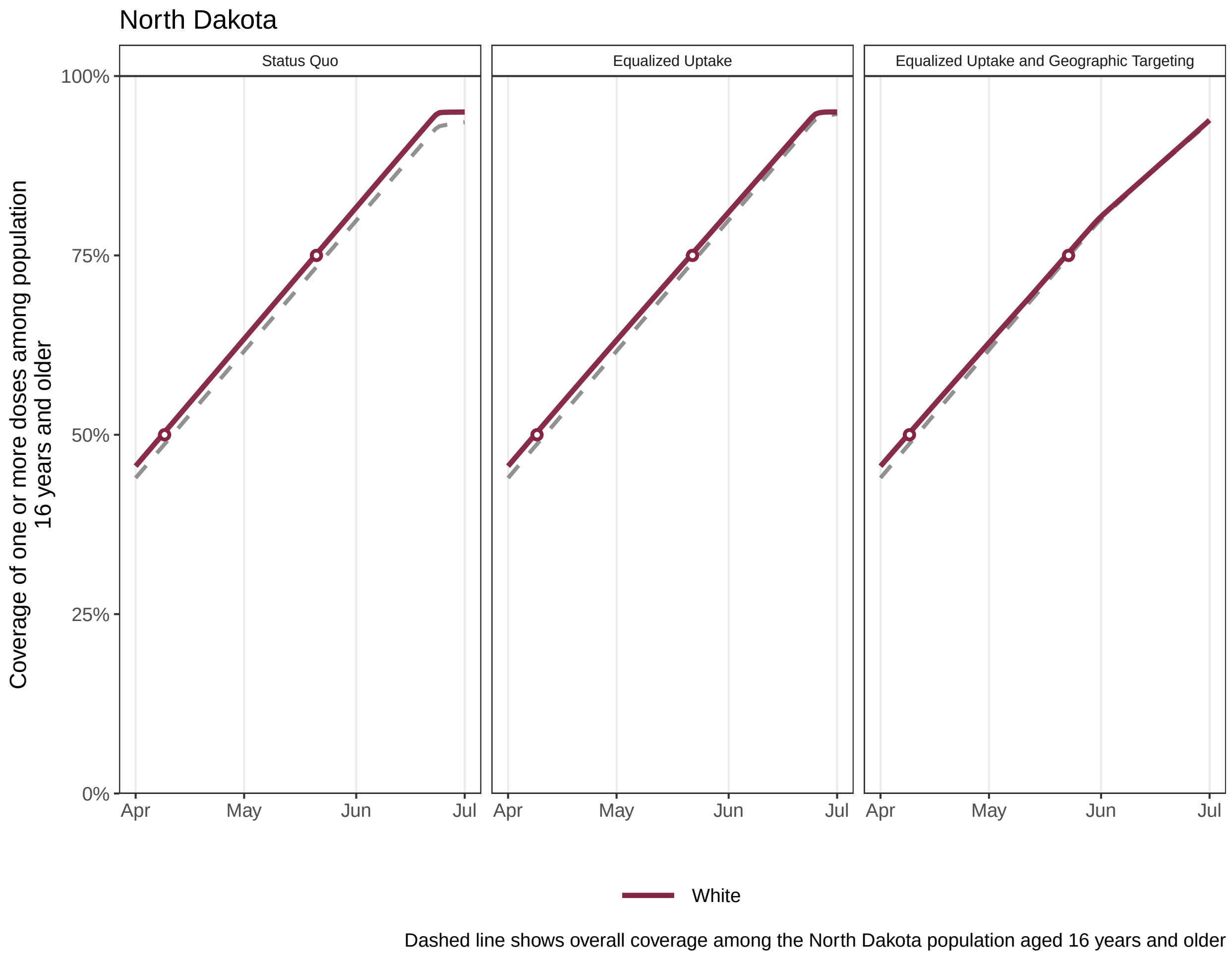

North Dakota
Status Quo
Equalized Uptake
Equalized Uptake and Geographic Targeting
Coverage of one or more doses among population 16 years and older
100%
75%
50%
25%
0%
Apr
May
Jun
Jul
White
Dashed line shows overall coverage among the North Dakota population aged 16 years and older

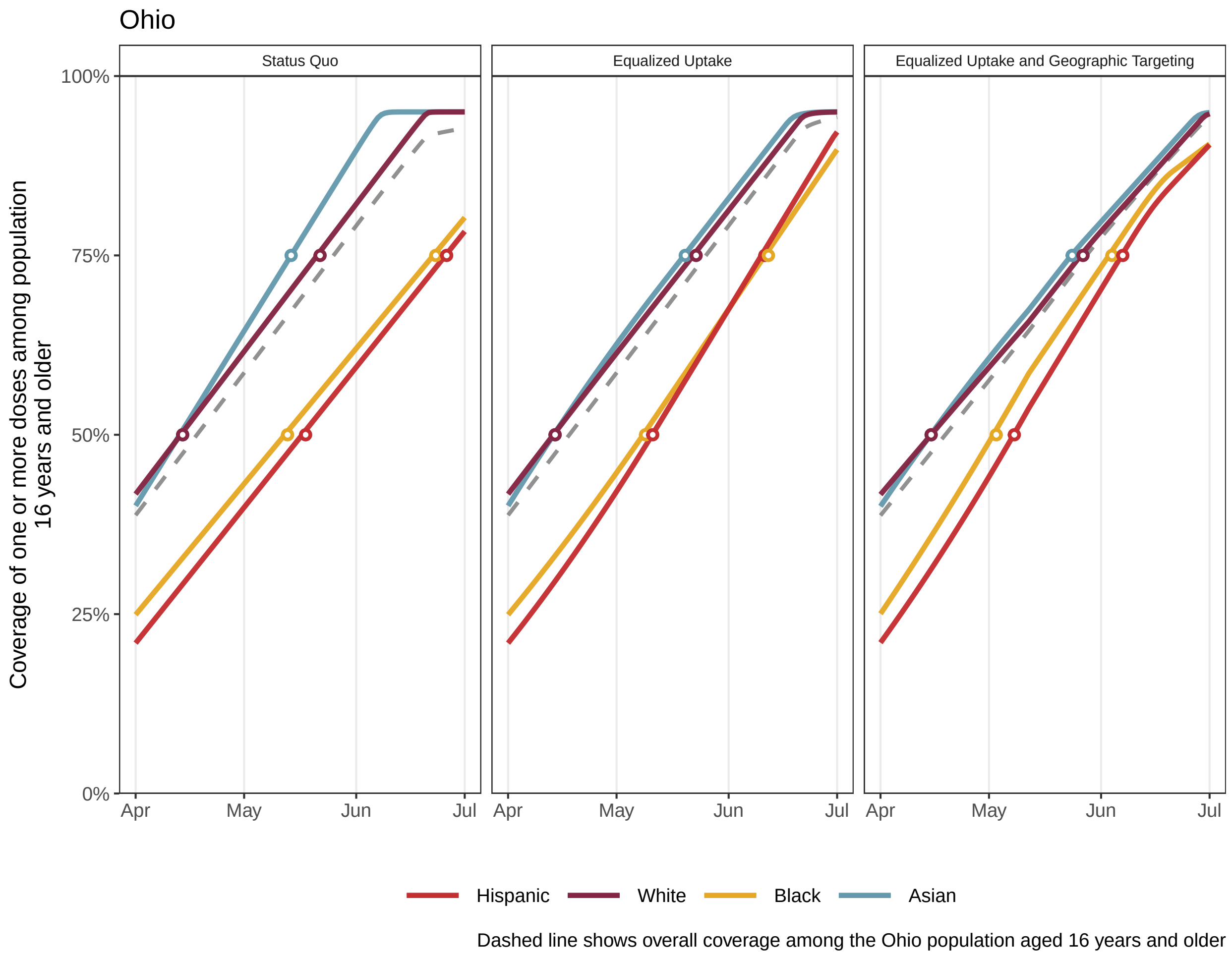
Ohio
Status Quo
Equalized Uptake
Equalized Uptake and Geographic Targeting
Coverage of one or more doses among population 16 years and older
100%
75%
50%
25%
0%
Apr
May
Jun
Jul
Hispanic
White
Black
Asian
Dashed line shows overall coverage among the Ohio population aged 16 years and older

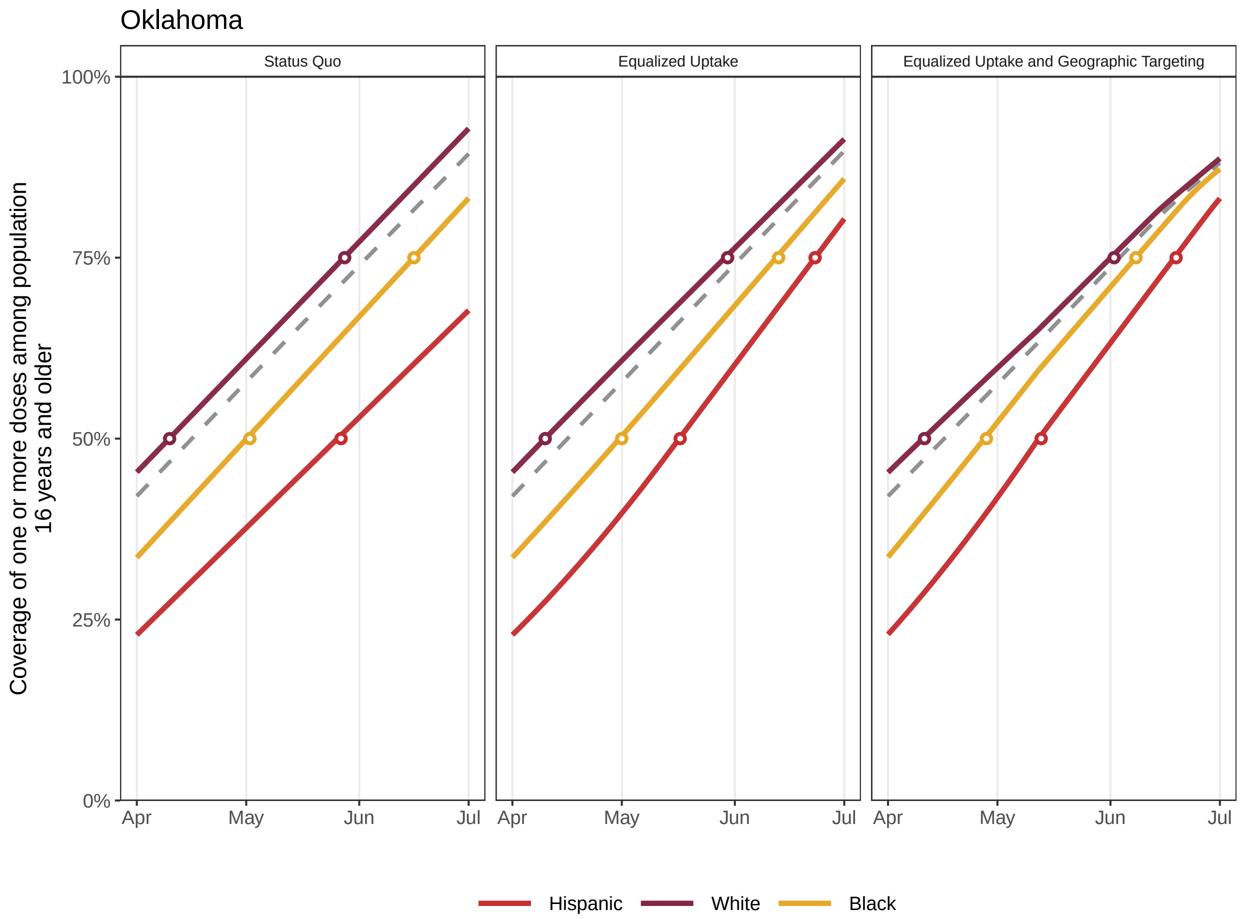


Dashed line shows overall coverage among the Oklahoma population aged 16 years and older

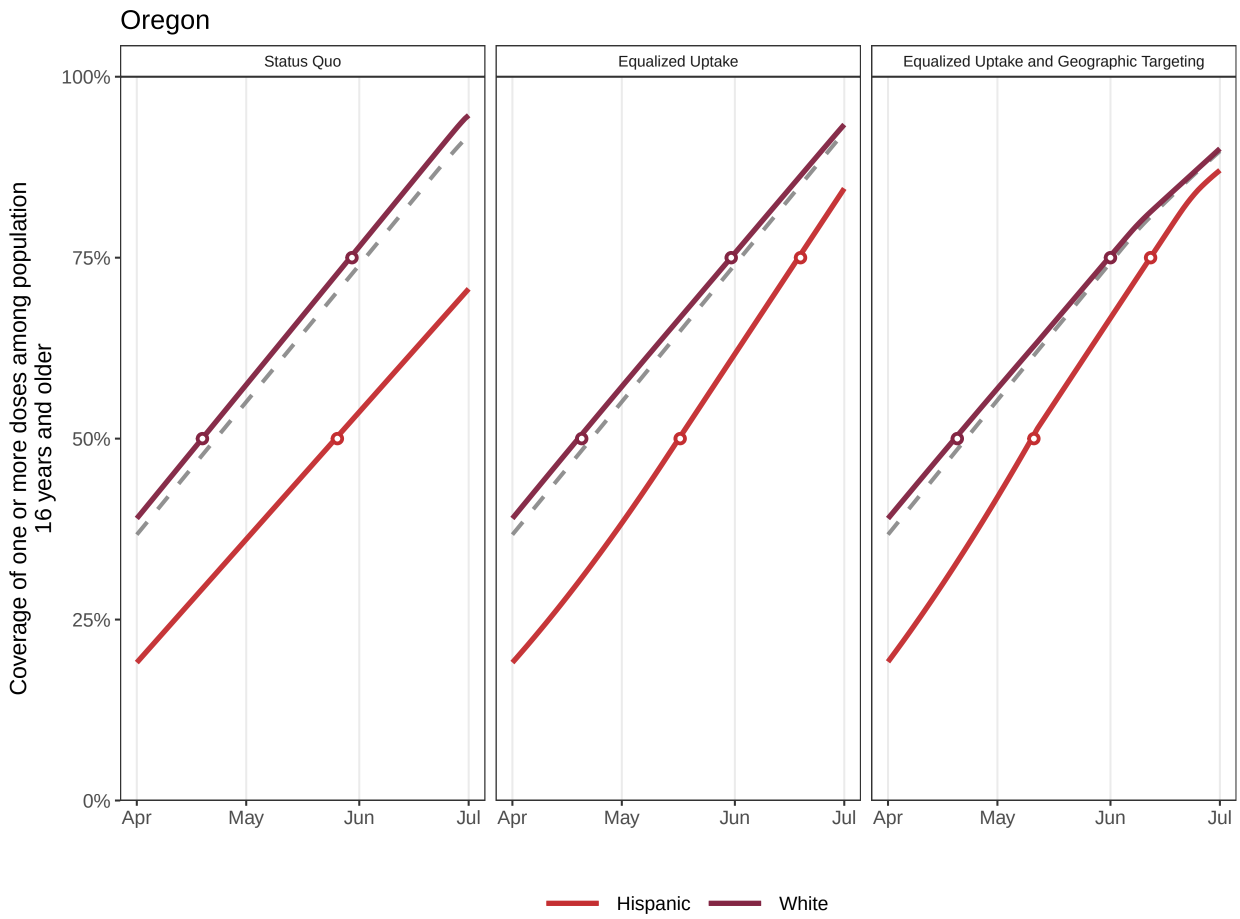


Dashed line shows overall coverage among the Oregon population aged 16 years and older

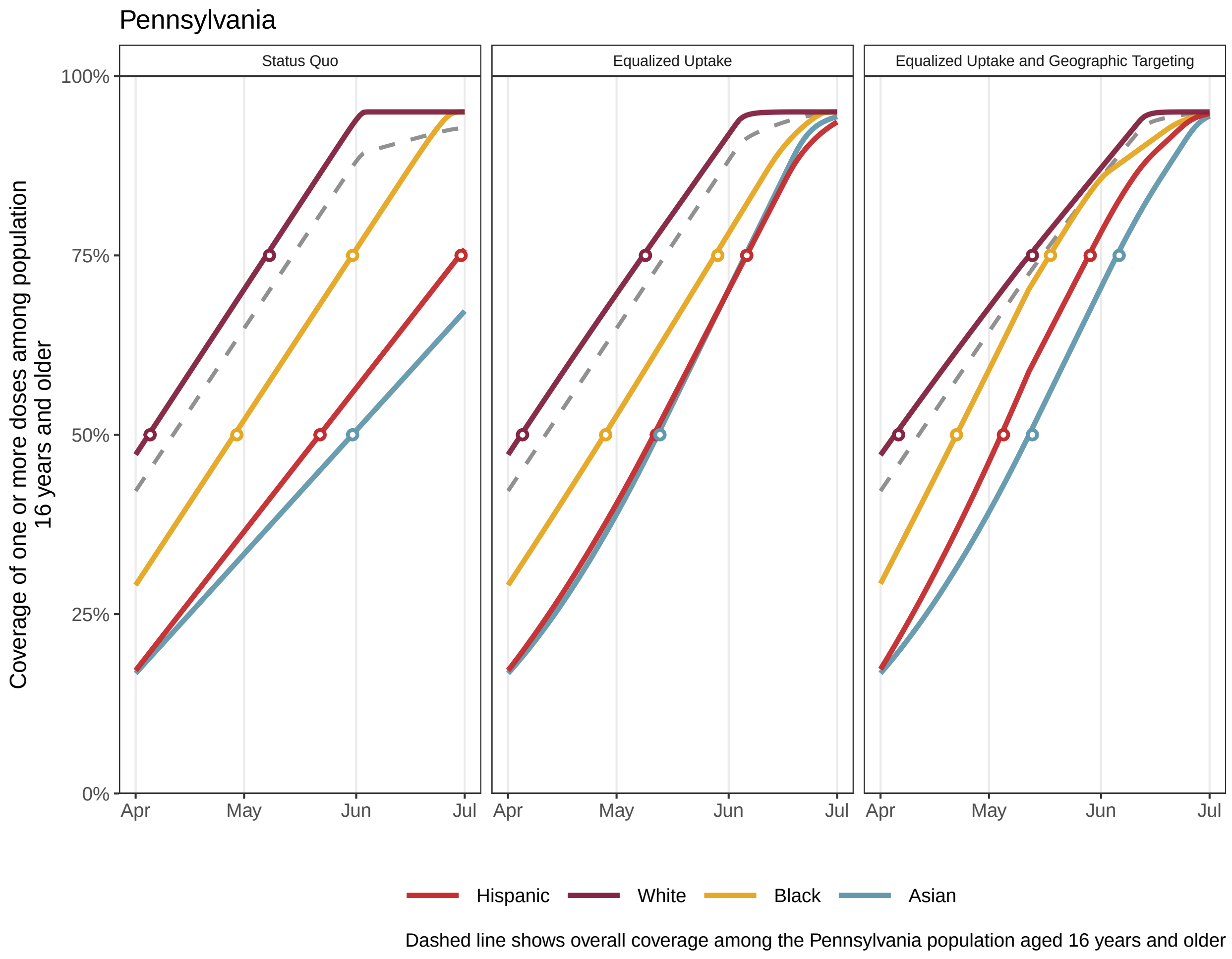

Pennsylvania
Status Quo
Equalized Uptake
Equalized Uptake and Geographic Targeting
Coverage of one or more doses among population
16 years and older
100%
75%
50%
25%
0%
Apr
May
Jun
Jul
Hispanic
White
Black
Asian
Dashed line shows overall coverage among the Pennsylvania population aged 16 years and older

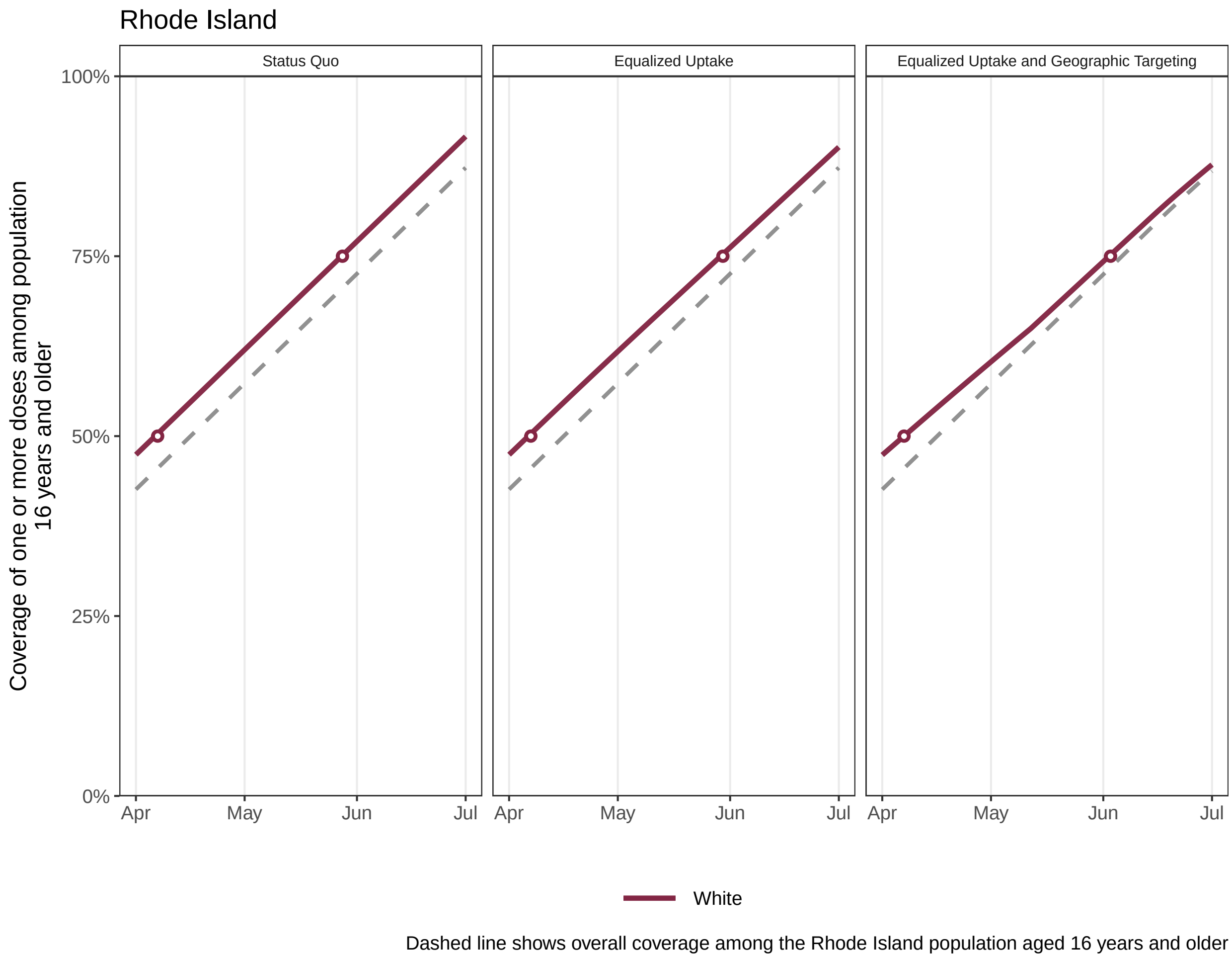
Rhode Island
Status Quo
Equalized Uptake
Equalized Uptake and Geographic Targeting
Coverage of one or more doses among population 16 years and older
100%
75%
50%
25%
0%
Apr
May
Jun
Jul
White
Dashed line shows overall coverage among the Rhode Island population aged 16 years and older

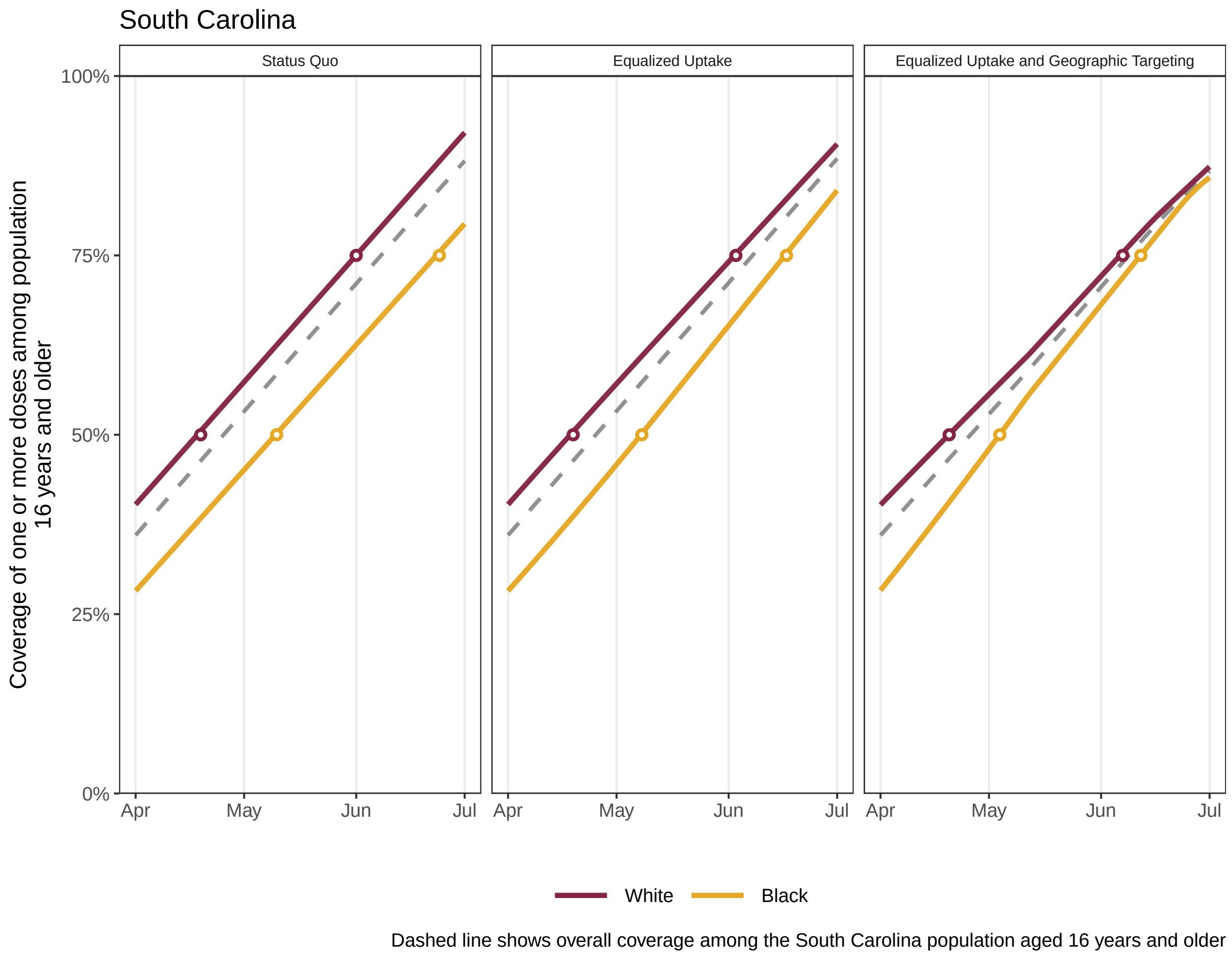
South Carolina
Status Quo
Equalized Uptake
Equalized Uptake and Geographic Targeting
Coverage of one or more doses among population 16 years and older
100%
75%
50%
25%
0%
Apr
May
Jun
Jul
White
Black
Dashed line shows overall coverage among the South Carolina population aged 16 years and older

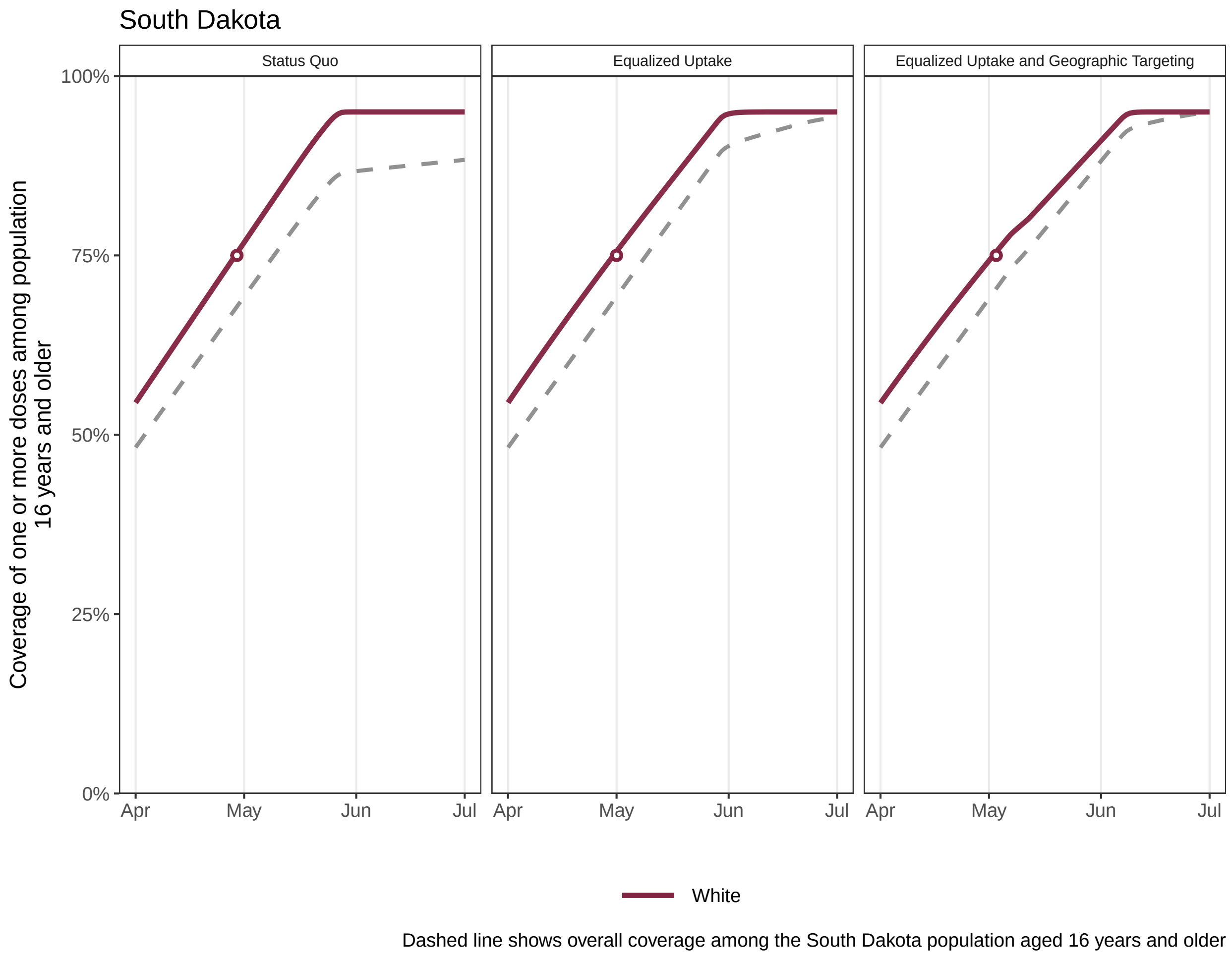
South Dakota
Status Quo
Equalized Uptake
Equalized Uptake and Geographic Targeting
Coverage of one or more doses among population 16 years and older
100%
75%
50%
25%
0%
Apr
May
Jun
Jul
White
Dashed line shows overall coverage among the South Dakota population aged 16 years and older

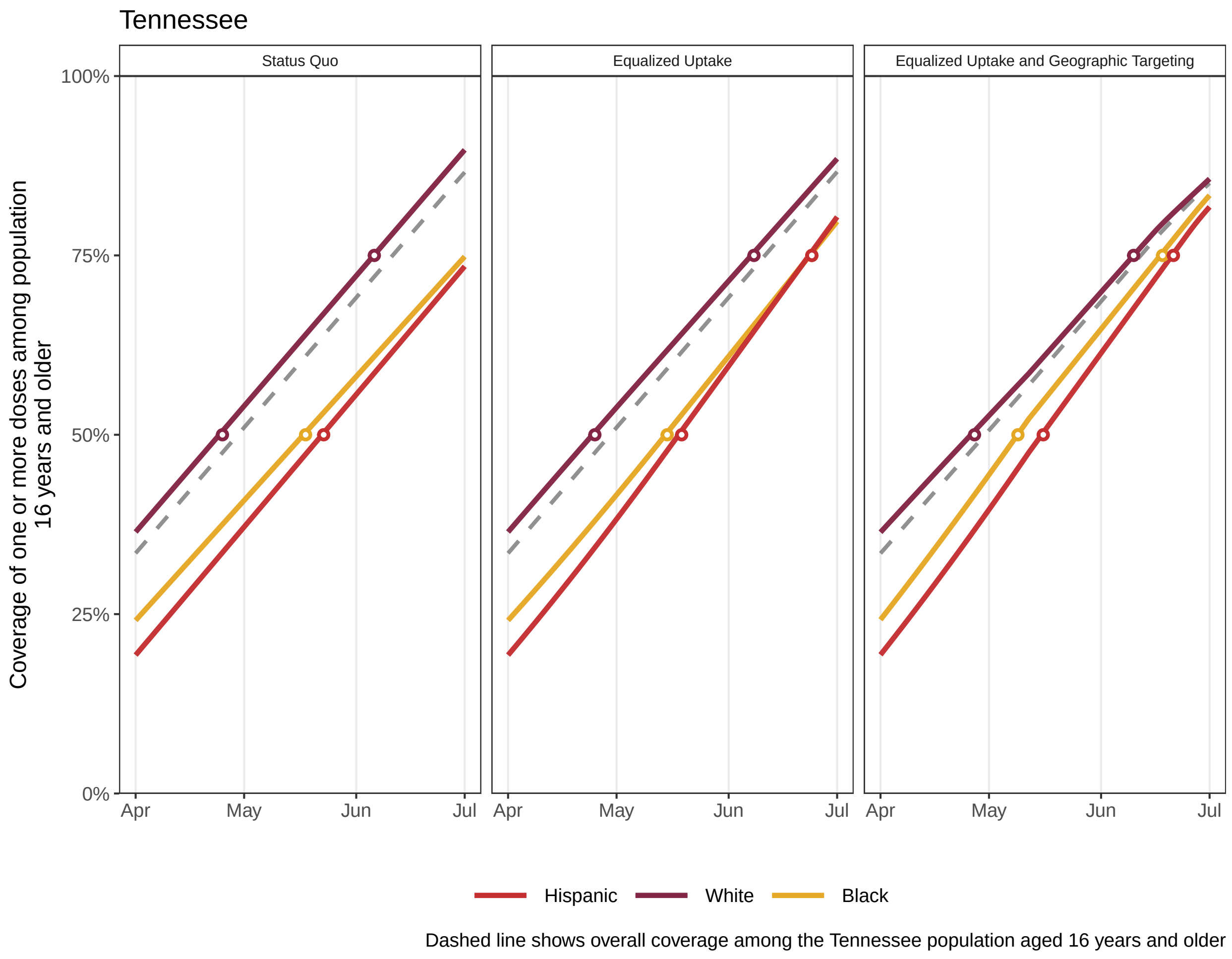
Tennessee
Status Quo
Equalized Uptake
Equalized Uptake and Geographic Targeting
Coverage of one or more doses among population 16 years and older
100%
75%
50%
25%
0%
Apr
May
Jun
Jul
Hispanic
White
Black
Dashed line shows overall coverage among the Tennessee population aged 16 years and older

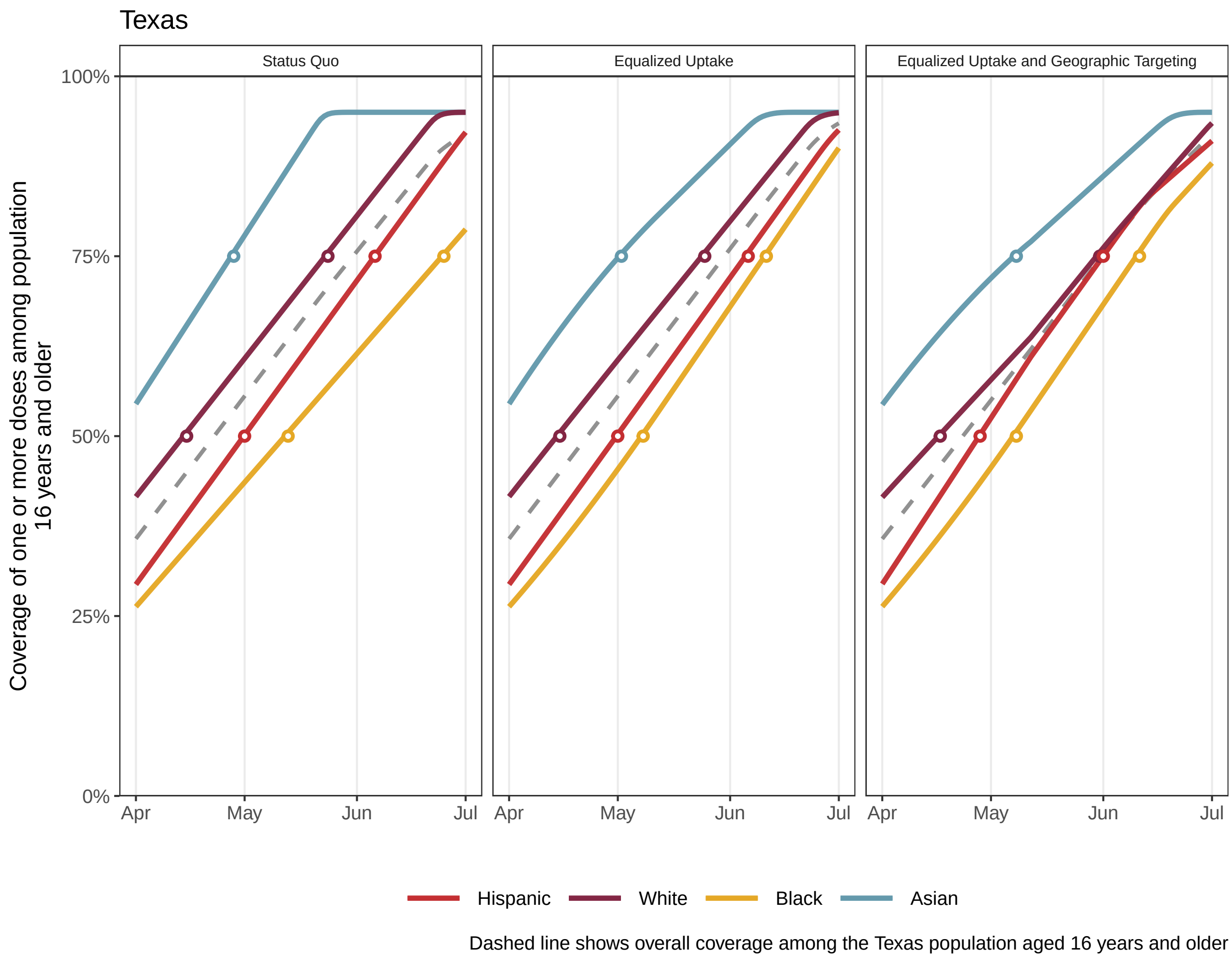
Texas
Status Quo
Equalized Uptake
Equalized Uptake and Geographic Targeting
Coverage of one or more doses among population 16 years and older
100%
75%
50%
25%
0%
Apr
May
Jun
Jul
Hispanic
White
Black
Asian
Dashed line shows overall coverage among the Texas population aged 16 years and older

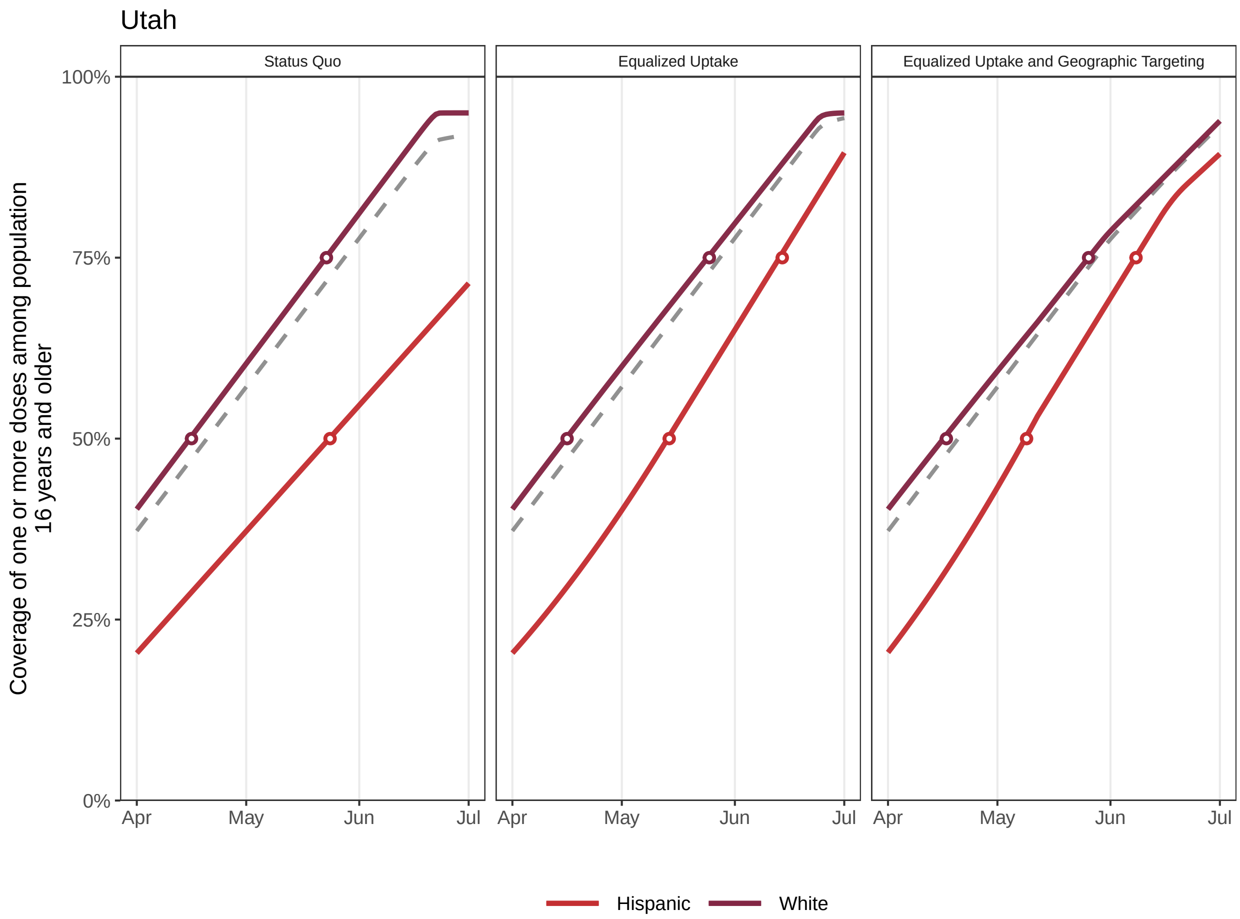


Dashed line shows overall coverage among the Utah population aged 16 years and older

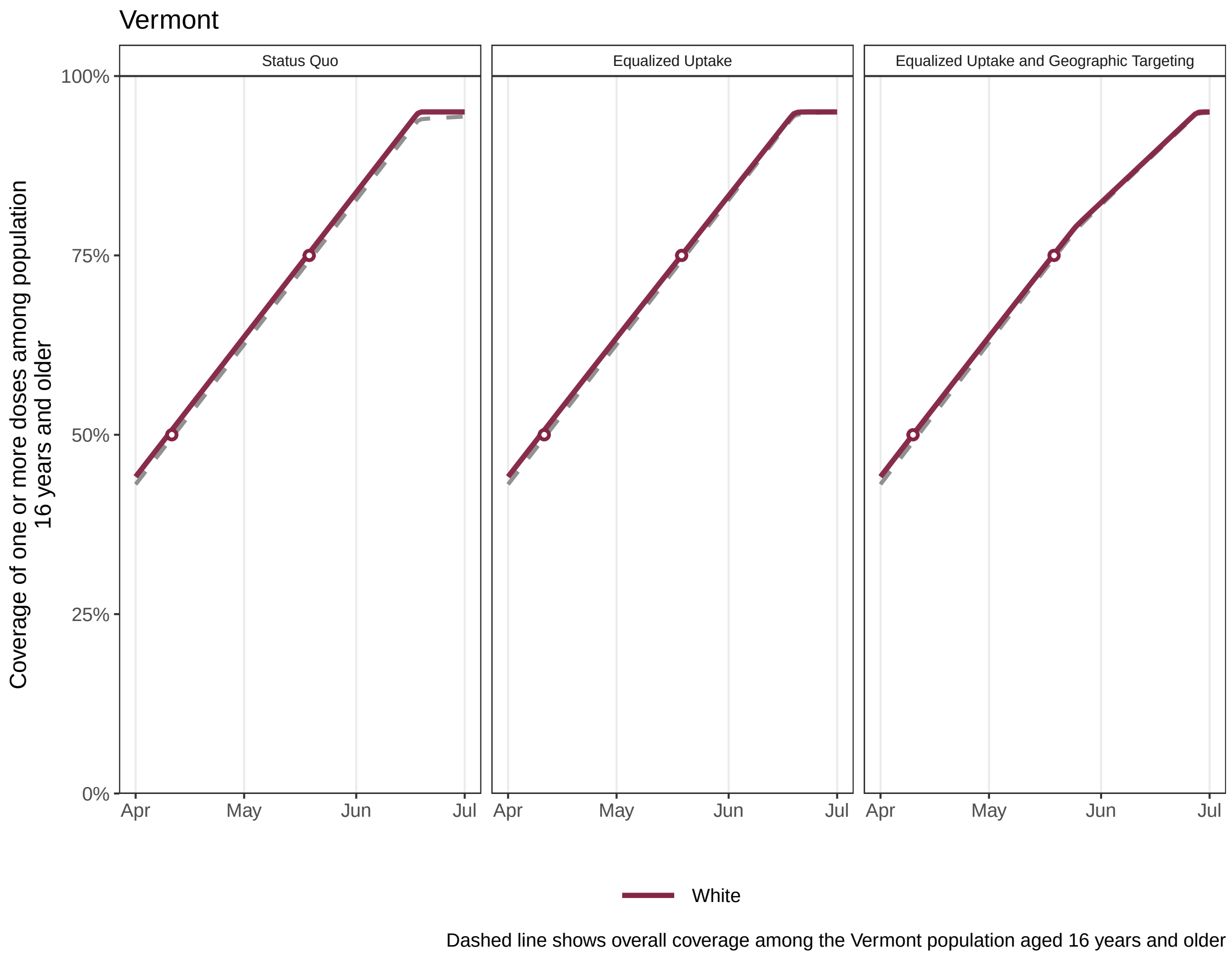
Vermont
Status Quo
Equalized Uptake
Equalized Uptake and Geographic Targeting
Coverage of one or more doses among population 16 years and older
100%
75%
50%
25%
0%
Apr
May
Jun
Jul
White
Dashed line shows overall coverage among the Vermont population aged 16 years and older

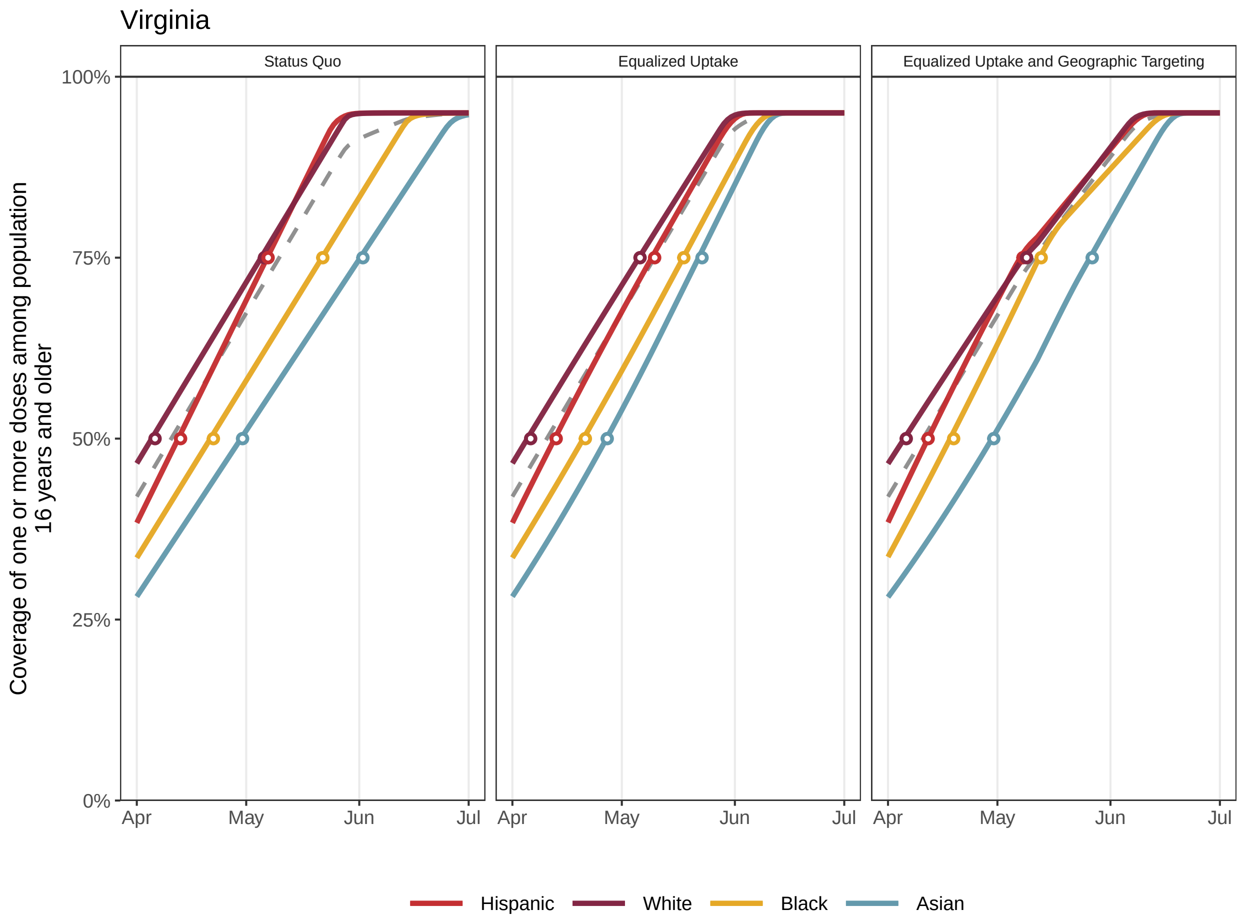


Dashed line shows overall coverage among the Virginia population aged 16 years and older

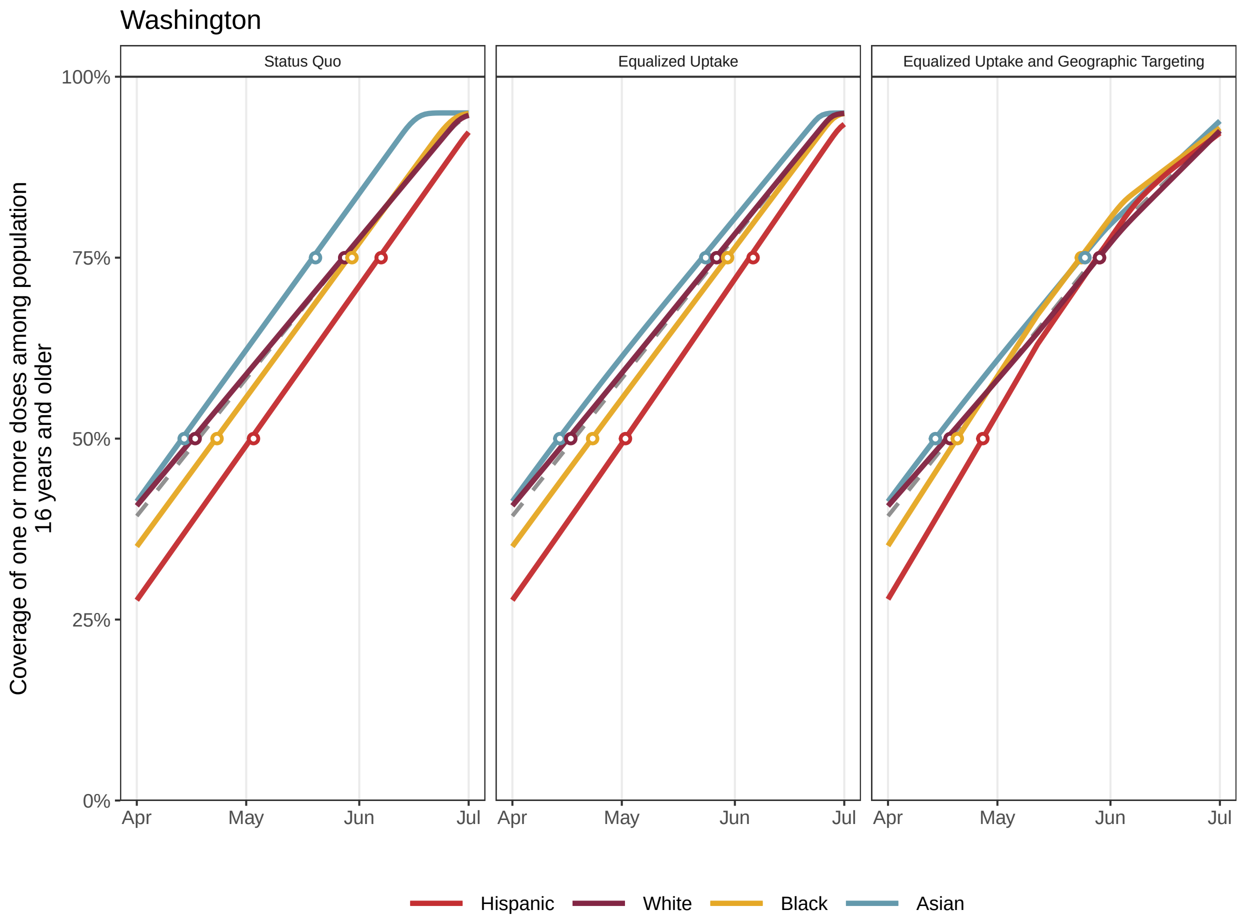


Dashed line shows overall coverage among the Washington population aged 16 years and older

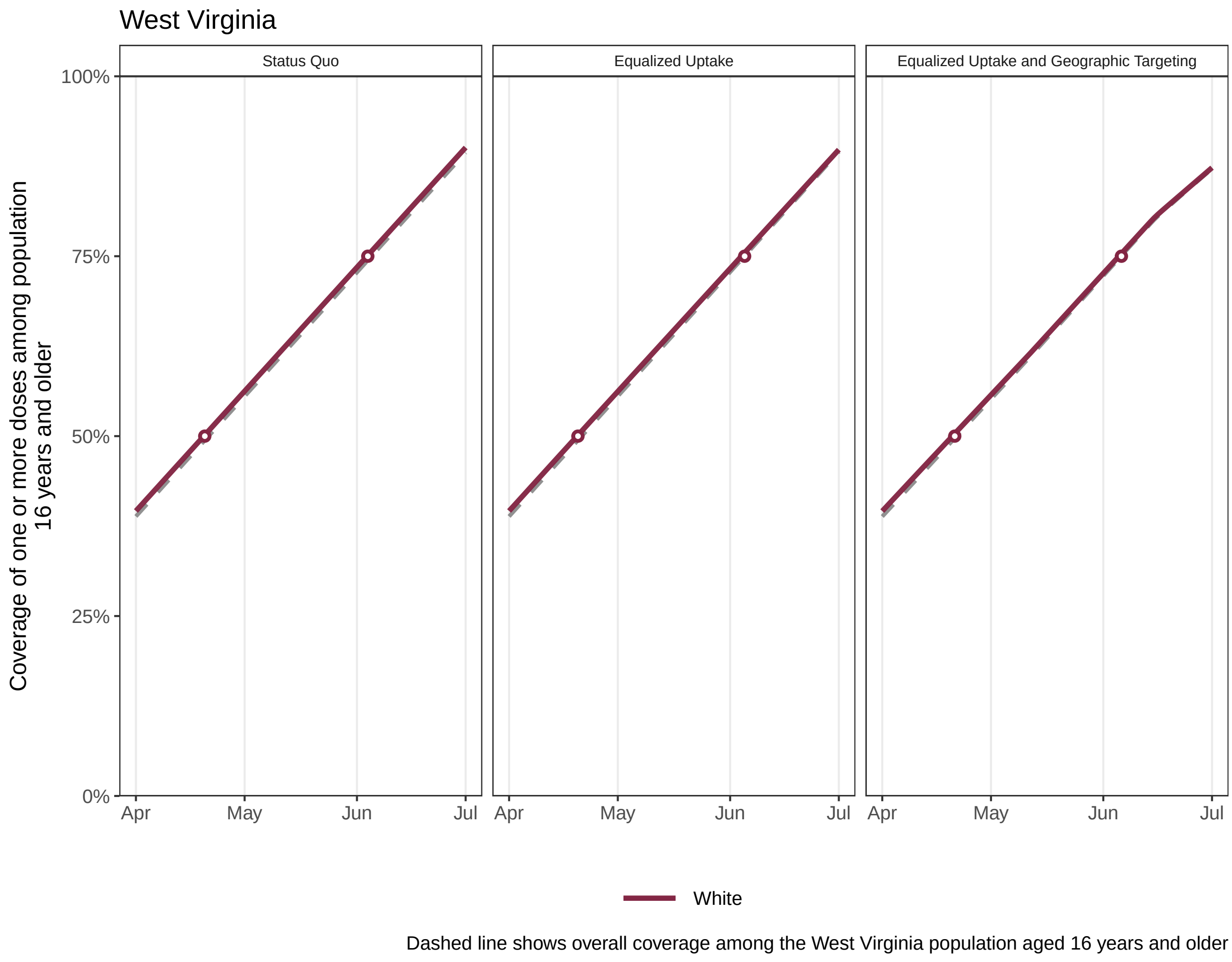
West Virginia
Status Quo
Equalized Uptake
Equalized Uptake and Geographic Targeting
Coverage of one or more doses among population 16 years and older
100%
75%
50%
25%
0%
Apr
May
Jun
Jul
White
Dashed line shows overall coverage among the West Virginia population aged 16 years and older

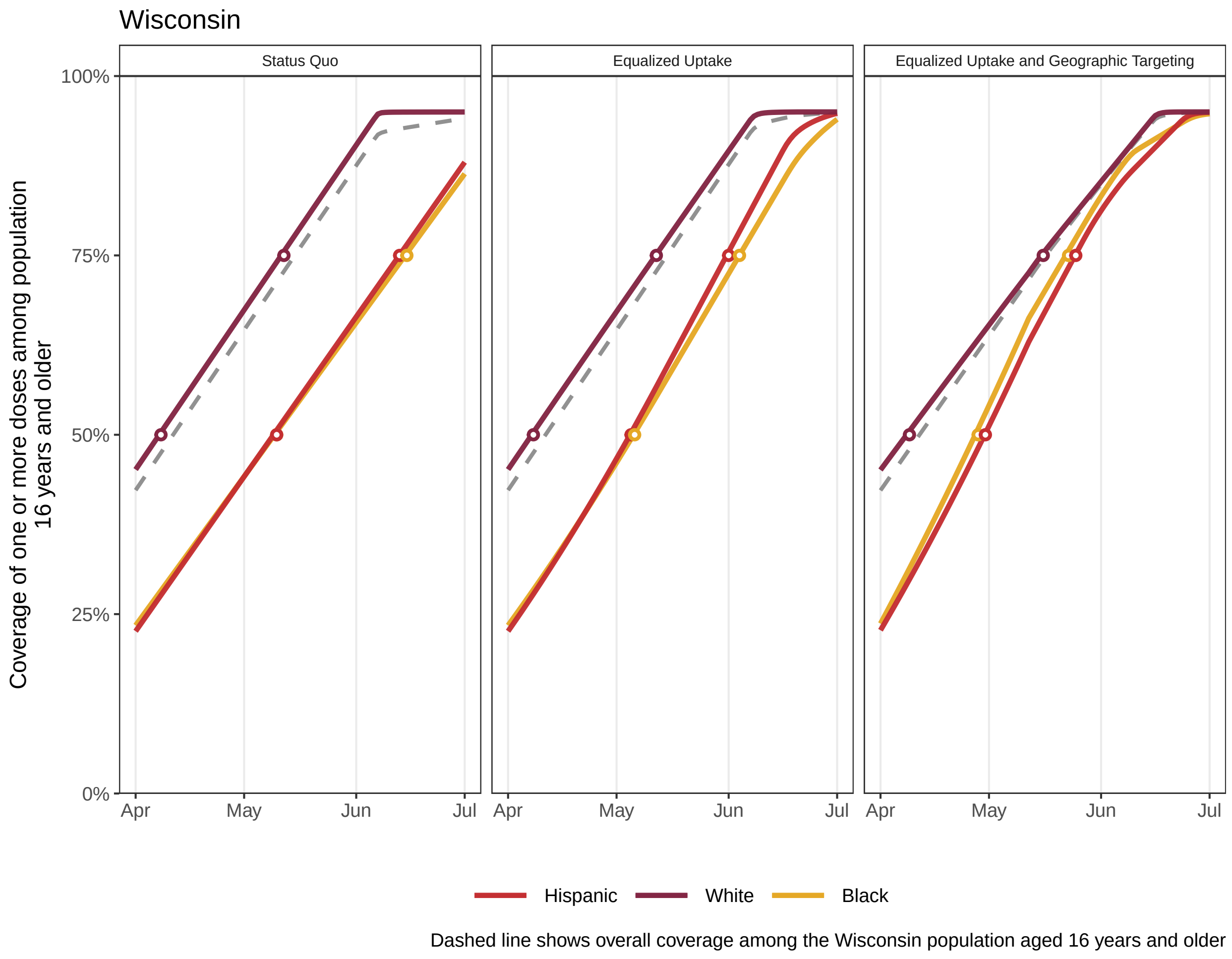

Wisconsin
Status Quo
Equalized Uptake
Equalized Uptake and Geographic Targeting
100%
75%
50%
25%
0%
Coverage of one or more doses among population 16 years and older
Apr
May
Jun
Jul
Hispanic
White
Black
Dashed line shows overall coverage among the Wisconsin population aged 16 years and older

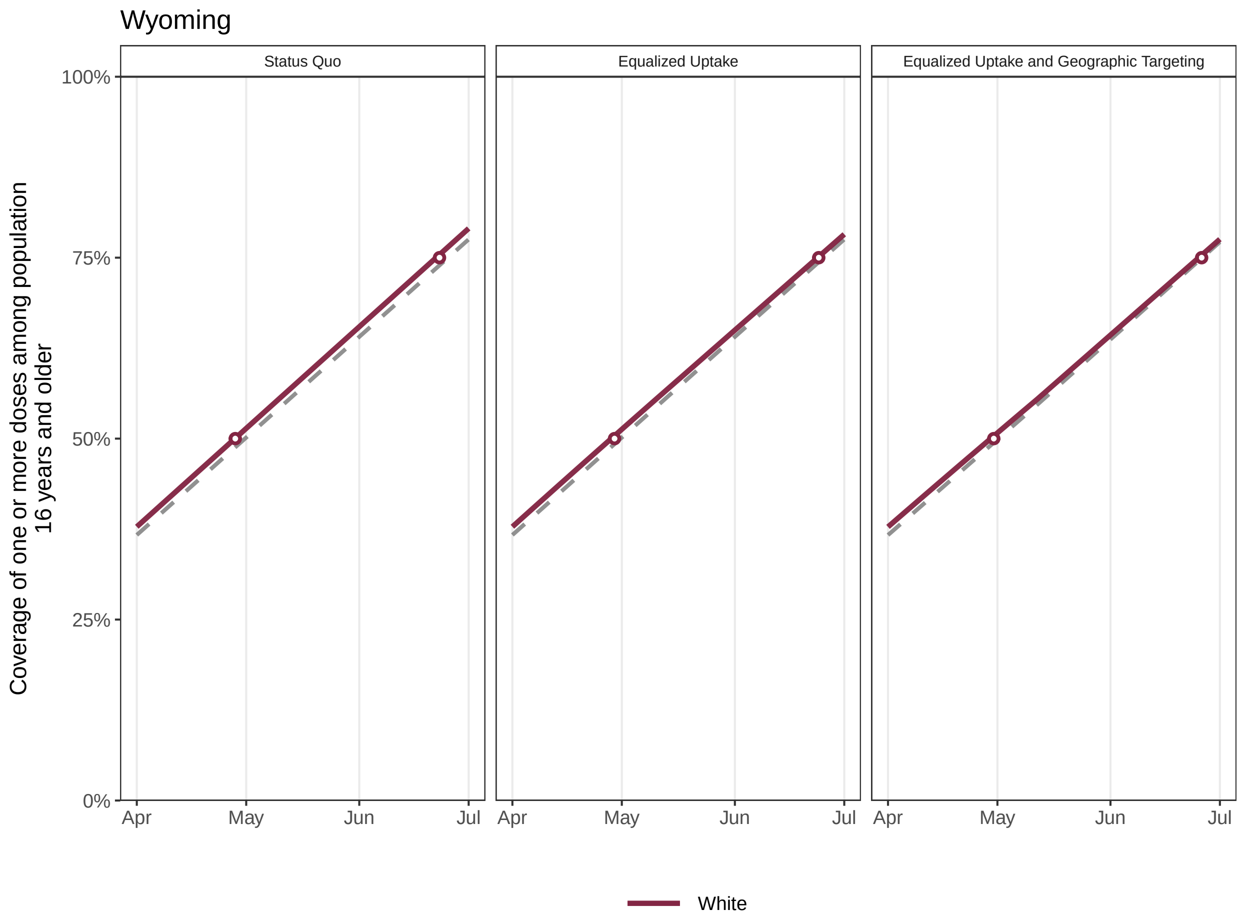


Dashed line shows overall coverage among the Wyoming population aged 16 years and older
